# Phenome-wide association of multiallelic copy number variation in 422,170 UK Biobank individuals reveals novel genetic loci associated with disease

**DOI:** 10.64898/2026.06.03.26354825

**Authors:** Micaela Eisenberg, Richard Packer, Nick Shrine, German Demidov, Harry Pack, Edward J. Hollox, Katherine Fawcett

**Author notes:** Contributed equally (Edward J. Hollox, Katherine Fawcett). Correspondence to, Prof Ed Hollox, Dr Katherine Fawcett.

## Abstract

The contribution of multi-allelic CNVs (mCNVs) to disease risk has not been widely studied. This is largely because they have been difficult to characterise at a large-scale genome-wide, and are often not strongly associated with flanking SNVs, limiting imputation. Improved understanding of the role of mCNVs in disease risk could lead to novel insights into the pathobiology of disease. We robustly typed 69 mCNVs from UK Biobank whole exome sequences in discovery (n=150,682) and replication sets (n=269,317). Discovery and replication PheWAS used clinically-curated composite phenotypes by integrating self-report, primary and secondary health care data to interrogate these variants, for unrelated British individuals of African, European and Central/South Asian ancestries. 173 mCNV-phenotype associations were detected from 26 mCNVs, of which 114 associations replicated. One of eight potentially novel mCNV-phenotype signals was independent of neighbouring associated SNVs, the association of Sulfotransferase 1A1 and 1A2 genes (*SULT1A1*/*SULT1A2)* with estimated glomerular filtration rate (eGFR) in individuals of European ancestry (meta-analysed p=1.05×10^-9^, beta=0.016 [0.011; 0.021]). Other potentially novel associations include Golgi phosphoprotein 3 (*GOLPH3)* with the cardiovascular phenotype bundle branch block in individuals of South Asian ancestry (meta-analysed p=3.35×10^-6^, OR=2.13 [1.53, 2.96]) and alpha amylase 2B (*AMY2B*) with ventricular fibrillation and flutter in individuals of European ancestry (meta-analysed p=2.48×10^-6^, OR=1.50 [1.26; 1.78]). In summary, we show that accurate typing of biobank-scale sample sizes can identify associations between traits and mCNVs, acting through a gene dosage relationship. Our work provides several novel likely causative variants contributing to particular traits of clinical importance and immediately suggest a putative functional mechanism for the observed associations.

## Introduction

Structural variants account for a substantial amount of genetic variation, with estimates from long read sequencing suggesting that structural variants account for between 5-10 Mb of sequence variation in a genome ^1–3^. Given the extent of structural variation in the genome, its importance in influencing diseases and other traits has been of considerable interest. The role of structural variants in rare diseases ^4^ and neurodevelopmental disorders ^5,6^ is well established, but the contribution of structural variants to common diseases has been much more challenging to determine ^7^.

Multiallelic copy number variants (mCNVs) are a subset of structural variants with more than two copy number alleles segregating in a population. This generates a series of diploid genotypes, each typically differing from the next by a single copy. For example, the beta-defensin locus has five different copy number alleles at polymorphic frequency (>1%) which combine to give genotypes ranging from 1 copy per diploid genome to 8 copies per diplod genome ^8^. The role of mCNVs in disease remains unclear, due to limited datasets and challenges in reliably detecting differences in DNA dosage. Previous studies have often relied upon lower-resolution single nucleotide variant (SNV) array data with a low signal:noise ratio ^9^ to call mCNVs. Furthermore, mCNVs can have limited or complex patterns of linkage disequilibrium with neighbouring SNV alleles ^10^, meaning that any disease or trait association caused by the mCNV may not be effectively discovered through association of flanking SNVs tested in a more conventional SNV-based genome-wide association study (GWAS). Accurate typing of mCNVs in large cohorts may therefore reveal novel associations with common diseases and traits. Moreover, mCNVs often span large genomic regions containing whole genes or disrupting parts of genes, suggesting that they are more likely than SNVs to be the functional variant.

To date, there have been few convincing, replicated associations of mCNVs with complex disease. A mCNV affecting multiple genes at the beta-defensin locus has been associated with psoriasis ^11,12^, and a mCNV affecting the complement factor H gene (*CFH*) has been associated with age-related macular degeneration ^13,14^. Most convincingly, mCNV at the complement C4 genes *C4A* and *C4B* has been shown to contribute to susceptibility to schizophrenia ^15^. Most mCNV associations have some degree of flanking SNV support from GWAS, and have been followed by detailed follow-up study of variation at the individual locus.

There have been several previous studies examining CNVs more generally with complex disease using biobank scale datasets. For example, deletions and duplications detected using whole exome sequences of about 200,000 individuals of European ancestries have been tested for association with 78 phenotypes based on ICD10 codes, revealing 862 potential associations, and illustrating the value of directly typing CNVs in genetic association studies ^16^. Subsequent analyses have used the full UK Biobank exome sequencing data to identify rare CNVs, again focusing on individuals of European ancestry, and identifying associations of CNVs with common traits and diseases ^17^. A smaller study using 35,000 individuals focused on mCNVs detected using whole genome sequences, and identified 63 associations with variable number tandem repeats (VNTRs) and multicopy genes ^18^.

Most recently, CNVs called from whole genome sequences of individuals from UK Biobank were used in a large-scale phenome-wide association study (PheWAS). By using paired-end mapping and sequence read depth approaches implemented in DRAGEN, 80,147 copy number gains and 102,717 copy number losses larger than 10kb were called on the autosomes, and the subsequent PheWAS revealed extensive associations with binary and quantitative traits, supporting the role of CNVs affecting common disease ^19^.

In this study we advance the field by taking an approach complementary to the previous studies. We focus on determining at scale, in the UK Biobank whole exome sequencing (WES) dataset, precise copy numbers of 69 mCNVs across autosomes and the X chromosome that directly disrupt entire, or parts of, genes. Typing mCNVs with a bin size of one exon allows for detection of smaller, more common variants. We then conduct a PheWAS on up to 1,701 phenotypes derived from both UK Biobank data and linked primary healthcare data. We apply this approach not only to those UK individuals of European ancestry, but of non-European ancestries as well.

## Methods

### UK Biobank

UK Biobank (UKB) is a large-scale population-based study of approximately 500,000 UK volunteers aged 40-69 years old at the time of recruitment and from varying ethnic and sociodemographic backgrounds ^20,21^. The cohort is longitudinal, with linked electronic healthcare data from secondary care (full cohort), primary care (approximately 45% of the cohort) and repeated baseline assessments (approximately 4%). UK Biobank obtained ethical approval from the North West Multi-centre Research Ethics Committee and informed consent was obtained for all participants. The UK Biobank protocols are overseen by the UK Biobank Ethics Advisory Committee (https://www.ukbiobank.ac.uk/ethics/).

This study was conducted under application IDs 93555, 43027 and 56607 and used 469,952 whole exome sequences (Supplementary methods) which were generated from blood samples collected at the baseline visit ^22^.

### mCNV calling and quality control

We typed mCNVs in UK Biobank whole exome sequences with ClinCNV as previously described ^23^. ClinCNV is a read depth-based caller which employs Gaussian mixture modelling to infer copy number of loci in individuals from a distribution of normalised short-read sequence coverage. We defined multiallelic CNVs (mCNVs) as loci where more than 5% of individuals were not diploid, and more than three different diploid copy numbers were present across the cohort. Our initial analysis of 200,635 exome sequences identified 75 mCNV loci, each confirmed by visual inspection of read depth and observation of clear clustering of different sequence read depths across the samples, each cluster representing a distinct diploid copy number for that locus (Supplementary table 1, Supplementary figure 1). These exome sequences were from an initial batch of 49,953 exomes (Release 1) together with a subsequent batch of 150,682 exomes (Release 2, Supplementary methods). Analysis of these batches separately as a quality control step highlighted substantial differences in CNV detection and read depth profiles between the two batches, possibly because a different batch of oligonucleotide probes was used for Release 1 exomes compared to the remaining samples (Supplementary Methods). Because of this observation, and that individuals included in Release 1 were not randomly selected ^24^, we subsequently excluded Release 1 results from our PheWAS, and focused on the 71 mCNVs reliably typed on Release 2 exomes (Supplementary table 1).

Copy number allele frequencies and the F-statistic for each of the 71 loci were estimated from diploid copy number counts using CNVice ^25^. The F-statistic quantifies how much the observed homozygosity at a locus deviates from that expected under Hardy–Weinberg equilibrium (Supplementary file 1).

The 71 loci were typed using ClinCNV in a replication cohort of a further batch of exome sequences (Release 3, n = 269,317). We checked for typing consistency between the Release 2 and 3 exomes with chi square tests and correcting for overinflation of copy numbers relative to the discovery cohort (n = 17) (Supplementary figure 2, Supplementary methods). We removed two loci which did not have consistent typing between the Release 2 and 3 cohorts, as the clustering patterns of copy number plots were distinct from each other (Supplementary figure 3). 69 loci remained for subsequent phenome-wide association analysis.

### Phenome-wide association analysis

A phenome-wide association study (PheWAS) was conducted in two stages with DeepPheWAS ^26^, stratified by ancestry (relative to 1000 Genomes Project and Human Genome Diversity Project reference populations as previously determined by Karczewski and colleagues ^27^) as well as by sex for X linked loci (as described in the Supplementary Methods). The included health relevant phenotypes spanned primary care, anthropometry, disease status and biomarker levels. The discovery PheWAS (Release 2) was conducted for 66 autosomal mCNVs in the 131,139 European (EUR), 1,943 African (AFR) and 3,085 Central/South Asian (CSA) individuals (Supplementary table 3). Covariates for association were age, sex and the first 10 principal components derived from genome-wide SNV data (generated by Karczewski and colleagues ^27^). Population specific false discovery rate (FDR) thresholds were set to define the significance of mCNV-phenotype associations. A FDR ≤ 0.05 threshold was used in the AFR and CSA groups, while this threshold was adjusted to FDR ≤ 0.01 for the larger EUR group. For five loci on chromosome X, we conducted a sex stratified PheWAS (Supplementary table 4), including covariates of age and 10 principal components. We attempted to replicate all mCNV-phenotype pairs which passed the significance threshold by repeating the PheWAS approach on the replication cohort (Release 3) for 69 mCNV loci with consistent typing, comprising of 65 autosomal mCNVs and 4 loci on chromosome X, with 236,045 EUR participants, 4,622 CSA participants and 3,578 AFR participants (Supplementary table 3).

An association from the replication cohort was prioritised for follow up where the p-value for association in stage 2 was less than 0.05 and the direction of effect was the same as the ancestry matched discovery PheWAS. A mCNV-phenotype association was deemed novel if it had not been reported previously and overlapping gene(s) had not been associated with the phenotype in SNV-based studies, where the SNV was assigned to the nearest gene (according to GWAS Catalog and the ExPheWAS browser). Where there was a single novel association for a particular mCNV, that mCNV-phenotype association was carried forward for further investigation. For loci with more than one novel association, an association with the smallest p-value was selected from each phenotype category.

To test whether these associations were statistically independent of nearby SNVs that associated with the same phenotypes, we conditioned the mCNV-phenotype association on the top imputed SNV associations with the same trait in the Release 3 cohort. To identify candidate SNVs, we used REGENIE ^28^ to test the association of the relevant trait with quality controlled SNVs occurring within 2Mb of the mCNV coordinates. The SNVs showing independent associations with the trait were selected by Genome-wide Complex Trait Analysis (GCTA) conditional and joint analysis ^29,30^, in a stepwise repeated process of identifying the SNV with the most significant p-value and conditioning the remaining variants on this variant until no more associations remained. As this analysis was focused on specific regions of interest, the significance threshold was relaxed to include associations which had reached suggestive significance (conditioned p value < 1×10^-5^). The SNVs were included as covariates in the regression of the mCNV and the associated phenotype. mCNVs were considered statistically independent of the SNV where the conditioned p value remained significant (p<0.05) (Supplementary methods).

### Expression analysis of genes within mCNVs

The 71 mCNVs typed in the discovery cohort were typed in 837 whole genome sequences from the Genotype-Tissue Expression (GTEx) cohort, comprising of 590 samples sequenced using a PCR+ approach, and 247 samples sequenced using a PCR-free approach ^31^. PCR+ and PCR-free samples were typed separately to avoid possible systematic coverage differences resulting from the library preparation protocols. Copy number distributions of mCNVs were checked for clustering and consistency against the UKB copy number distributions by visual inspection, and six loci were excluded. Of these six excluded loci, one locus was not well typed in PCR+ and was monoallelic in PCR-free. For three loci, the locus was not well typed in the PCR-free samples, but well typed in PCR+ samples, so only copy numbers from the PCR+ samples were retained. Two mCNVs affected genes are not included in GTEx v8, and so were excluded. Copy numbers from 63 mCNV loci, which included 84 genes, were carried forward for correlation analysis (Supplementary table 1).

Expression levels for all tissues were extracted from the GTEx v8 bulk tissue expression data from RNA-seq as transcripts per million (TPM) values. For each of the 84 genes affected by a mCNV, copy numbers at the appropriate locus were correlated with gene expression levels via the Pearson correlation coefficient. The Bonferroni corrected p-value significance threshold for the number of tests was p ≤ 1×10^-5^.

Blood plasma protein levels had previously been determined for 54,219 UK Biobank participants ^32^. Of the 2,923 proteins assessed, there were 10 proteins which are coded for by genes overlapping 9 mCNV loci typed in UK Biobank participants. We extracted the proteomics data for these 10 proteins from the baseline visit data, for participants with both proteomics data and mCNV data from Release 3 of exome sequencing (n = 25,746).

The protein level data was expressed as a normalised protein expression level (NPX), where the raw protein levels were log2 transformed for an approximately normal distribution. The Spearman correlation coefficient of mCNV copy number against normalised protein level was calculated for each of the included mCNV-protein pairs. The Bonferroni corrected p-value significance threshold for the number of tests was p ≤ 0.005.

### Locus-specific analysis

To inspect the boundaries of the six prioritised mCNVs, we visualised the read coverage in corresponding whole genome sequences from UK Biobank participants ^33^. Using the Integrated Genomics Viewer (IGV) ^34^, we visualised genomic regions from individuals with the highest and lowest observed copy numbers for the locus. The mCNV boundary was determined as a point where coverage dropped below or increased above the baseline (Supplementary methods).

Given the greater capability of long-read sequencing for detection of larger and more complex SVs, we evaluated deletion and duplication alleles at the six mCNV loci in long read assemblies from the Human Pangenome Reference Consortium (HPRC) ^35^. Dot plots of sequence similarity between HPRC genomes and the T2T-CHM13 reference genome ^36^ for loci of interest were generated with ModDotPlot ^37^, and SVbyeye ^38^ was used to visualise variants of interest. GRCh38 was used as the comparative reference genome for the *GSTM1* gene, as the gene is absent in T2T-CHM13.

### Code availability

Code used to generate the mCNV copy numbers from UK Biobank exome sequences can be found at https://github.com/micaela-eisenberg/mcnv_ukbiobank_wes.

### Data availability

The mCNV copy numbers generated in this work have been returned to UK Biobank (linked to application 93555) and will be available through the UK Biobank Research Analysis Platform. UK Biobank sequencing CRAM files and SNP genotypes are available to registered researchers through the UK Biobank Research Analysis Platform. Supplementary files referenced are located at https://doi.org/10.25392/leicester.data.32221299.

## Results

### Phenome-wide association analyses

We conducted a large-scale PheWAS on robustly typed mCNVs across the UK Biobank. We used a two-stage study design for our analysis, initially calling the selected mCNVs in our discovery cohort, followed by a phenome-wide association analysis on n = 143,781 individuals, and repeating the analysis on a replication cohort of n = 269,317 individuals. These included individuals of different ancestries, grouped by previous analysis ^27^ (Supplementary tables 3, 4) into three broad ancestry groups: a group reflecting majority European ancestry (EUR), a group representing sub-Saharan African ancestry (AFR), and a group representing broad Central and South Asian ancestry (CSA).

Our phenome-wide association study of 66 autosomal mCNVs in the discovery cohort detected a total of 123 associations across 20 mCNVs in the individuals of European ancestry (EUR) group, with false discovery rate (FDR) ≤ 0.01. In the individuals of African ancestry (AFR) group, 22 phenotype associations were identified from three mCNV loci, and in the individuals of Central and South Asian ancestry (CSA) group there were two associations from two mCNVs, with a lower FDR threshold of 0.05 in these smaller ancestry groups. We also detected 26 lipid-related phenotypic associations with a single mCNV mapping to the opsin gene *OPN1LW* in AFR females. No associations with X chromosome mCNVs were detected in males or in individuals of other ancestries (Supplementary files 3, 4).

We tested mCNV-phenotype pairs from the discovery in our replication cohort (n = 269,317). In individuals of European ancestry, 113 associations from 18 autosomal mCNVs were replicated. This included many of the strongest associations detected in the discovery PheWAS, such as chr1:161589510-161591451 (*FCGR2C* gene) with total protein (p = 2.29×10^-62^ in the discovery cohort and p = 2.22×10^-113^ in the replication cohort) and chr1:25272547-25329136 (*RHD* gene) with mean sphered cell volume (p = 9.71×10^-43^ in the discovery cohort and p = 1.31×10^-60^ in the replication cohort). No associations in the AFR ancestry group were replicated. In the CSA ancestry group, the positive association between chr5:32126211-32126636 (*GOLPH3* gene) copy number and risk of the cardiovascular trait bundle branch block replicated (Table 1, Supplementary file 4). Thus, a total of 114 associations replicated in the replication cohort.

**Table 1.** Novel replicated mCNV-phenotype associations.

| Region | Gene(s) | Phenotype | Discovery p-value | Discovery Beta or OR [95% CI] | Replication n p-value | Replication Beta or OR [95% CI] | Group |
| --- | --- | --- | --- | --- | --- | --- | --- |
| chr1:25272547-25329136 | *RHD*,<br>*RSRP1* | Blood Potassium (primary care) | $2.5 \times 10^{-5}$ | 0.028 [0.015; 0.041] | $6.4 \times 10^{-10}$ | 0.031 [0.021; 0.041] | EUR |
| chr1:103571602-103579501 | *AMY2B* | Ventricular fibrillation and flutter | $4.1 \times 10^{-6}$ | 1.9 [1.4; 2.4] | 0.036 | OR 1.3 [1.0; 1.6] | EUR |
| chr1:109690270-109690564 | *GSTM1* | Cholesteryl Esters in Small HDL | $1.8 \times 10^{-4}$ | -0.033 [-0.05; -0.016] | $8.6 \times 10^{-10}$ | -0.041 [-0.054; -0.028] | EUR |
| chr1:109690270-109690564 | *GSTM1* | Cholesterol in Small HDL | $1.4 \times 10^{-4}$ | -0.034 [-0.051; -0.016] | $1.8 \times 10^{-9}$ | -0.04 [-0.053; -0.027] | EUR |
| chr1:109690270-109690564 | *GSTM1* | Cancer of urinary organs (incl. kidney and bladder)<sup>a</sup> | $5.4 \times 10^{-7}$ | 0.84 [0.78; 0.90] | $4.4 \times 10^{-9}$ | OR 0.86 [0.82; 0.91] | EUR |
| chr1:109690270-109690564 | *GSTM1* | Average Diameter for LDL Particles | $3.2 \times 10^{-5}$ | 0.036 [0.019; 0.053] | $1.7 \times 10^{-7}$ | 0.034 [0.021; 0.046] | EUR |
| chr1:109690270-109690564 | *GSTM1* | Free Cholesterol to Total Lipids in Very Large VLDL % | $1.2 \times 10^{-4}$ | 0.036 [0.018; 0.054] | $1.1 \times 10^{-5}$ | 0.031 [0.017; 0.045] | EUR |
| chr1:109690270-109690564 | *GSTM1* | Phospholipids to Total Lipids in Very Large VLDL % | $7.8 \times 10^{-5}$ | 0.037 [0.019; 0.055] | $1.5 \times 10^{-5}$ | 0.03 [0.016; 0.044] | EUR |
| chr1:109690270-109690564 | *GSTM1* | Triglycerides to Total Lipids in Very Large VLDL % | $1.6 \times 10^{-4}$ | -0.031 [-0.048; -0.015] | $6.4 \times 10^{-5}$ | -0.026 [-0.039; -0.013] | EUR |
| chr1:109690270-109690564 | *GSTM1* | Triglycerides to Total Lipids in Large VLDL % | $2.3 \times 10^{-5}$ | -0.037 [-0.055; -0.02] | $1.9 \times 10^{-4}$ | -0.025 [-0.038; -0.012] | EUR |
| chr1:109690270-109690564 | *GSTM1* | Cholesterol to Total Lipids in Large VLDL % | $6.4 \times 10^{-5}$ | 0.035 [0.018; 0.052] | $3.08 \times 10^{-4}$ | 0.024 [0.011; 0.036] | EUR |
| chr1:109690270-109690564 | *GSTM1* | Cholesteryl Esters to Total Lipids in Large VLDL % | $1.6 \times 10^{-4}$ | 0.032 [0.015; 0.049] | $5.5 \times 10^{-4}$ | 0.022 [0.0096; 0.035] | EUR |
| chr1:109690270-109690564 | *GSTM1* | Free Cholesterol to Total Lipids in Large VLDL % | $8.0 \times 10^{-5}$ | 0.035 [0.018; 0.053] | $7.3 \times 10^{-4}$ | 0.023 [0.0095; 0.036] | EUR |
| chr1:109690270-109690564 | *GSTM1* | Phospholipids to Total Lipids in Medium VLDL % | $1.6 \times 10^{-4}$ | 0.032 [0.015; 0.049] | 0.0018 | 0.02 [0.0075; 0.033] | EUR |
| chr1:109690270-109690564 | *GSTM1* | Cholesteryl Esters to Total Lipids in Small VLDL % | $1.2 \times 10^{-5}$ | 0.038 [0.021; 0.056] | 0.0054 | 0.018 [0.0054; 0.031] | EUR |
| chr1:109690270-109690564 | *GSTM1* | Cholesterol to Total Lipids in Small VLDL % | $1.1 \times 10^{-4}$ | 0.034 [0.017; 0.051] | 0.020 | 0.015 [0.0024; 0.028] | EUR |
| chr5:32126211-32126636 | *GOLPH3* | Bundle branch block | $4.0 \times 10^{-6}$ | 3.4 [1.9, 5.5] | 0.021 | OR 1.6 [1.0; 2.4] | CSA |
| chr7:100734168-100738613 | ZAN | Mean corpuscular volume <sup>b</sup> | 3.5x10<sup>-21</sup> | 0.044 [0.035; 0.054] | 3.9x10<sup>-37</sup> | 0.045 [0.038; 0.052] | EUR |
| chr7:100734168-100738613 | ZAN | Mean sphered cell volume <sup>b</sup> | 2.4x10<sup>-7</sup> | 0.025 [0.015; 0.034] | 1.5x10<sup>-12</sup> | 0.025 [0.018; 0.032] | EUR |
| chr7:100734168-100738613 | ZAN | Platelet count <sup>b</sup> | 8.8x10<sup>-7</sup> | 0.023 [0.014; 0.032] | 1.6x10<sup>-12</sup> | 0.024 [0.018; 0.031] | EUR |
| chr7:100734168-100738613 | ZAN | Haemoglobin concentration <sup>b</sup> | 3.0x10<sup>-5</sup> | -0.018 [-0.025; -0.01] | 5.6x10<sup>-11</sup> | -0.019 [-0.024; -0.013] | EUR |
| chr7:100734168-100738613 | ZAN | Mean corpuscular haemoglobin concentration <sup>b</sup> | 9.4x10<sup>-8</sup> | 0.025 [0.016; 0.035] | 1.0x10<sup>-9</sup> | 0.022 [0.015; 0.029] | EUR |
| chr7:100734168-100738613 | ZAN | Red blood cell (erythrocyte) distribution width <sup>b</sup> | 2.2x10<sup>-18</sup> | -0.042 [-0.051; -0.032] | 2.1x10<sup>-9</sup> | -0.021 [-0.028; -0.014] | EUR |
| chr16:28595548-28610191 | *SULT1A1*,<br>*SULT1A2* | eGFR | 2.3x10<sup>-6</sup> | 0.019 [0.011; 0.027] | 1.8x10<sup>-5</sup> | 0.014 [0.0074; 0.02] | EUR |
| chr16:28595548-28610191 | *SULT1A1*,<br>*SULT1A2* | Cholelithiasis and cholecystitis | 1.3x10<sup>-6</sup> | 0.91 [0.87; 0.94] | 0.0020 | OR 0.95 [0.93; 0.98] | EUR |

<sup>a</sup> Although the association of the *GSTM1* null allele with bladder cancer has previously been identified <sup>42,43</sup>, the cancer of the urinary organs phenotype was initially considered separately as it included cancers of the kidneys, bladder, ureter and urethra. However, further investigation revealed that the association signal was driven by the bladder cancer component and no significant associations were detected for cancers of the other urinary organs. <sup>b</sup> Although these associations meet the definition of novelty defined in these methods, they have subsequently been reported <sup>44</sup>.

Of the 19 mCNVs associated with a phenotype following replication, nine are associated with a single phenotype, and ten are pleiotropic, being associated with more than one phenotype (Figure 1a, 1b, 1c). One mCNV, covering the *GSTM1* (Glutathione S-transferase Mu 1) gene, was associated with 42 phenotypes across four phenotype categories: blood biochemistry, blood count, metabolomics and neoplasms (Figure 1a, Figure 1c). These phenotypes included bladder cancer, as well as several measurements of lipid and blood cell levels. Glutathione S-transferase detoxifies electrophilic compounds, including products of oxidative stress and environmental toxins, by conjugating them with glutathione, and copy number is known to correlate with enzyme activity ^39^. The mCNV affecting the *MUC22* (mucin 22) gene is associated with 12 phenotypes across 6 phenotype categories (musculoskeletal, digestive, blood count, spirometry, anthropometry and blood biochemistry) (Figure 1a), however *MUC22* is within the human leucocyte antigen (HLA) region on chromosome 6, and these associations are likely due to linkage disequilibrium between the mCNV and causative variants within HLA genes. 53.5% (n = 61) of all significant associations were categorised under blood biochemistry and blood count. Seventeen of the twenty replicated phenotype associations with the most significant FDR and p-values were within blood biochemistry and blood count.

**Figure 1.**
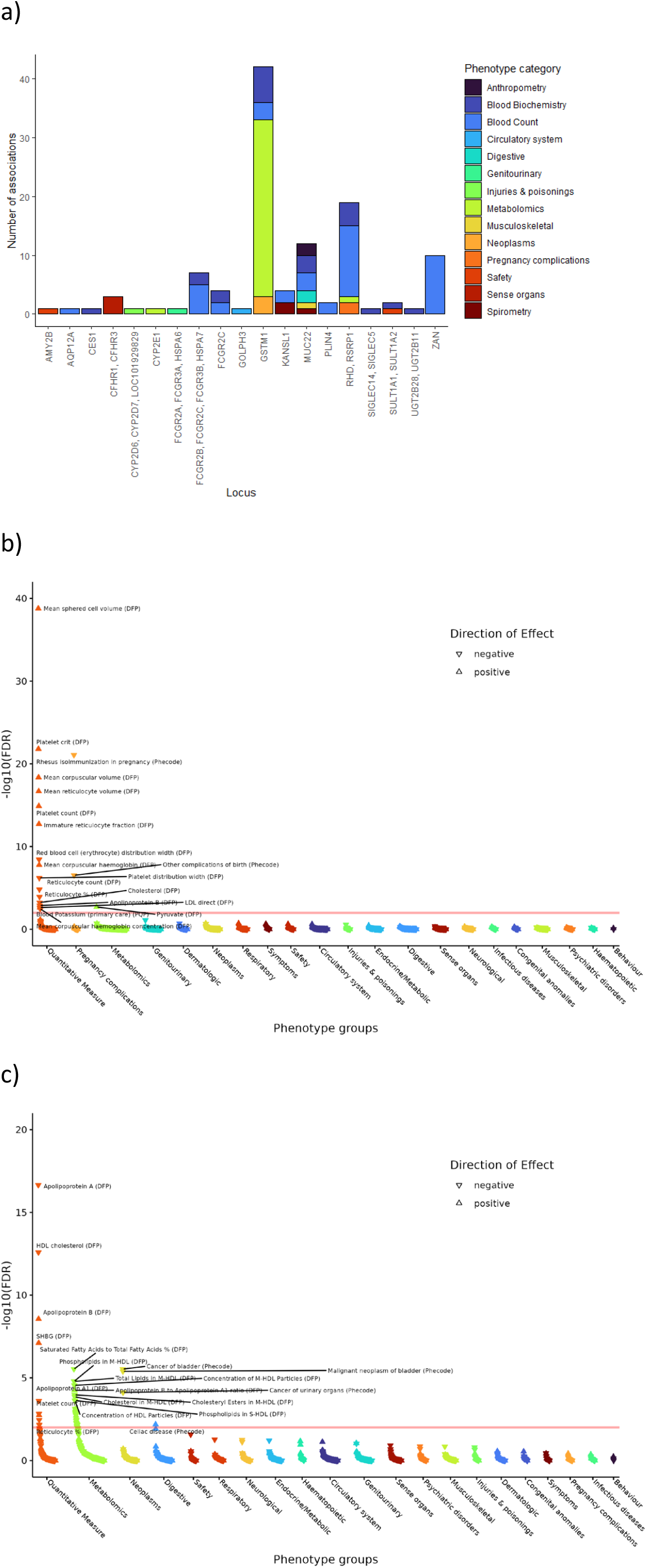
Phenome-wide association study of mCNVs shows extensive pleiotropy. a) Number of phenotypes reproducibly associated with mCNVs (labelled by the genes they affect). Bars are coloured according to the phenotype categories. b) Example PheWAS results for the chr1:25272547-25329136 mCNV, involving the RHD and RSRP1 genes, from individuals of European ancestry from the Release 2 UK Biobank whole exomes (n =131,139). The false discovery rate threshold of 0.01 is indicated by the red line. c) Example PheWAS results for the chr1:109690270-109690564 mCNV, involving the GSTM1 gene, from individuals of European ancestry from the Release 2 UK Biobank whole exomes (n = 131,139). The false discovery rate threshold of 0.01 is indicated by the red line.

We filtered our mCNV-phenotype associations based on novelty by removing associations previously identified in three previous large-scale studies of CNV in the UK Biobank and TopMed cohorts ^16–18^, or where SNV-based GWAS had suggested that particular gene-phenotype association (GWAS Catalog, ^40^, gene-based PheWAS on UK Biobank ^41^). This left 25 novel, replicated, phenotype associations with six mCNVs which we characterised in more detail (Table 1).

### Localisation of association signals

In the Release 3 PheWAS, there were 25 associations from six mCNVs which were potentially novel, the association had a p < 0.05 in the replication analysis and the direction of effect was consistent with the discovery analysis (Table 1). We wished to discover whether the strongest phenotypic association with the mCNV could also be explained by linkage disequilibrium with neighbouring SNVs, as this cannot be inferred directly by detecting an association signal. Eight associations from these six mCNV loci were selected for follow up, where at least one association with the smallest p-value was selected per phenotype category per locus (e.g blood count and neoplasms, where there were several associations for similar phenotypes).

We identified SNVs within a 2Mb window of the mCNV that were also associated with the same phenotype, and then conditioned the mCNV-phenotype association on these associated SNVs by including the SNV as a covariate in the regression model (Supplementary methods). We regarded the mCNV-phenotype association to be statistically independent of the SNV-phenotype association if the p value of the mCNV-phenotype association remained significant in the full model (p<0.05) (Supplementary Tables 5, 6). We were not able to perform conditional analyses for the mCNVs within the *GOLPH3* and *AMY2B* genes, as the signals in the Release 3 cohort were too weak. Similarly, we were unable to analyse the *SULT1A1/SULT1A2* mCNV association with cholelithiasis and cholecystitis, as no SNVs within 2Mb of the mCNV met the selection criteria.

We found one mCNV association signal, *SULT1A1*/*SULT1A2* genes with eGFR, which was independent of neighbouring associated SNVs. Both the mCNV and SNV (rs116938877) at *SULTA1/SULTA2* were still associated with eGFR in the full regression model, suggesting that both contribute to the phenotype. As the other three mCNVs were not independent of the neighbouring variants, the functional variant could not be formally distinguished (Supplementary Tables 5, 6). In the case of the *GSTM1* and *RHD* mCNVs, this was due to high LD between the mCNV and SNV (e.g *GSTM1* and rs140584594 r^2^ = 0.97; *RHD* and rs3079633 r^2^ = 0.96). While the *ZAN* mCNV and rs9801017 were not in linkage disequilibrium (r^2^ = 0.27), these variants were not statistically independent. This suggests that both variants may contribute to phenotypic variation in this locus.

### Genomic structure of phenotype-associated mCNVs

For our six mCNVs selected for follow-up, we wished to understand the extent and genomic structure of the CNV. This would allow us to explore the consequence of different copy number alleles on the gene sequence and predict likely functional consequences of the mCNV. As our study initially called mCNV using sparse exome data, the size of the mCNV was not always known precisely. To address this limitation, we used short read whole genome sequences (WGS) from UK Biobank ^33^ and the full diploid assemblies of samples from the Human Pangenome Reference Consortium (HPRC) ^35^ to fully characterise the six mCNVs with novel genetic associations (Table 2).

**Table 2.** Genomic characterisation of selected mCNVs.

| Gene(s) | WGS coordinates | Diploid copy number range | Size of mCNV unit (kb) | Supported by pangenome assemblies? | Supported by literature? |
| --- | --- | --- | --- | --- | --- |
| *RHD*, *RSRP1* | chr1:25266032-25335866 | 0-4 | 70 | Yes | Yes |
| *AMY2B* | Could not be defined | 0-7 | n/a | No | No |
| *GSTM1* | chr1:109682556-109700514 | 0-4 | 18 | Yes | Yes |
| *GOLPH3* | chr5:32106077-32168303 | 2-6 | 62 | No | Yes |
| *ZAN* | chr7:100729965-100743273 | 0-3 | 13 | Yes | Yes |
| *SULT1A1*,<br>*SULT1A2* | chr16:28602638-28611039 | 0-8 | 8 | Yes | Yes |

For four mCNVs, we could identify zero copy individuals from the UKB replication cohort, indicating homozygosity for a deletion allele. We interrogated the WGS sequence read pileups using the IGV tool to visualise and measure the extent of the deletion allele (Supplementary figures 4-9). Repeating this approach on extreme higher copy number individuals to identify the extent of duplication alleles, which may not be as visually clear as homozygous deletion, confirmed a similar size for three mCNVs. (Supplementary figures 4-9). This might be expected given the reciprocal nature of mCNV generation by non-allelic homologous recombination. The *ZAN* duplication allele was not characterised by extreme enough read depth difference to estimate its boundaries visually (Supplementary figure 8). Additionally, *AMY2B* copy number 0 was not detected in the replication cohort and could not be interrogated visually.

For some common mCNVs, different alleles are represented within the human pangenome reference samples. We identified different alleles using prebuilt pangenome graphs for copy number variable regions ^45^, and generated alignment plots and similarity-based dot plots to visually estimate mCNV size (Supplementary figures 10-21). These estimates were compared to the estimates from short read WGS, and to reports in the literature, if available (Table 2). The WES derived coordinates for all six loci overlapped with WGS estimated coordinates, which were in turn supported by HPRC long read data in 4 of the 6 loci, and by the literature in 5 of 6 loci. This indicated that these mCNVs were true positives.

The *AMY2B* and *GOLPH3* loci were not supported by all three forms of evidence. The *AMY2B* mCNV was typed separately from copy number variation of the adjacent amylase genes with WES. Increased WGS read coverage in both *AMY2B* and *AMY2A* was seen in samples with high *AMY2B* copy number, suggesting that copy numbers of these genes varied together (Supplementary figure 5). A similar pattern was seen in a HPRC sample (Supplementary figure 14), in which the copy number repeat unit included *AMY2B, AMY2A* and *AMY1A*, consistent the previous findings ^46–49^. However, the amylase locus is highly repetitive and enriched with segmental duplications ^46–50^, resulting in poor mapping quality and reads mismapped to the *AMY1* genes with short read sequencing. Visual analysis of WGS read coverage alone could not locate the right hand CNV break point and determine if *AMY1A* occurred in the same repeat unit as the *AMY2* genes (Supplementary figure 4).

The *GOLPH3* mCNV was not identified in the 5 HPRC samples, although it was well characterised in WGS. This was likely a limitation of the sample size as the 100kb window of sequence assessed included *GOLPH3* and the 3’ end of *PDZD2* (Supplementary figure 7), which would be sufficient to detect the mCNV if it was present. This analysis suggests that it is possible to detect true mCNVs with WES, but it is necessary to establish the variant boundaries with whole genome sequencing approaches. Long read sequencing would be preferred for repetitive regions.

We wanted to assess whether the mCNVs are likely to mediate their functional consequences via a gene dosage affect, such that an increase in gene copy number directly increases gene mRNA levels and protein levels. To assess correlation of mCNV copy number with expression levels, we typed 71 mCNVs on the GTEx individuals using ClinCNV on WGS, and correlated with expression levels of the genes overlapping the mCNV (Figure 2, Supplementary figure 22, Supplementary file 5). In general, the data support a gene dosage model for the effect of mCNVs, where 73.8% of correlations across all tissues were positive, irrespective of statistical significance. When correcting for multiple testing (p < 1×10^-5^), 13.4% of correlation across all gene-tissue pairs were positive and statistically significant and 0.72% of these correlations were negative and statistically significant (Supplementary figure 22, Supplementary file 5). When considering all genes, 60 genes (71.4%) had a statistically significant correlation of expression and copy number in at least one tissue (Supplementary file 5). In particular, the *GSTM1* mCNV shows strong positive correlations with *GSTM1* canonical transcript levels (r range across tissues = 0.5-0.97), which agrees with previous work showing positive correlation of *GSTM1* copy number with gene expression ^18^. Others show essentially no correlation, such as *GOLPH3* where 83.3% of the correlations across tissues lie between −0.1 and 0.1 (Supplementary file 5). As the *GOLPH3* mCNV boundaries exclude the gene promoter, this may explain the overall weak effect of mCNV copy number on gene expression (Supplementary figure 7). Genes such as *ZAN* have almost no correlation with expression in the tissue most relevant to the associated phenotype, where whole blood is the most relevant GTEx tissue for mean corpuscular volume (r=0.05, p=0.27. Supplementary file 5). This may be a particular consequence the prioritised mCNVs and might not be the applicable to all mCNVs genome-wide.

**Figure 2.**
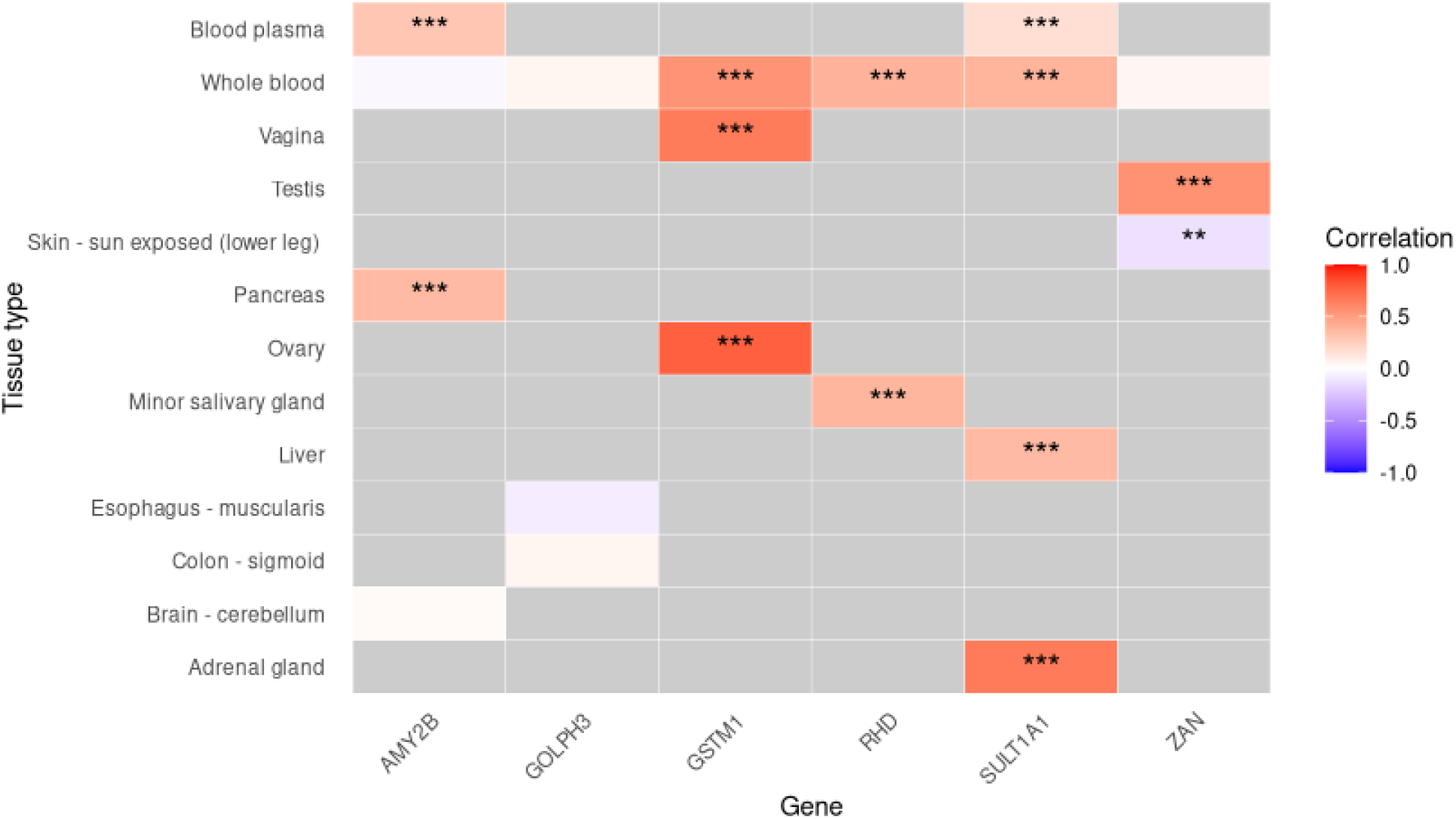
Correlation of expression and copy number of selected mCNVs. Correlation between biological molecule expression level and mCNV copy number, for the two highest-expressing tissues and whole blood, for each gene (Supplementary file 5). The top row is based on UK Biobank proteomic data from blood plasma, other rows are GTEx transcript-level data from RNAseq. Of the genes shown, proteomic data was only available for pancreatic amylase (AMY2B) and sulfotransferase 1A1 (SULT1A1). Asterisks indicate the significance of the correlation where ** indicates p<0.01 and *** indicates p<0.001. No symbol indicates p>0.05.

To assess the effect of mCNVs on protein levels, we used blood plasma proteomic profiles from 54,219 UK Biobank participants ^32^. Of the 2,923 proteins assessed, there were 10 proteins which are coded for by genes annotated to the 9 mCNV loci, expressed as a normalised protein expression level (NPX). For the two prioritised loci where protein levels in blood plasma are detected (pancreatic amylase and sulfotransferase 1A1), there was a significant positive correlation between gene copy number and enzyme levels (p < 5×10^-324^ and p= 4.44×10^-149^ respectively) (Figure 2, Supplementary file 5).

## Discussion

We present here a large-scale analysis of genes showing multiallelic copy number variation in humans. We typed multiallelic CNVs precisely in over 400,000 people from the UK, and tested for association with 2,487 clinically-relevant curated phenotypes from the UK Biobank study and associated primary care data. We discovered 174 mCNV-phenotype associations in our discovery cohort of 136,167 people, 114 of which we subsequently replicated in a cohort of 244,245 individuals. We rediscovered well-known mCNV-phenotype associations, such as *CFHR* with macular degeneration ^13,18,51,52^, and *RHD* with rhesus alloimmunisation and adverse pregnancy outcomes ^16,53–55^. We also found overlap with previous mCNV studies, and gene-phenotype associations previously identified by SNV studies. Taken together, these observations validate the approach used in this study.

Of our 114 mCNV-phenotype associations, 25 were considered novel (Table 1). The *RHD* locus is well-known, as homozygous deletion of the gene causes the rhesus negative blood group phenotype ^53,55^, and duplications at this locus are now well-established. However, in addition to the well-known association with rhesus alloimmunisation and adverse pregnancy outcomes ^53,55^, we show a positive association of *RHD* copy number and blood potassium levels. The mechanism behind this is unclear, but since the RhD protein is an erythrocyte membrane channel, it is possible that variation in the amount of RhD protein in the membrane may affect hemolysis rates, either *in vivo* or during blood draw, thereby increasing leakage of potassium ions into the plasma. *RHD* copy number was positively correlated with gene expression in 48 out of 54 tissue types analysed (Supplementary figure 22).

We found an association of an 18 kb mCNV, affecting the final three exons of *GOLPH3* and the final exon of *PDZD2*, with the cardiovascular phenotype bundle branch block, in UK individuals of central and south Asian ancestry. *GOLPH3* encodes the membrane protein golgi phosphoprotein 3 in the Golgi apparatus, involved in retention and targeting particularly of glycosylation enzymes ^56^. This association was not detected in EUR and AFR participants (AFR replication p=0.99. This phenotype was excluded in the discovery PheWAS as there were too few AFR cases. EUR discovery p=0.04, replication p=0.5). While higher *GOLPH3* mCNV copy number (copy number ≥ 4) was primarily detected in CSA and EUR participants, these high copy numbers were more frequent in CSA participants, e.g copy number 4 occurred at 0.74% in CSA and 0.49% in EUR (Supplementary file 6). These higher frequency copy numbers may in part explain why this association was detectable in CSA and not other groups, despite similar phenotype prevalence in CSA (approximately 3%) and EUR (approximately 2%) groups in the full cohort.

Increased copy number of Sulfotransferase Family 1A Member 1 *SULT1A1* (ranging between 0 to 8 copies per diploid genome) was positively associated with estimated glomerular filtration rate (eGFR, beta=0.019), and reduced risk of cholelithiasis and cholecystitis (gallstones, OR=0.91), and these association signals were independent of neighbouring SNVs. Importantly, the beta values or odds ratios represent risk per extra copy, and, given the extensive copy number range between 0 and 8, individuals with 8 copies represent about two thirds of the gallstone risk compared to a zero-copy individual. GWAS has previously identified the alcohol/hydroxysteroid sulfotransferase gene *SULT2A1*, on chromosome 19, to be associated with gallstone formation^57–59^, likely by affecting the sulfonation of bile acids. In contrast to SULT2, SULT1A1 is a phenolic sulfotransferase which has broad specificity, and, although also expressed in the kidney, it is the most abundant sulfotransferase in the liver ^60^. Although a mechanistic link to gallstone risk is not yet clear, further indication of a link may be seen in Sult1a1 knock out mice, which exhibited abnormal gallbladder morphology or enlarged gallbladder ^61^. *SULT1A1* copy number was correlated with *SULT1A1* mRNA expression in the GTEx kidney cortex (r=0.39, p=1.89×10^-3^) although at nominal significance probably due to small sample size (n=61). In the liver a similar correlation is observed (r=0.36, p=7.98×10^-7^)., strongly significant in a larger sample size (n=181).

The mCNV involving the *ZAN* gene shows association with a variety of red blood cell traits. *ZAN* encodes the protein zonadhesin, which is a sperm membrane protein that mediates species-specificity sperm-egg interaction ^62^. The positive correlation of the mCNV with expression levels of zonadhesin in the testis strongly suggests a consequence for fertility phenotypes not assessed in this PheWAS, and we suggest that this should be assessed in further studies. This mCNV is, however, also distal to the *EPO* gene encoding erythropoietin, the hormone regulating red blood cell production, so the mCNV may affect regulation of this gene. Indeed, the mCNV copy number is significantly correlated with erythropoietin levels in blood serum from the UKB cohort (rho=0.036, p=5.9×10^-8^). Erythropoietin is synthesised primarily in specialised fibroblasts in the border between the medulla and the cortex of the kidney, so the lack of evidence of association of the mCNV with *EPO* gene expression (kidney cortex r=0.144, p=0.267) in the GTEx cohort may be due to limited sampling of the kidney cortex (n=61) and medulla (n=4).

Comparison of the 24 putatively novel mCNV-phenotype associations with findings from two recent CNV-focused PheWAS based on UK Biobank WGS, shows that six associations involving the *ZAN* mCNV have already been reported ^44^ (Table 1). None of the novel associations in this study were detected in the recent PheWAS of CNVs identified by DRAGEN, despite detection of >10kb deletions and/or duplications overlapping the *ZAN, GOLPH3* and *SULT1A1* mCNVs from our study ^19^. Therefore, 18 mCNV associations can still be considered novel. Additionally, 33% of the mCNVs detected in this study do not overlap with reported DRAGEN CNVs ^19^, including the well-established *RHD* deletion, suggesting that the DRAGEN pipeline may have limitations in detecting genome-wide mCNVs of varying length. A strength of our study was the two-stage PheWAS approach with DeepPheWAS which integrates clinically curated phenotypes, primary care data and quantitative biomarker measures with a phecode framework to increase statistical power for detection of more nuanced, clinically relevant phenotypes than standard ICD codes. Indeed, a key contributing factor to the non-overlap of associations with other CNV-focused PheWAS is the definition of phenotypes. Previous studies defined their phenotypes with ICD10 codes rather than incorporate a phecode framework ^19,44^, which combines ICD code derived information for more nuanced and clinically relevant definitions. These studies therefore complement ours in furthering the detection and study of CNVs in population-based analyses.

Interestingly, we find no associations with the *CCL3L1* and *AMY2* loci and UK Biobank phenotypes despite evidence of disease association from previous studies. For example, we find no evidence of any phenotype associated with the mCNV carrying the *CCL3L1* locus, where there has been contradictory evidence of disease association published ^63–67^. Although we do not genotype the *AMY1* mCNV directly, we do type the CNV structurally-related pancreatic amylase *AMY2* CNV ^47,50^, and find no evidence of association with obesity or obesity-related phenotypes ^48^.

There are two general observations from this study. Firstly, across the mCNVs that show novel associations, a gene dosage effect model is generally supported, showing that the most likely mechanism underlying most associations is the effect of gene copy number on protein levels via altering transcript abundance. The correlation between gene copy number and mRNA level in the two highest-expressing tissues is reflected, except for the *ZAN* gene, in whole blood, supporting the use of whole blood in determining the effect of copy number as a proxy for a more disease-relevant tissue. Secondly, we show that mCNVs can show extensive pleiotropic effects, echoing studies in yeast, and emphasising the important role of complex variants in disease ^68^.

We find 18 novel associations of mCNVs with health traits, which include novel and established multiallelic loci. One of the eight prioritised mCNV associations represented an association signal independent of nearby variants, while in four associations, the mCNV and SNV contributed to the same signal. This supports previous findings that the causal variant of reported SNV-phenotype associations could, in some cases, be a structural variant tagged by the SNV ^69^. The remaining three associations were unpowered for conditional analysis in the replication cohort. Although non-novel mCNV associations were not assessed in this study, it is possible that mCNVs and SNVs associating with the same phenotype may be independent and could be investigated in future. Our study, along with the two previously mentioned studies ^19,44^, is one of three PheWAS of mCNVs in a large-scale dataset, that did not focus exclusively on European ancestry participants. These studies contribute towards a growing body of evidence of the role of mCNVs in disease across human populations. Population specific disease aetiologies, as well as shared signals across global populations, can only be revealed when large scale studies intentionally include participants of diverse ancestries.

Although we could not reliably type all known mCNVs that affect genes, given the associations we have found even with the small proportion of all common mCNVs typed, it is likely that many associations remain undiscovered. While real genic mCNVs could be detected with exome sequencing, analysis of short-read whole genome sequences on biobank scale data will undoubtedly enable the accurate genotyping of more mCNVs currently inaccessible through whole exome sequencing. Long read sequencing will also improve genotyping of particular mCNVs and other structural variants, and applying this to even a small proportion of UK Biobank participant genomes will reveal disease associations directly, and indirectly through improved reference panels for structural variants.

## Acknowledgements

This work was supported by a Wellcome Trust DTP PhD studentship awarded to ME (grant number 218505/Z/19/Z). This research used the ALICE High Performance Computing Facility at the University of Leicester and was conducted using the UK Biobank Resource under application numbers 93555, 43027 and 56607. We would like to thank the UK Biobank participants for their contributions.

## Author contributions

ME: formal analysis, investigation, visualisation, conceptualisation, data curation, writing – original draft, writing – review and editing; RJ: methodology, software, writing – review and editing; NS: methodology, software, writing – review and editing; GD: methodology, software, writing – review and editing; HP: visualisation; EJH: conceptualisation, supervision, writing – original draft, writing – review and editing; KF: conceptualisation, supervision, data curation, writing – review and editing

## References

1. Ebert, P., Audano, P.A., Zhu, Q., Rodriguez-Martin, B., Porubsky, D., Bonder, M.J., Sulovari, A., Ebler, J., Zhou, W., Serra Mari, R., et al. (2021). Haplotype-resolved diverse human genomes and integrated analysis of structural variation. Science 372, eabf7117. 10.1126/science.abf7117.

2. Schloissnig, S., Pani, S., Ebler, J., Hain, C., Tsapalou, V., Söylev, A., Hüther, P., Ashraf, H., Prodanov, T., Asparuhova, M., et al. (2025). Structural variation in 1,019 diverse humans based on long-read sequencing. Nature. 10.1038/s41586-025-09290-7.

3. Beyter, D., Ingimundardottir, H., Oddsson, A., Eggertsson, H.P., Bjornsson, E., Jonsson, H., Atlason, B.A., Kristmundsdottir, S., Mehringer, S., Hardarson, M.T., et al. (2021). Long-read sequencing of 3,622 Icelanders provides insight into the role of structural variants in human diseases and other traits. Nature Genetics 53, 779–786. 10.1038/s41588-021-00865-4.

4. Jung, H., Yang, T.-P., Walker, S., Danecek, P., Garcia-Salinas, O.I., Neville, M.D.C., Christopher, J., Cortés-Ciriano, I., Firth, H., Scally, A., et al. (2025). Complex de novo structural variants are an underestimated cause of rare disorders. Nature Communications 16, 9528. 10.1038/s41467-025-64722-2.

5. Hiatt, S.M., Lawlor, J.M.J., Handley, L.H., Latner, D.R., Bonnstetter, Z.T., Finnila, C.R., Thompson, M.L., Boston, L.B., Williams, M., Rodriguez Nunez, I., et al. (2024). Long-read genome sequencing and variant reanalysis increase diagnostic yield in neurodevelopmental disorders. Genome Research 34, 1747–1762.

6. Brandler, W.M., Antaki, D., Gujral, M., Kleiber, M.L., Whitney, J., Maile, M.S., Hong, O., Chapman, T.R., Tan, S., Tandon, P., et al. (2018). Paternally inherited cis-regulatory structural variants are associated with autism. Science 360, 327–331. 10.1126/science.aan2261.

7. Harris, L., McDonagh, E.M., Zhang, X., Fawcett, K., Foreman, A., Daneck, P., Sergouniotis, P.I., Parkinson, H., Mazzarotto, F., Inouye, M., et al. (2025). Genome-wide association testing beyond SNPs. Nature Reviews Genetics 26, 156–170. 10.1038/s41576-024-00778-y.

8. Hollox, E.J., and Abujaber, R. (2017). Evolution and Diversity of Defensins in Vertebrates. In Evolutionary Biology: Self/Nonself Evolution, Species and Complex Traits Evolution, Methods and Concepts, P. Pontarotti, ed. (Springer International Publishing), pp. 27–50. 10.1007/978-3-319-61569-1_2.

9. Pinto, D., Darvishi, K., Shi, X., Rajan, D., Rigler, D., Fitzgerald, T., Lionel, A.C., Thiruvahindrapuram, B., MacDonald, J.R., Mills, R., et al. (2011). Comprehensive assessment of array-based platforms and calling algorithms for detection of copy number variants. Nature Biotechnology 29, 512–520. 10.1038/nbt.1852.

10. Handsaker, R.E., Van Doren, V., Berman, J.R., Genovese, G., Kashin, S., Boettger, L.M., and McCarroll, S.A. (2015). Large multiallelic copy number variations in humans. Nature Genetics 47, 296–303. 10.1038/ng.3200.

11. Stuart, P.E., Hüffmeier, U., Nair, R.P., Palla, R., Tejasvi, T., Schalkwijk, J., Elder, J.T., Reis, A., and Armour, J.A.L. (2012). Association of β-Defensin Copy Number and Psoriasis in Three Cohorts of European Origin. Journal of Investigative Dermatology 132, 2407–2413. 10.1038/jid.2012.191.

12. Hollox, E.J., Huffmeier, U., Zeeuwen, P.L.J.M., Palla, R., Lascorz, J., Rodijk-Olthuis, D., Van De Kerkhof, P.C.M., Traupe, H., De Jongh, G., Heijer, M.D., et al. (2008). Psoriasis is associated with increased β-defensin genomic copy number. Nature Genetics 40, 23–25. 10.1038/ng.2007.48.

13. Hughes, A.E., Orr, N., Esfandiary, H., Diaz-Torres, M., Goodship, T., and Chakravarthy, U. (2006). A common CFH haplotype, with deletion of CFHR1 and CFHR3, is associated with lower risk of age-related macular degeneration. Nat Genet 38, 1173–1177. 10.1038/ng1890.

14. Cantsilieris, S., Nelson, B.J., Huddleston, J., Baker, C., Harshman, L., Penewit, K., Munson, K.M., Sorensen, M., Welch, A.E., Dang, V., et al. (2018). Recurrent structural variation, clustered sites of selection, and disease risk for the complement factor H (CFH) gene family. Proceedings of the National Academy of Sciences 115, E4433–E4442. 10.1073/pnas.1717600115.

15. Sekar, A., Bialas, A.R., de Rivera, H., Davis, A., Hammond, T.R., Kamitaki, N., Tooley, K., Presumey, J., Baum, M., Van Doren, V., et al. (2016). Schizophrenia risk from complex variation of complement component 4. Nature 530, 177–183. 10.1038/nature16549.

16. Fitzgerald, T., and Birney, E. (2022). CNest: A novel copy number association discovery method uncovers 862 new associations from 200,629 whole-exome sequence datasets in the UK Biobank. Cell Genomics 2, 100167. 10.1016/j.xgen.2022.100167.

17. Hujoel, M.L.A., Handsaker, R.E., Sherman, M.A., Kamitaki, N., Barton, A.R., Mukamel, R.E., Terao, C., McCarroll, S.A., and Loh, P.-R. (2024). Protein-altering variants at copy number-variable regions influence diverse human phenotypes. Nature Genetics 56, 569–578. 10.1038/s41588-024-01684-z.

18. Garg, P., Jadhav, B., Lee, W., Rodriguez, O.L., Martin-Trujillo, A., and Sharp, A.J. (2022). A phenome-wide association study identifies effects of copy-number variation of VNTRs and multicopy genes on multiple human traits. The American Journal of Human Genetics 109, 1065–1076. 10.1016/j.ajhg.2022.04.016.

19. Zou, X.Z., Hu, F., Lou, H., Burren, O.S., Li, X., Megy, K., Wheeler, E., Wu, Q., Atanur, S.S., Karpinski, M., et al. (2026). Phenome-wide analysis of copy number variants in 470,727 UK Biobank genomes. Nature. 10.1038/s41586-025-10087-x.

20. Bycroft, C., Freeman, C., Petkova, D., Band, G., Elliott, L.T., Sharp, K., Motyer, A., Vukcevic, D., Delaneau, O., O’Connell, J., et al. (2018). The UK Biobank resource with deep phenotyping and genomic data. Nature 562, 203–209. 10.1038/s41586-018-0579-z.

21. Allen, N.E., Lacey, B., Lawlor, D.A., Pell, J.P., Gallacher, J., Smeeth, L., Elliott, P., Matthews, P.M., Lyons, R.A., Whetton, A.D., et al. (2024). Prospective study design and data analysis in UK Biobank. Science Translational Medicine 16, eadf4428. 10.1126/scitranslmed.adf4428.

22. Backman, J.D., Li, A.H., Marcketta, A., Sun, D., Mbatchou, J., Kessler, M.D., Benner, C., Liu, D., Locke, A.E., Balasubramanian, S., et al. (2021). Exome sequencing and analysis of 454,787 UK Biobank participants. Nature 599, 628–634. 10.1038/s41586-021-04103-z.

23. Fawcett, K.A., Demidov, G., Shrine, N., Paynton, M.L., Ossowski, S., Sayers, I., Wain, L.V., and Hollox, E.J. (2022). Exome-wide analysis of copy number variation shows association of the human leukocyte antigen region with asthma in UK Biobank. BMC Medical Genomics 15. 10.1186/s12920-022-01268-y.

24. Van Hout, C.V., Tachmazidou, I., Backman, J.D., Hoffman, J.D., Liu, D., Pandey, A.K., Gonzaga-Jauregui, C., Khalid, S., Ye, B., Banerjee, N., et al. (2020). Exome sequencing and characterization of 49,960 individuals in the UK Biobank. Nature 586, 749–756. 10.1038/s41586-020-2853-0.

25. Zuccherato, L.W., Schneider, S., Tarazona-Santos, E., Hardwick, R.J., Berg, D.E., Bogle, H., Gouveia, M.H., Machado, L.R., Machado, M., Rodrigues-Soares, F., et al. (2017). Population genetics of immune-related multilocus copy number variation in Native Americans. J R Soc Interface 14. 10.1098/rsif.2017.0057.

26. Packer, R.J., Williams, A.T., Hennah, W., Eisenberg, M.T., Shrine, N., Fawcett, K.A., Pearson, W., Guyatt, A.L., Edris, A., Hollox, E.J., et al. (2023). DeepPheWAS: an R package for phenotype generation and association analysis for phenome-wide association studies. Bioinformatics 39. 10.1093/bioinformatics/btad073.

27. Karczewski, K.J., Gupta, R., Kanai, M., Lu, W., Tsuo, K., Wang, Y., Walters, R.K., Turley, P., Callier, S., Shah, N.N., et al. (2025). Pan-UK Biobank genome-wide association analyses enhance discovery and resolution of ancestry-enriched effects. Nature Genetics. 10.1038/s41588-025-02335-7.

28. Mbatchou, J., Barnard, L., Backman, J., Marcketta, A., Kosmicki, J.A., Ziyatdinov, A., Benner, C., O’Dushlaine, C., Barber, M., Boutkov, B., et al. (2021). Computationally efficient whole-genome regression for quantitative and binary traits. Nature Genetics 53, 1097–1103. 10.1038/s41588-021-00870-7.

29. Yang, J., Ferreira, T., Morris, A.P., Medland, S.E., Madden, P.A.F., Heath, A.C., Martin, N.G., Montgomery, G.W., Weedon, M.N., Loos, R.J., et al. (2012). Conditional and joint multiple-SNP analysis of GWAS summary statistics identifies additional variants influencing complex traits. Nature Genetics 44, 369–375. 10.1038/ng.2213.

30. Yang, J., Lee, S.H., Goddard, M.E., and Visscher, P.M. (2011). GCTA: A Tool for Genome-wide Complex Trait Analysis. The American Journal of Human Genetics 88, 76–82. 10.1016/j.ajhg.2010.11.011.

31. The GTEx Consortium, Aguet, F., Anand, S., Ardlie, K.G., Gabriel, S., Getz, G.A., Graubert, A., Hadley, K., Handsaker, R.E., Huang, K.H., et al. (2020). The GTEx Consortium atlas of genetic regulatory effects across human tissues. Science 369, 1318–1330. 10.1126/science.aaz1776.

32. Sun, B.B., Chiou, J., Traylor, M., Benner, C., Hsu, Y.-H., Richardson, T.G., Surendran, P., Mahajan, A., Robins, C., Vasquez-Grinnell, S.G., et al. (2023). Plasma proteomic associations with genetics and health in the UK Biobank. Nature 622, 329–338. 10.1038/s41586-023-06592-6.

33. Carss, K., Halldorsson, B.V., Hou, L., Liu, J., Wheeler, E., Lo, Y., Kundu, K., Huang, Z., Lacey, B., Dhindsa, R.S., et al. (2025). Whole-genome sequencing of 490,640 UK Biobank participants. Nature. 10.1038/s41586-025-09272-9.

34. Thorvaldsdóttir, H., Robinson, J.T., and Mesirov, J.P. (2013). Integrative Genomics Viewer (IGV): high-performance genomics data visualization and exploration. Briefings in Bioinformatics 14, 178–192. 10.1093/bib/bbs017.

35. Liao, W.-W., Asri, M., Ebler, J., Doerr, D., Haukness, M., Hickey, G., Lu, S., Lucas, J.K., Monlong, J., Abel, H.J., et al. (2023). A draft human pangenome reference. Nature 617, 312–324. 10.1038/s41586-023-05896-x.

36. Nurk, S., Koren, S., Rhie, A., Rautiainen, M., Bzikadze, A.V., Mikheenko, A., Vollger, M.R., Altemose, N., Uralsky, L., Gershman, A., et al. (2022). The complete sequence of a human genome. Science 376, 44–53. 10.1126/science.abj6987.

37. Sweeten, A.P., Schatz, M.C., and Phillippy, A.M. (2024). ModDotPlot—rapid and interactive visualization of tandem repeats. Bioinformatics 40, btae493. 10.1093/bioinformatics/btae493.

38. Porubsky, D., Guitart, X., Yoo, D., Dishuck, P.C., Harvey, W.T., and Eichler, E.E. (2025). SVbyEye: a visual tool to characterize structural variation among whole-genome assemblies. Bioinformatics 41, btaf332. 10.1093/bioinformatics/btaf332.

39. McLellan, R.A., Oscarson, M., Alexandrie, A.-K., Seidegård, J., Evans, D.A.P., Rannug, A., and Ingelman-Sundberg, M. (1997). Characterization of a Human GlutathioneS-Transferase μ Cluster Containing a DuplicatedGSTM1 Gene that Causes Ultrarapid Enzyme Activity. Molecular Pharmacology 52, 958–965. 10.1124/mol.52.6.958.

40. Cerezo, M., Sollis, E., Ji, Y., Lewis, E., Abid, A., Bircan Karatuğ O., Hall, P., Hayhurst, J., John, S., Mosaku, A., et al. (2025). The NHGRI-EBI GWAS Catalog: standards for reusability, sustainability and diversity. Nucleic Acids Research 53, D998–D1005. 10.1093/nar/gkae1070.

41. Legault, M.-A., Perreault, L.-P.L., Tardif, J.-C., and Dubé, M.-P. (2022). ExPheWas: a platform for cis-Mendelian randomization and gene-based association scans. Nucleic Acids Research 50, W305–W311. 10.1093/nar/gkac289.

42. Engel, L.S., Taioli, E., Pfeiffer, R., Garcia-Closas, M., Marcus, P.M., Lan, Q., Boffetta, P., Vineis, P., Autrup, H., Bell, D.A., et al. (2002). Pooled Analysis and Meta-analysis of Glutathione S-Transferase M1 and Bladder Cancer: A HuGE Review. American Journal of Epidemiology 156, 95–109. 10.1093/aje/kwf018.

43. Zhou, T., Li, H.-Y., Xie, W.-J., Zhong, Z., Zhong, H., and Lin, Z.-J. (2018). Association of Glutathione S-transferase gene polymorphism with bladder Cancer susceptibility. BMC Cancer 18, 1088. 10.1186/s12885-018-5014-1.

44. Garg, P., Jadhav, B., Shadrina, M., Martin-Trujillo, A., and Sharp, A.J. (2026). A phenome-wide association study of CNVs genotyped from genome sequencing read depth in the UK Biobank. The American Journal of Human Genetics. 10.1016/j.ajhg.2026.04.013.

45. Li, H., Marin, M., and Farhat, M.R. (2024). Exploring gene content with pangene graphs. Bioinformatics 40, btae456. 10.1093/bioinformatics/btae456.

46. Bolognini, D., Halgren, A., Lou, R.N., Raveane, A., Rocha, J.L., Guarracino, A., Soranzo, N., Chin, C.-S., Garrison, E., and Sudmant, P.H. (2024). Recurrent evolution and selection shape structural diversity at the amylase locus. Nature 634, 617–625. 10.1038/s41586-024-07911-1.

47. Carpenter, D., Dhar, S., Mitchell, L.M., Fu, B., Tyson, J., Shwan, N.A.A., Yang, F., Thomas, M.G., and Armour, J.A.L. (2015). Obesity, starch digestion and amylase: association between copy number variants at human salivary (AMY1) and pancreatic (AMY2) amylase genes. Human Molecular Genetics 24, 3472–3480. 10.1093/hmg/ddv098.

48. Usher, C.L., Handsaker, R.E., Esko, T., Tuke, M.A., Weedon, M.N., Hastie, A.R., Cao, H., Moon, J.E., Kashin, S., Fuchsberger, C., et al. (2015). Structural forms of the human amylase locus and their relationships to SNPs, haplotypes and obesity. Nature Genetics 47, 921–925. 10.1038/ng.3340.

49. Yilmaz, F., Karageorgiou, C., Kim, K., Pajic, P., Scheer, K., Human Genome Structural Variation, C Beck, C.R., Torregrossa, A.-M., Lee, C., Gokcumen, O., et al. (2024). Reconstruction of the human amylase locus reveals ancient duplications seeding modern-day variation. Science 386, eadn0609. 10.1126/science.adn0609.

50. Shwan, N.A.A., Louzada, S., Yang, F., and Armour, J.A.L. (2017). Recurrent Rearrangements of Human Amylase Genes Create Multiple Independent CNV Series. Human Mutation 38, 532–539. 10.1002/humu.23182.

51. Lorés-Motta, L., van Beek, A.E., Willems, E., Zandstra, J., van Mierlo, G., Einhaus, A., Mary, J.L., Stucki, C., Bakker, B., Hoyng, C.B., et al. (2021). Common haplotypes at the CFH locus and low-frequency variants in CFHR2 and CFHR5 associate with systemic FHR concentrations and age-related macular degeneration. Am J Hum Genet 108, 1367–1384. 10.1016/j.ajhg.2021.06.002.

52. Pappas, C.M., Zouache, M.A., Matthews, S., Faust, C.D., Hageman, J.L., Williams, B.L., Richards, B.T., and Hageman, G.S. (2021). Protective chromosome 1q32 haplotypes mitigate risk for age-related macular degeneration associated with the CFH-CFHR5 and ARMS2/HTRA1 loci. Hum Genomics 15, 60. 10.1186/s40246-021-00359-8.

53. Wagner, F.F., and Flegel, W.A. (2000). RHD gene deletion occurred in the Rhesus box. Blood 95, 3662–3668.

54. Urbaniak, S.J., and Greiss, M.A. (2000). RhD haemolytic disease of the fetus and the newborn. Blood Rev 14, 44–61. 10.1054/blre.1999.0123.

55. Colin, Y., Chérif-Zahar, B., Le Van Kim, C., Raynal, V., Van Huffel, V., and Cartron, J.-P. (1991). Genetic Basis of the RhD-Positive and RhD-Negative Blood Group Polymorphism as Determined by Southern Analysis. Blood 78, 2747–2752. 10.1182/blood.V78.10.2747.2747.

56. Welch, L.G., Peak-Chew, S.-Y., Begum, F., Stevens, T.J., and Munro, S. (2021). GOLPH3 and GOLPH3L are broad-spectrum COPI adaptors for sorting into intra-Golgi transport vesicles. Journal of Cell Biology 220, e202106115. 10.1083/jcb.202106115.

57. Fairfield, C.J., Drake, T.M., Pius, R., Bretherick Andrew D., Campbell, A., Clark, D.W., Fallowfield Jonathan A., Hayward, C., Henderson, N.C., Iakovliev, A., et al. (2022). Genome-wide analysis identifies gallstone-susceptibility loci including genes regulating gastrointestinal motility. Hepatology 75.

58. Joshi, A.D., Andersson, C., Buch, S., Stender, S., Noordam, R., Weng, L.-C., Weeke, P.E., Auer, P.L., Boehm, B., Chen, C., et al. (2016). Four Susceptibility Loci for Gallstone Disease Identified in a Meta-analysis of Genome-Wide Association Studies. Gastroenterology 151, 351–363.e328. 10.1053/j.gastro.2016.04.007.

59. Ferkingstad, E., Oddsson, A., Gretarsdottir, S., Benonisdottir, S., Thorleifsson, G., Deaton, A.M., Jonsson, S., Stefansson, O.A., Norddahl, G.L., Zink, F., et al. (2018). Genome-wide association meta-analysis yields 20 loci associated with gallstone disease. Nature Communications 9, 5101. 10.1038/s41467-018-07460-y.

60. Xie, Y., and Xie, W. (2020). The Role of Sulfotransferases in Liver Diseases. Drug Metabolism and Disposition 48, 742–749. 10.1124/dmd.120.000074.

61. Wilson, R., Bülbül Ataç, T., Cheng, T.K., Frost, A., Güneş, O., Kan, M., Keskivali-Bond, P., López Gómez, F., McLaughlin, J., Mucha, J., et al. (2026). International Mouse Phenotyping Consortium Portal: facilitating investigation of gene function and providing insights into human disease. Nucleic Acids Research 54, D1133–D1142. 10.1093/nar/gkaf1148.

62. Tardif, S., Wilson, M.D., Wagner, R., Hunt, P., Gertsenstein, M., Nagy, A., Lobe, C., Koop, B.F., and Hardy, D.M. (2010). Zonadhesin Is Essential for Species Specificity of Sperm Adhesion to the Egg Zona Pellucida*. Journal of Biological Chemistry 285, 24863–24870. 10.1074/jbc.M110.123125.

63. Cantsilieris, S., Western, P.S., Baird, P.N., and White, S.J. (2014). Technical considerations for genotyping multi-allelic copy number variation (CNV), in regions of segmental duplication. BMC Genomics 15, 329. 10.1186/1471-2164-15-329.

64. Field, S.F., Howson, J.M.M., Maier, L.M., Walker, S., Walker, N.M., Smyth, D.J., Armour, J.A.L., Clayton, D.G., and Todd, J.A. (2009). Experimental aspects of copy number variant assays at CCL3L1. Nature Medicine 15, 1115–1117. 10.1038/nm1009-1115.

65. Gonzalez, E., Kulkarni, H., Bolivar, H., Mangano, A., Sanchez, R., Catano, G., Nibbs, R.J., Freedman, B.I., Quinones, M.P., Bamshad, M.J., et al. (2005). The Influence of <i>CCL3L1</i> Gene-Containing Segmental Duplications on HIV-1/AIDS Susceptibility. Science 307, 1434–1440. doi:10.1126/science.1101160.

66. He, W., Kulkarni, H., Castiblanco, J., Shimizu, C., Aluyen, U., Maldonado, R., Carrillo, A., Griffin, M., Lipsitt, A., Beachy, L., et al. (2009). Reply to: “CCL3L1 and HIV/AIDS susceptibility” and “Experimental aspects of copy number variant assays at CCL3L1”. Nature Medicine 15, 1117–1120. 10.1038/nm1009-1117.

67. Bhattacharya, T., Stanton, J., Kim, E.-Y., Kunstman, K.J., Phair, J.P., Jacobson, L.P., and Wolinsky, S.M. (2009). CCL3L1 and HIV/AIDS susceptibility. Nature Medicine 15, 1112–1115. 10.1038/nm1009-1112.

68. Loegler, V., Thiele, P., Teyssonnière, E., Tsouris, A., Brach, G., Cruaud, C., Payen, E., Engelen, S., Dunham, M.J., Hou, J., et al. (2025). From genotype to phenotype with 1,086 near telomere-to-telomere yeast genomes. Nature 648, 649–658. 10.1038/s41586-025-09637-0.

69. Collins, R.L., Brand, H., Karczewski, K.J., Zhao, X., Alföldi, J., Francioli, L.C., Khera, A.V., Lowther, C., Gauthier, L.D., Wang, H., et al. (2020). A structural variation reference for medical and population genetics. Nature 581, 444–451. 10.1038/s41586-020-2287-8.

