## Supplementary Material for "Phenome-wide association of multiallelic copy number variation in 422,170 UK Biobank individuals reveals novel genetic loci associated with disease"

### Supplementary methods

*Whole exome sequences from UK Biobank*

WES have been generated for a total of 469,909 UK Biobank (UKB) participants. The first release of 50,000 whole exome sequences used, for exome enrichment prior to sequencing, oligonucleotide probes from IDT's xGen probe library "lot 1", together with additional probes from NimbleGen VCRome to cover exonic regions which the xGen library did not cover satisfactorily. Seventy-five base pair paired-end reads were sequenced on the Illumina NovaSeq 6000 with the S2 flow cell and reads were processed with the SPB variant calling pipeline which involves aligning the reads to the GRCh38 reference sequence with BWA-MEM [1,2].

The overall methods for exome sequencing were retained for the subsequent batches of whole exome sequences, using "lot 2" of the same oligonucleotide probe collections. These batches were sequenced with the S4 flow cell on the same instrument [3]. Mapping of reads was conducted by the Original Quality Functional Equivalent (OQFE) Pipeline from the 150,000 exome data release onwards [4]. This protocol used BWA-MEM to align reads to the full GRCh38 reference and includes alternate contigs in an alt-aware manner [5]. On average, approximately 95% of the targeted regions were sequenced at 20x coverage or higher [2,3,5].

The WES were released at several time points. The initial release from March 2019 included 50,000 individuals. Another 150,000 participants were released in October 2020 and was followed by a further 100,000 participants in September 2021. The final release of 150,000 participants was released in July 2022. In this study, the March 2019 release is referred to as Release 1 (n = 49,953), the October 2020 release will be referred to as Release 2 (n = 150,682), the September 2021 and July 2022 releases will be referred to together as Release 3 (n = 269,317).

*Determining genetic ancestry*

Genetic ancestry affiliations were previously determined by Karczewski and colleagues using the 1000 Genomes Project data (KGP) and the Human Genome Diversity Panel (HGDP) as reference panels [6]. Ancestry data was available for 430,761 participants across the three exome releases (Supplementary table 2).

*Calling mCNV copy number in the discovery cohort*

The initial data set included the interim 200k release of whole exomes (Release 1 and Release 2, n = 200,635) [7]. Participants from all population ancestries were included. CRAM files containing the raw sequencing reads mapped to the GRCh38 genome build were available for analysis. CNVs were called using ClinCNV (https://github.com/imgag/ClinCNV), a read depth based caller which can be used to detect germline CNVs from WES and WGS [8] (with the parameters lengthG = 0, scoreG = 20, minimumNumOfElemsInCluster = 50). ClinCNV defines a CNV when more than 5% of the sample are not diploid at this site. To call copy numbers, the pipeline generates a statistical model for expected copy numbers, by applying a square root transformation to the median normalised coverage values (normalised coverage = square root of observed copy number divided by the most frequent copy number). Thereafter, constrained Gaussian mixture modelling is applied to determine the copy number from the normalised coverage [9] with read depth modelled as a Poisson distribution. The square root transformation reduces right skewness and stabilises the variance of the read depth-based on read counts. This results in the variance for different copy numbers being approximately the same and the normalised coverage of the most common copy number is centred on 1 [9].

On Release 1 data, ClinCNV identified regions that show evidence of different read depths amongst the samples [7]. The CNVs were confirmed by visual inspection of a plot of each individual's normalised coverage, and individuals were clustered and integer copy number called.

On Release 2 data, ClinCNV was used to call mCNVs using the coordinates of mCNVs identified in the Release 1. The precise coordinates of certain mCNVs differed in the Release 1 and 2 sets (Supplementary table 1), although the mCNVs detected by these coordinates overlapped by a minimum of 92%. The exception to this was the *SPACA5* and *ZNF630* locus, where there was a 3% overlap between the Release 1 and 2 coordinates, as only the Release 1 coordinates extended into the first two exons of the adjacent downstream gene. This locus was not excluded. The mCNVs detected in Release 1 overlapped with 62.5% of common mCNVs identified in phase 3 of the 1000 genomes project [7].

Two loci were removed as copy numbers were absent for the Release 2 participants. One locus (chr7:56832004-56832356) was removed as the distribution of copy numbers was not equivalent between Releases 1 and 2. For example, 2,442 copy number 0 individuals were detected in Release 1 while there were 2 participants in Release 2 with the same diplotype. The *CFHR1/CFHR3* locus (Release 1 coordinates chr1:196830499-196831999 and Release 2 coordinates chr1:196774886-196831999) was initially treated as two separate loci, based on the differing coordinates. These loci were determined to be highly correlated to the point of being identical, and were thereafter treated as a single locus. This left 71 mCNV loci in total (Supplementary table 1).

*Quality control of mCNV copy numbers*

To check if the typing of mCNVs was consistent between the Release 1 and 2 sets, chi square tests were performed. This was to investigate if the proportions of copy number calls from the two cohorts were different. The significance threshold for copy number inconsistency was set at $p < 0.01$ and consistency where $p>0.01$.

*mCNV allele frequency and annotation*

Inbreeding Coefficients Estimation for CNV data (CNVice) (https://github.com/mairarodrigues/cnvice) was used to calculate the copy number allele frequencies from diploid copy numbers, and estimated diploid copy number frequencies for each mCNV, assuming Hardy-Weinberg equilibrium (HWE) and conditioned on the observed diploid copy number frequencies provided to the algorithm. Additionally, CNVice can estimate the F statistic at each locus, to indicate if there is a deviation of the population homozygosity from HWE, allowing diplotype frequencies to depart from HWE [10]. We annotated the mCNVs with RefSeq genes using AnnotSV v3.4.2 [11,12] (accessed June 2025).

*Typing mCNVs in Release 3 exome sequences*

The 71 mCNV loci called on Release 2 were subsequently called using ClinCNV on the 269,317 exome sequences from Release 3. Average read-depth was calculated using ngs-bits (https://github.com/imgag/ngs-bits) for the exome capture regions, with a bin size of one exon, which intersected with the coordinates of the 71 mCNV regions plus 100kb of upstream and downstream flanking sequence. For the loci where the Release 2 coordinates had differed from Release 1, the Release 2 coordinates were retained. The end coordinate of one locus (chr9:66986986-66989121) was changed to that from Release 1 (chr9:66986986-66987520), as the Release 2 coordinates resulted in a plot with a continuous single cluster seemingly artificially subset into inferred copy numbers, rather than the clear and consistent normalised coverage clusters associated with the Release 1 coordinates. Visual inspection of the normalised coverage distribution of each mCNV identified 17 mCNVs where the copy numbers assigned to normalised coverage clusters were multiplied by a constant amount when compared to Release 2 copy numbers. For example, copy number 2 was the most commonly assigned copy number at the *PRAMEF14*/*PRAMEF15*/*PRAMEF19* mCNV locus in Release 2, but the equivalent normalised coverage cluster was assigned as copy number 4 in Release 3. Therefore, these could be corrected by dividing all copy number values by a common factor – in the example case, 2 – and re-clustering to call integer copy number (Supplementary figure 2).

*Phenome-wide association discovery study in autosomes*

Phenome-wide association studies (PheWAS) were performed using the DeepPheWAS R package (https://github.com/Richard-Packer/DeepPheWAS) [13]. Quantitative traits underwent rank-based inverse normal transformation prior to analysis. The phenotype inclusion thresholds were 50 cases for a binary trait and 100 individuals for a quantitative trait, which resulted in 1,701 phenotypes being included in the analysis of EUR participants, 508 phenotypes for the AFR participants, and 603 phenotypes for CSA participants. Participants were filtered using the kinship coefficient previously generated by UKB with Kinship-based Inference for Genome-wide association studies (KING) to remove related individuals where the kinship coefficient > 0.177, representing individuals that are more closely related than second degree relatives [14].

A separate PheWAS was conducted for each ancestry group. For the Release 2 discovery cohort, there were 131,139 European (EUR), 1,943 African (AFR) and 3,085 Central/South Asian (CSA) individuals in this analysis (Supplementary table 3). East Asian (n = 911), Middle Eastern (n = 510) and American individuals (n = 323) were excluded, as the analyses would be insufficiently powered by the small sample sizes.

The covariates for analysis were age, sex and 10 principal components calculated from genome-wide SNV genotype data for ancestry to attempt to account for fine structure within the genetic ancestry groups. The principal components were provided by UKB [15].

A linear regression model was used to analyse continuous traits, while logistic regression was applied to binary traits and each individual's copy number was coded according to their predicted integer diploid copy number for each CNV, assuming a dose-dependent effect of copy number. The false discovery rate (FDR) threshold for significant associations was set at 0.05 for the AFR and CSA groups. With a larger sample size and increased statistical power in the EUR group, a stricter FDR threshold of 0.01 was used.

*Phenome-wide association discovery study on the X chromosome*

A sex-stratified PheWAS approach was utilised to analyse the chromosome X mCNVs. This approach accounted for possible confounding effects of differing X chromosome dosage between sexes on the detected associations by generating sex stratified phenotypes with DeepPheWAS. The phenotype files were generated based on the Release 2 individuals who had mCNV data only.

The samples were then separated by inferred sex, which is based on Affymetrix probe intensity on the X and Y chromosomes, and 176 participants with possible sex chromosome aneuploidies were excluded [15]. Copy numbers at these loci were tested for association with the sex-stratified phenotype data, with separate analyses conducted for inferred genetic males and females in each of the three population groups (Supplementary table 4). Covariates for the analysis were age and 10 principal components of genome-wide SNV genotypes.

*Phenome-wide association replication study*

Associations that were detected in the Release 2 participants were tested by PheWAS in the Release 3 UKB exomes. For this replication analysis, there were 236,045 EUR participants, 4,622 CSA participants and 3,578 AFR participants (Supplementary tables 3, 4). 308 individuals with putative sex chromosome aneuploidies were detected in Release 3, and were excluded from the X linked PheWAS. As before, the mCNV copy number were tested for association with all available phenotypes, with separate analyses for autosomal and X linked loci in each of the EUR, AFR and CSA ancestry populations.

*Assessment of novelty of replicated associations identified in this study*

To assess novelty, we compared our set of genetic associations with three other CNV association studies. Fitzgerald and Birney performed a GWAS on 200,629 European ancestry individuals from UKB exome Releases 1 and 2 in this study, CNVs were detected genome-wide with CNest, and were tested with 54 binary phenotypes based on ICD codes and 24 quantitative phenotypes using logistic and linear regressions with covariates of sex, age, sequencing batch and 10 PCs [16]. A genome-wide significance threshold of $5x10^{-8}$ was used.

Hujoel et al analysed CNV on 454,682 UKB whole exome sequences from Releases 1, 2 and 3, using a combined read-depth approach and SNV-based haplotype information [17]. These CNVs were associated with 57 quantitative traits using BOLT-LMM [18], where the covariates for association were assessment centre, genotyping array, exome release, sex, age, age squared and 20 PCs. A genome-wide significance threshold of $5x10^{-8}$ was used [17].

Garg et al conducted a PheWAS of variable number tandem repeats and multi-copy genes from approximately 35,000 individuals from a variety of studies from the Trans-Omics for Precision Medicine (TOPMed) initiative [19,20]. Participants were stratified into European, South Asian, East Asian, African, or American ancestry. 1,105 multi-copy genes were identified with CNVnator v0.4.1 from PCR-free WGS from 645 samples from the Human Genome Diversity panel [21,22]. These genes were genotyped in the TOPMed WGS samples using mosdepth [23]. 283 quantitative and binary phenotypes were included in this study. Discovery PheWAS were performed with REGENIE [24] with covariates of age, sex and three principal components for ancestry. The PheWAS analyses were divided by the TOPMed substudy and ancestry group, where there was sufficient unrelated samples for association testing and meta-analysed with METAL [25]. An association was considered significant in the discovery analysis where FDR < 0.01. The genotyping and PheWAS approach was replicated for 11 quantitative traits in 10,127 participants from the TopMed BioMe cohort [20].

In addition to CNV-based studies, we analysed evidence of prior association between genes and traits from published SNV-based GWASs. Two SNV-based datasets were utilised as reference for novelty of mCNV-phenotype association. Firstly, a gene-based PheWAS was consulted [26], where collections of common SNV genotypes (minor allele frequency ≥ 0.01 and imputation information quality score ≥ 0.6) from within the protein coding and long non-coding RNA genes were utilised as input. The authors included 400,000 unrelated European ancestry UKB individuals and tested for association of 1,746 phenotypes with 10 PCs. The false discovery threshold was set at 0.05 and p-values for gene level associations were Bonferroni corrected for multiple testing.

Secondly, the GWAS Catalog (https://www.ebi.ac.uk/gwas/home) was consulted [27]. The GWAS Catalog is a curated resource of summary statistics from GWAS studies which are primarily based on SNV associations. Although CNV-based summary statistics can be submitted to the GWAS Catalog, these can only be queried by looking up the relevant study number. CNV-based summary statistics are currently not displayed alongside SNV-based GWAS summary statistics at trait, variant and gene levels.

An association was considered as previously detected if, in one of the above sources, an association of the same or similar phenotype was found with: 1) a CNV which intersected with the mCNV coordinates or 2) SNV(s), which were attributed to the genes intersecting with the mCNV. If the mCNV-phenotype association was not described in any of these sources, it was considered novel.

*Prioritising replicated associations for conditional analysis*

From the list of replicated, novel associations, we selected at least one association from each mCNV locus to carry forward to conditional analyses, according to the following procedure:

If a single phenotype associated with a mCNV, that association was included. If more than one phenotype was associated with a mCNV and these represented different phenotype categories, we selected one association, with the smallest p-value, from each category. If there were several similar phenotypes from the same phenotype category associated with the same mCNV (e.g. metabolomic and blood count phenotypes), the association with the smallest p-value was selected. This resulted in eight putatively novel associations to follow up.

The p-values and effect sizes of these associations from the discovery and replication PheWAS were meta-analysed. Fisher's method was utilised to meta-analyse the p-values [28]. This method assumes that the p-values which are being combined are from independent tests, but that they test the same overall hypothesis. As this method does not consider the effect size of association tests, the effect sizes of the associations were meta-analysed via the fixed effects model with the meta R package with the metagen function (https://cran.r-project.org/web/packages/meta/index.html) [29]. The fixed effects model was chosen as the discovery and replication cohorts were derived from the UKB dataset and analysed with the same methods. As such, the same genetic effect was expected to be detected. Inverse variance weighting pooled the effect sizes, such that the PheWAS analysis which had a lower variance and a more precise estimate of the effect size was weighted higher than the analysis with higher variance [30]. As the meta metagen function expects approximately normally distributed input data, input odds ratios for binary traits were log transformed. These values were meta-analysed and converted back to odds ratios by the odds ratio method.

*Identifying SNVs which associated with the same phenotypes as the prioritised mCNVs*

To determine whether SNVs and indels surrounding the mCNVs associated with the same traits in UKB, we tested for association of the prioritised phenotypes with SNV genotypes in the region 1Mb up and downstream of the mCNV coordinates. The genotypes used for this study were sourced from the genotype calling of 784,256 variants in the UK Biobank participants. The first 50,000 participants were genotyped with the Affymetrix UK BiLEVE Axiom array and the remaining 450,000 participants were genotyped by the Affymetrix UKBB Axiom array. A broader set of genotypes were imputed with a merged reference panel of the Haplotype Reference Consortium and the UK10K haplotype resources [15].

Quality control of these SNV genotypes was conducted with PLINK 2.0 (www.cog-genomics.org/plink/2.0/) [31]. Genotypes from the Release 3 individuals were considered in this analysis. The full set of genome-wide variants were filtered for being autosomal, minor allele frequency > 0.01, minor allele count > 100, Hardy-Weinberg equilibrium test p-value > $1x10^{-15}$. Variants were excluded for sample missingness > 0.1 and genotype missingness > 0.1. This filtering process was conducted separately in the EUR and CSA ancestry participants.

The same phenotype data from the PheWAS, which was generated by DeepPheWAS, was utilised for this analysis. Case and control status for binary traits were extracted. Quantitative phenotypes were extracted in the inverse normal transformed form and in the raw phenotype measures. The exception was the blood potassium level phenotype, as this phenotype was defined by DeepPheWAS, and only the normalised measures were available.

Association testing was conducted with REGENIE v3.1.1 to identify associations of SNVs in a 2Mb window around the mCNV boundaries with the phenotypes of interest (https://github.com/rgcgithub/regenie) [24]. As REGENIE assumes that quantitative phenotypes are normally distributed, inverse normal transformed quantitative phenotypes were used. For binary traits, the case-control status for the phenotype was required. For each phenotype, step 1 was run with the leave-one-out chromosome setting and 1,000 variants per block. For step 2, SNVs from the 1Mb window upstream and downstream of the mCNV were tested for association with each phenotype. This included all imputed genotypes from the 2Mb window around the mCNVs. Covariates for the association were age, $age^2$, sex, array and 10 PCs. Post association testing, the variants were filtered for MAF ≥ 0.01 and imputation quality ≥ 0.3.

The Genome-wide Complex Trait Analysis (GCTA) software was utilised to identify independent variants which associate with the phenotype to include as covariates in conditional analysis (https://yanglab.westlake.edu.cn/software/gcta/#Overview) [32,33]. The conditional and joint (GCTA-COJO) method was used to select the SNVs which passed a conditional p-value threshold ($p < 1x10^{-5}$). An ancestry-matched reference population of 10,000 random UK Biobank samples was used to estimate the LD structure of the tested imputed variants in the EUR Release 3 samples. As the CSA group totalled approximately 4,000, the entire sample was utilised as a LD reference panel for CSA Release 3 samples. The summary statistics from the targeted association testing for each phenotype were utilised as input for this step. The SNVs included in the selection process had passed the quality control checks and fell within the 2Mb window of the predicted mCNV start site. SNVs were selected if their conditional p-values < $1x10^{-5}$. This relaxed threshold was selected as the analysis was no longer focused on genome-wide variation; thus the genome-wide significance threshold would be too stringent.

The genotypes for the selected SNVs were extracted from the imputed data with PLINK. The square of the Pearson correlation coefficient ($r^2$) of mCNV copy number against SNV genotype was calculated for each locus, excluding missing genotypes, as an approximation of pairwise linkage disequilibrium between the two variants [34].

*Conditional analysis of prioritised mCNVs*

The conditional analyses were performed in the Release 3 individuals. In all subsequent analyses, logistic regression was applied for binary traits and linear regression was applied for quantitative phenotypes. For the quantitative phenotypes, regressions were performed with inverse normal transformed normalised and raw phenotypes.

Separate regression models were generated for each mCNV and selected SNV at each locus, against the phenotype. Subsequently, the regression of the mCNV copy number against phenotype was performed, where the covariates were age, $age^2$, sex, 10 PCs, and the selected SNV for that mCNV locus. For the *ZAN* locus, where more than one SNV was selected, the mCNV was conditioned on all selected SNVs simultaneously. A mCNV was considered independent of the SNV when the p-value of the conditioned mCNV was less than 0.05. Conversely, where the mCNV p-value exceeded this threshold, the mCNV was considered not independent of the SNV.

Variance inflation factors (VIF) for the regression models which included the mCNV and SNV were calculated, with the car R package (https://cran.r-project.org/package=car) [35]. VIF are a means of detecting multicollinearity in the regression, where predictor variables may be more related to other predictors than the outcome. Convention suggests that a variable with VIF > 5 is highly colinear with other variables in the data, while VIF = 1 suggests that the variable is not colinear with other variables [36]. In this analysis, VIF were calculated for the individual mCNV and SNVs included in the regression, to determine to what extent they were more related to each other than the phenotype.

*Estimating mCNV boundaries in UK Biobank whole genome sequences*

UK Biobank generated whole genome sequences for 490,640 participants with 151bp paired-end sequencing on the Illumina NovaSeq 6000 instruments, using S4 flowcells (v1.0 chemistry). On average, the whole genome was sequenced at 32.5x coverage [37,38]. Reads were aligned to GRCh38 by two separate methods: with the Illumina DRAGEN v3.7.8 pipeline [37], or using BWA-MEM and the Genome Analysis Toolkit (GATK) pipeline [1,39].

We used the GATK WGS CRAM files to define the boundaries of six mCNVs. We selected, where possible, two Release 3 samples for each of the ClinCNV-predicted minimum and maximum copy numbers. Using IGV, we visually inspected read coverage around the predicted mCNV coordinates for at least two samples per locus [40], in the context of annotated segmental duplications [41,42]. The read depth was used a proxy for the copy number of the visualised region. In a diploid genome, regions with two copies exhibit read depth consistent with a baseline level. Increased read depth relative to this baseline indicates the presence of a duplication, as more reads map to that region. Conversely, when fewer reads map to a deletion, which is indicated by decreased read depth relative to the baseline level. Additional samples were included to refine the coordinates of the *RHD* (4 samples with copy number 3) and *AMY2B* mCNVs (4 samples with copy number 8). The mCNV boundary was determined as a point where coverage dropped below or increased above the baseline.

*Estimating mCNV boundaries with long read assemblies from the Human Pangenome Reference Consortium*

Publicly-available long read genome assemblies were assessed to independently determine mCNV boundaries. The Human Pangenome Reference Consortium (HPRC) project generated assembled, phased, diploid genomes for an initial group of 47 participants [43]. To investigate the presence of mCNVs in a subset of HPRC samples, we selected five genomes from individuals from European and African ancestry groups, the two largest population ancestry groups in UKB. This included one European ancestry sample and one random sample from each continental African group, where one population group was represented per country (Supplementary table 7). Although the majority of the associations at these loci were detected in the UKB EUR ancestry group, the prioritised mCNVs were detected in all UKB ancestry groups. Furthermore, the variation present in non-African ancestry samples was likely captured in the African ancestry samples, owing to out of Africa migrations of human populations over evolutionary time [44,45].

At the time of analysis, the sample assemblies were at different stages of completeness. The HG002 reference assembly (February 2022 release) included fully assembled chromosomes and RefSeq gene annotations [46]. The gene annotations for this sample can be accessed through the NCBI genome annotation browser (https://www.ncbi.nlm.nih.gov/datasets/gene/GCA_021950905.1/; https://www.ncbi.nlm.nih.gov/datasets/gene/GCA_021951015.1/). Assemblies from the May 2021 release were available for the remaining samples, where chromosome scaffolds and gene annotations from Ensembl were available [43]. Ensembl gene annotation is automated and may not always be accurate [47]. For example, when *GSTM1* was deleted, *GSTM1* transcripts mapped to the similar *GSTM2* and *GSTM5* genes.

At each locus a 100kb window centred on a protein-coding gene transcript, which the prioritised mCNVs mapped to, was extracted from the UCSC genome browser for the maternal and paternal chromosomes, with the length of the extracted region was sufficient to capture the canonical transcript of each gene. For HG03453, the *RHD* gene was not annotated. Instead, the coordinates of the locus were centred on the *RSRP1* gene, which is upstream of *RHD* in the GRCh38 and T2T-CHM13 reference genomes. RefSeq gene annotations were used for the HG002 maternal and paternal chromosomes. As the *GSTM1* gene was not annotated on the HG002 maternal chromosome, the Ensembl gene annotation from UCSC genome browser was used instead.

ModDotPlot v0.9.3 [48] was used to generate dot plots of each region. A dot plot displays similarity and differences between two queried sequences, which can be used to detect copy number variants. ModDotPlot uses a k-mer based approach to compare sequences of interest and compute average nucleotide identity k-mers, without aligning the sequences [48]. To detect duplications within individual chromosomes, self-self plots for the maternal and paternal chromosomes for each region were generated with the ModDotPlot static mode. Additionally, a comparative plot of the individual's paternal and maternal chromosomes was generated for each region to detect heterozygous deletions.

To estimate the coordinates of the detected deletions and duplications, we selected one example of each variant and generated a dot plot to compare each variant to a reference genome. T2T-CHM13 was used as the reference for the *RHD*, *SULT1A1* and *ZAN* loci. T2T-CHM13 is a gapless reference human genome assembly, which generated complete sequences of the autosomal and X chromosomes by long read sequencing [49]. As T2T-CHM13 fills in previously unresolved gaps in GRCh38 which included repetitive and copy number variable regions, it is the preferred reference for SV calling in long read genome sequencing [49,50]. Since *GSTM1* is absent in T2T-CHM13 [51], *GSTM1* variants were aligned to GRCh38. For deletions, the reference gene sequence was extracted with an additional 10kb of flanking sequence was sufficient to resolve breakpoints. For the duplications visualised, a larger window around the reference gene was required to resolve the breakpoints, which was dependent on the length of the duplication. The flanking sequences were expanded to 25kb for *GSTM1* and 35kb for *SULT1A1*. For ease of visualisation, the reverse complement was extracted for HPRC sequences which were in the opposite orientation to the reference genome.

Two further HPRC samples, HG00741 and HG01106, were selected to investigate the breakpoints of duplications in *RHD* and the amylase locus, which had been detected in prior work on the HPRC samples [43,52]. These two variants were not detected in the initial analysis of the 5 randomly selected HPRC samples. With the UCSC genome browser, we extracted a 200kb window centred on the ENST04980155573.1 *RHD* transcript for HG01106 paternal assembly. Additionally, we extracted an approximately 580kb from the HG00741 paternal assembly. This region included the entire amylase locus, with a total of 180kb of flanking sequence.

To estimate the breakpoints of these mCNVs, we constructed dot plots comparing these variants to T2T-CHM13, and also used SVbyeye on selected chromosomes to visualise mCNV structure [53].

*Typing mCNVs in Genotype-Tissue Expression (GTEx) Project whole genome sequences*

GTEx generated whole genome sequences (WGS) from 869 GTEx donors with 151bp paired end reads, to a median depth of 32x. These were included in the GTEx v8 release. The first group of samples (n = 79) were sequenced on the Illumina HiSeq 2000 with a PCR-based protocol. The subsequent set (n = 820) was sequenced on the Illumina HiSeq X, where library preparation was conducted for separate groups of samples with a PCR-based protocol (PCR+) and a PCR-free protocol. Seventeen samples were sequenced with both protocols and a further 2 samples (GTEX-1269C and GTEX-1399S) were sequenced in duplicate with the PCR-free protocol. In quality control steps performed by GTEx, 30 low quality sequencing replicates were excluded. This was in addition to excluding WGS from donors who had chromosomal abnormalities (autosomal and sex chromosome aneuploidies, as well as >1Mb duplications or deletions), documented sepsis or cerebral palsy. Further exclusion criteria included participants with sex mismatches, participants who were related to another donor, and samples which were analysed with a different pipeline [54]. Sequencing reads were aligned to GRCh38 with BWA-MEM [1], including alternate contigs, the HLA region and decoy sequences [54]. The aligned WGS CRAM files are hosted by the National Human Genome Research Institute Genomic Data Science Analysis, Visualization, and Informatics Lab-space repository and can be accessed through the Terra cloud platform (https://app.terra.bio/). The CRAM files were accessed through dbGAP accession number phs000424.v8.p2 under project #39477.

In this analysis, we included 837 WGS, of which there were 590 PCR+ samples and 247 PCR-free samples. The two duplicate PCR-free WGS were excluded from this analysis. For the 17 samples which were generated through both amplification approaches, the PCR-free WGS was preferred.

The 71 mCNV loci, which were typed in Release 3 of UKB exomes, were typed in the 837 GTEx samples. Average read coverage was generated for 1kb non-overlapping windows of the genome. The 1kb window size was sufficient for typing 55 mCNV loci, however, 16 mCNVs were too small to be typed in this way, and non-overlapping windows of 50bp were generated for the 16 smaller mCNVs, for 100kb upstream and downstream of each one to ensure that each smaller mCNV overlapped at least 2 typing windows.

PCR+ and PCR-free samples were typed separately to avoid possible coverage differences in the different library preparation protocols. Copy number typing was redone for loci where the copy number plot was visually overinflated relative to the distribution in the UKB Release 2 and 3 exomes. Additionally, loci were excluded if there was no clear distinction between copy number cluster in one or both batches. A total of 65 mCNVs were carried forward for gene expression analysis.

*Correlation of gene expression and mCNV copy number in GTEx*

Gene expression measures were sourced from the GTEx v8 bulk tissue RNA-seq data [54]. Genes which were not included in the GTEx v8 data could not be assessed. The TPM values for each gene of interest were extracted from all bulk tissue gene TPM files from GTEx v8. The Pearson correlation coefficient of gene copy number against gene TPM values was calculated for all tissues: where the mCNV was typed in both the PCR+ and PCR-free sets, the samples were pooled together, while only PCR+ samples were included for loci where the mCNV was not well typed in the PCR-free samples. Pearson correlation was used because gene TPMs have been normalised by transcript length, making them suitable for evaluating linear relationships between variables. A total of 84 genes were included in the correlation analysis.

### Supplementary references


1. Li, H. (2013). Aligning sequence reads, clone sequences and assembly contigs with BWA-MEM. arXiv preprint arXiv:1303.3997.
2. Van Hout, C.V., Tachmazidou, I., Backman, J.D., Hoffman, J.D., Liu, D., Pandey, A.K., Gonzaga-Jauregui, C., Khalid, S., Ye, B., Banerjee, N., et al. (2020). Exome sequencing and characterization of 49,960 individuals in the UK Biobank. Nature *586*, 749-756. 10.1038/s41586-020-2853-0.
3. Backman, J.D., Li, A.H., Marcketta, A., Sun, D., Mbatchou, J., Kessler, M.D., Benner, C., Liu, D., Locke, A.E., Balasubramanian, S., et al. (2021). Exome sequencing and analysis of 454,787 UK Biobank participants. Nature *599*, 628-634. 10.1038/s41586-021-04103-z.
4. Krasheninina, O., Hwang, Y.-C., Bai, X., Zalcman, A., Maxwell, E., Reid, J.G., and Salerno, W.J. (2020). Open-source mapping and variant calling for large-scale NGS data from original base-quality scores. bioRxiv, 2020.2012.2015.356360. 10.1101/2020.12.15.356360.
5. Szustakowski, J.D., Balasubramanian, S., Kvikstad, E., Khalid, S., Bronson, P.G., Sasson, A., Wong, E., Liu, D., Wade Davis, J., Haefliger, C., et al. (2021). Advancing human genetics research and drug discovery through exome sequencing of the UK Biobank. Nat Genet *53*, 942-948. 10.1038/s41588-021-00885-0.
6. Karczewski, K.J., Gupta, R., Kanai, M., Lu, W., Tsuo, K., Wang, Y., Walters, R.K., Turley, P., Callier, S., Shah, N.N., et al. (2025). Pan-UK Biobank genome-wide association analyses enhance discovery and resolution of ancestry-enriched effects. Nature Genetics. 10.1038/s41588-025-02335-7.
7. Fawcett, K.A., Demidov, G., Shrine, N., Paynton, M.L., Ossowski, S., Sayers, I., Wain, L.V., and Hollox, E.J. (2022). Exome-wide analysis of copy number variation shows association of the human leukocyte antigen region with asthma in UK Biobank. BMC Medical Genomics *15*. 10.1186/s12920-022-01268-y.
8. Demidov, G., and Ossowski, S. (2019). ClinCNV: novel method for allele-specific somatic copy-number alterations detection. 10.1101/837971.
9. Demidov, G. (2019). Methods for detection of germline and somatic copy-number variants in next generation sequencing data. (Universitat Pompeu Fabra).
10. Zuccherato, L.W., Schneider, S., Tarazona-Santos, E., Hardwick, R.J., Berg, D.E., Bogle, H., Gouveia, M.H., Machado, L.R., Machado, M., Rodrigues-Soares, F., et al. (2017). Population genetics of immune-related multilocus copy number variation in Native Americans. J R Soc Interface *14*. 10.1098/rsif.2017.0057.
11. Geoffroy, V., Herenger, Y., Kress, A., Stoetzel, C., Piton, A., Dollfus, H., and Muller, J. (2018). AnnotSV: an integrated tool for structural variations annotation. Bioinformatics *34*, 3572-3574. 10.1093/bioinformatics/bty304.
12. Geoffroy, V., Guignard, T., Kress, A., Gaillard, J.-B., Solli-Nowlan, T., Schalk, A., Gatinois, V., Dollfus, H., Scheidecker, S., and Muller, J. (2021). AnnotSV and knotAnnotSV: a web server for human structural variations annotations, ranking and analysis. Nucleic Acids Research *49*, W21-W28. 10.1093/nar/gkab402.
13. Packer, R.J., Williams, A.T., Hennah, W., Eisenberg, M.T., Shrine, N., Fawcett, K.A., Pearson, W., Guyatt, A.L., Edris, A., Hollox, E.J., et al. (2023). DeepPheWAS: an R package for phenotype generation and association analysis for phenome-wide association studies. Bioinformatics *39*. 10.1093/bioinformatics/btad073.
14. Manichaikul, A., Mychaleckyj, J.C., Rich, S.S., Daly, K., Sale, M., and Chen, W.M. (2010). Robust relationship inference in genome-wide association studies. Bioinformatics *26*, 2867-2873. 10.1093/bioinformatics/btq559.
15. Bycroft, C., Freeman, C., Petkova, D., Band, G., Elliott, L.T., Sharp, K., Motyer, A., Vukcevic, D., Delaneau, O., O'Connell, J., et al. (2018). The UK Biobank resource with deep phenotyping and genomic data. Nature *562*, 203-209. 10.1038/s41586-018-0579-z.

16. Fitzgerald, T., and Birney, E. (2022). CNest: A novel copy number association discovery method uncovers 862 new associations from 200,629 whole-exome sequence datasets in the UK Biobank. Cell Genomics *2*, 100167. https://doi.org/10.1016/j.xgen.2022.100167.
17. Hujoel, M.L.A., Handsaker, R.E., Sherman, M.A., Kamitaki, N., Barton, A.R., Mukamel, R.E., Terao, C., McCarroll, S.A., and Loh, P.-R. (2024). Protein-altering variants at copy number-variable regions influence diverse human phenotypes. Nature Genetics *56*, 569-578. 10.1038/s41588-024-01684-z.
18. Loh, P.-R., Kichaev, G., Gazal, S., Schoech, A.P., and Price, A.L. (2018). Mixed-model association for biobank-scale datasets. Nature Genetics *50*, 906-908. 10.1038/s41588-018-0144-6.
19. Stilp, A.M., Emery, L.S., Broome, J.G., Buth, E.J., Khan, A.T., Laurie, C.A., Wang, F.F., Wong, Q., Chen, D., D'Augustine, C.M., et al. (2021). A System for Phenotype Harmonization in the National Heart, Lung, and Blood Institute Trans-Omics for Precision Medicine (TOPMed) Program. American Journal of Epidemiology *190*, 1977-1992. 10.1093/aje/kwab115.
20. Garg, P., Jadhav, B., Lee, W., Rodriguez, O.L., Martin-Trujillo, A., and Sharp, A.J. (2022). A phenome-wide association study identifies effects of copy-number variation of VNTRs and multicopy genes on multiple human traits. The American Journal of Human Genetics *109*, 1065-1076. https://doi.org/10.1016/j.ajhg.2022.04.016.
21. Abyzov, A., Urban, A.E., Snyder, M., and Gerstein, M. (2011). CNVnator: an approach to discover, genotype, and characterize typical and atypical CNVs from family and population genome sequencing. Genome Res *21*, 974-984. 10.1101/gr.114876.110.
22. Almarri, M.A., Bergström, A., Prado-Martinez, J., Yang, F., Fu, B., Dunham, A.S., Chen, Y., Hurles, M.E., Tyler-Smith, C., and Xue, Y. (2020). Population Structure, Stratification, and Introgression of Human Structural Variation. Cell *182*, 189-199.e115. https://doi.org/10.1016/j.cell.2020.05.024.
23. Pedersen, B.S., and Quinlan, A.R. (2018). Mosdepth: quick coverage calculation for genomes and exomes. Bioinformatics *34*, 867-868. 10.1093/bioinformatics/btx699.
24. Mbatchou, J., Barnard, L., Backman, J., Marcketta, A., Kosmicki, J.A., Ziyatdinov, A., Benner, C., O'Dushlaine, C., Barber, M., Boutkov, B., et al. (2021). Computationally efficient whole-genome regression for quantitative and binary traits. Nature Genetics *53*, 1097-1103. 10.1038/s41588-021-00870-7.
25. Willer, C.J., Li, Y., and Abecasis, G.R. (2010). METAL: fast and efficient meta-analysis of genomewide association scans. Bioinformatics *26*, 2190-2191. 10.1093/bioinformatics/btq340.
26. Legault, M.-A., Perreault, L.-P.L., Tardif, J.-C., and Dubé, M.-P. (2022). ExPheWas: a platform for cis-Mendelian randomization and gene-based association scans. Nucleic Acids Research *50*, W305-W311. 10.1093/nar/gkac289.
27. Cerezo, M., Sollis, E., Ji, Y., Lewis, E., Abid, A., Bircan, Karatuğ O., Hall, P., Hayhurst, J., John, S., Mosaku, A., et al. (2025). The NHGRI-EBI GWAS Catalog: standards for reusability, sustainability and diversity. Nucleic Acids Research *53*, D998-D1005. 10.1093/nar/gkae1070.
28. Mosteller, F., and Fisher, R.A. (1948). Questions and Answers. The American Statistician *2*, 30-31. 10.2307/2681650.
29. Balduzzi, S., Rücker, G., and Schwarzer, G. (2019). How to perform a meta-analysis with R: a practical tutorial. BMJ Mental Health *22*, 153. 10.1136/ebmental-2019-300117.
30. Borenstein, M., Hedges, L.V., Higgins, J.P.T., and Rothstein, H.R. (2010). A basic introduction to fixed-effect and random-effects models for meta-analysis. Research Synthesis Methods *1*, 97-111. https://doi.org/10.1002/jrsm.12.
31. Chang, C.C., Chow, C.C., Tellier, L.C.A.M., Vattikuti, S., Purcell, S.M., and Lee, J.J. (2015). Second-generation PLINK: rising to the challenge of larger and richer datasets. GigaScience *4*, s13742-13015-10047-13748. 10.1186/s13742-015-0047-8.
32. Yang, J., Ferreira, T., Morris, A.P., Medland, S.E., Madden, P.A.F., Heath, A.C., Martin, N.G., Montgomery, G.W., Weedon, M.N., Loos, R.J., et al. (2012). Conditional and joint multiple-

SNP analysis of GWAS summary statistics identifies additional variants influencing complex traits. Nature Genetics *44*, 369-375. 10.1038/ng.2213.

33. Yang, J., Lee, S.H., Goddard, M.E., and Visscher, P.M. (2011). GCTA: A Tool for Genome-wide Complex Trait Analysis. The American Journal of Human Genetics *88*, 76-82. https://doi.org/10.1016/j.ajhg.2010.11.011.

34. Hill, W.G., and Robertson, A. (1968). Linkage disequilibrium in finite populations. Theoretical and Applied Genetics *38*, 226-231. 10.1007/BF01245622.

35. Fox, J., and Weisberg, S. (2019). An R Companion to Applied Regression, Third Edition (Sage).

36. Kim, J.H. (2019). Multicollinearity and misleading statistical results. Korean J Anesthesiol *72*, 558-569. 10.4097/kja.19087.

37. Li, S., Carss, K.J., Halldorsson, B.V., Cortes, A., and Consortium, U.K.B.W.-G.S. (2023). Whole-genome sequencing of half-a-million UK Biobank participants. medRxiv, 2023.2012.2006.23299426. 10.1101/2023.12.06.23299426.

38. Carss, K., Halldorsson, B.V., Hou, L., Liu, J., Wheeler, E., Lo, Y., Kundu, K., Huang, Z., Lacey, B., Dhindsa, R.S., et al. (2025). Whole-genome sequencing of 490,640 UK Biobank participants. Nature. 10.1038/s41586-025-09272-9.

39. Halldorsson, B.V., Eggertsson, H.P., Moore, K.H.S., Hauswedell, H., Eiriksson, O., Ulfarsson, M.O., Palsson, G., Hardarson, M.T., Oddsson, A., Jensson, B.O., et al. (2022). The sequences of 150,119 genomes in the UK Biobank. Nature *607*, 732-740. 10.1038/s41586-022-04965-x.

40. Robinson, J.T., Thorvaldsdottir, H., Turner, D., and Mesirov, J.P. (2023). igv.js: an embeddable JavaScript implementation of the Integrative Genomics Viewer (IGV). Bioinformatics *39*, btac830. 10.1093/bioinformatics/btac830.

41. Bailey, J.A., Gu, Z., Clark, R.A., Reinert, K., Samonte, R.V., Schwartz, S., Adams, M.D., Myers, E.W., Li, P.W., and Eichler, E.E. (2002). Recent Segmental Duplications in the Human Genome. Science *297*, 1003-1007. 10.1126/science.1072047.

42. Bailey, J.A., Yavor, A.M., Massa, H.F., Trask, B.J., and Eichler, E.E. (2001). Segmental Duplications: Organization and Impact Within the Current Human Genome Project Assembly. Genome Research *11*, 1005-1017. 10.1101/gr.187101.

43. Liao, W.-W., Asri, M., Ebler, J., Doerr, D., Haukness, M., Hickey, G., Lu, S., Lucas, J.K., Monlong, J., Abel, H.J., et al. (2023). A draft human pangenome reference. Nature *617*, 312-324. 10.1038/s41586-023-05896-x.

44. The 1000 Genomes Project Consortium (2015). A global reference for human genetic variation. Nature *526*, 68-74. 10.1038/nature15393.

45. Tucci, S., and Akey, J.M. (2019). The long walk to African genomics. Genome Biology *20*, 130. 10.1186/s13059-019-1740-1.

46. Jarvis, E.D., Formenti, G., Rhie, A., Guarracino, A., Yang, C., Wood, J., Tracey, A., Thibaud-Nissen, F., Vollger, M.R., Porubsky, D., et al. (2022). Semi-automated assembly of high-quality diploid human reference genomes. Nature *611*, 519-531. 10.1038/s41586-022-05325-5.

47. Hubbard, T., Barker, D., Birney, E., Cameron, G., Chen, Y., Clark, L., Cox, T., Cuff, J., Curwen, V., Down, T., et al. (2002). The Ensembl genome database project. Nucleic Acids Research *30*, 38-41. 10.1093/nar/30.1.38.

48. Sweeten, A.P., Schatz, M.C., and Phillippy, A.M. (2024). ModDotPlot—rapid and interactive visualization of tandem repeats. Bioinformatics *40*, btae493. 10.1093/bioinformatics/btae493.

49. Nurk, S., Koren, S., Rhie, A., Rautiainen, M., Bzikadze, A.V., Mikheenko, A., Vollger, M.R., Altemose, N., Uralsky, L., Gershman, A., et al. (2022). The complete sequence of a human genome. Science *376*, 44-53. 10.1126/science.abj6987.

50. Aganezov, S., Yan, S.M., Soto, D.C., Kirsche, M., Zarate, S., Avdeyev, P., Taylor, D.J., Shafin, K., Shumate, A., Xiao, C., et al. (2022). A complete reference genome improves analysis of human genetic variation. Science *376*, eabl3533. 10.1126/science.abl3533.

51. Yang, X., Wang, X., Zou, Y., Zhang, S., Xia, M., Fu, L., Vollger, M.R., Chen, N.-C., Taylor, D.J., Harvey, W.T., et al. (2023). Characterization of large-scale genomic differences in the first complete human genome. Genome Biology *24*, 157. 10.1186/s13059-023-02995-w.
52. Bolognini, D., Halgren, A., Lou, R.N., Raveane, A., Rocha, J.L., Guarracino, A., Soranzo, N., Chin, C.-S., Garrison, E., and Sudmant, P.H. (2024). Recurrent evolution and selection shape structural diversity at the amylase locus. Nature *634*, 617-625. 10.1038/s41586-024-07911-1.
53. Porubsky, D., Guitart, X., Yoo, D., Dishuck, P.C., Harvey, W.T., and Eichler, E.E. (2025). SVbyEye: a visual tool to characterize structural variation among whole-genome assemblies. Bioinformatics *41*, btaf332. 10.1093/bioinformatics/btaf332.
54. The GTEx Consortium, Aguet, F., Anand, S., Ardlie, K.G., Gabriel, S., Getz, G.A., Graubert, A., Hadley, K., Handsaker, R.E., Huang, K.H., et al. (2020). The GTEx Consortium atlas of genetic regulatory effects across human tissues. Science *369*, 1318-1330. 10.1126/science.aaz1776.
55. Zook, J.M., Catoe, D., McDaniel, J., Vang, L., Spies, N., Sidow, A., Weng, Z., Liu, Y., Mason, C.E., Alexander, N., et al. (2016). Extensive sequencing of seven human genomes to characterize benchmark reference materials. Scientific Data *3*, 160025. 10.1038/sdata.2016.25.
56. The International HapMap Consortium (2005). A haplotype map of the human genome. Nature *437*, 1299-1320. 10.1038/nature04226.

**Supplementary table 1:** The coordinates and genes associated with multiallelic copy number variants. Release 2 coordinates were carried forward for subsequent analysis, except where indicated.

| Release 1 GRCh38 coordinates | Gene(s) | Inclusion in Release 2 discovery PheWAS? (Differing coordinates) | Inclusion in Release 3 replication PheWAS? (Differing coordinates) | Inclusion for GTEx expression analysis? |
|---|---|---|---|---|
| chr1:1450683-1496204 | *ATAD3B, ATAD3C* | Yes | Yes | Yes |
| chr1:1703475-1707584 | *CDK11A* | Yes | Yes | No, poor copy number typing |
| chr1:12793227-12795685 | *PRAMEF1* | Yes (chr1:12793227-12795996) | Yes | Yes |
| chr1:13321702-13371900 | *PRAMEF14, PRAMEF15, PRAMEF19* | Yes | Yes | Yes |
| chr1:16048761-16049245 | *CLCNKB* | Yes | Yes | Yes (PCR+ only) |
| chr1:16922018-16948926 | *CROCC* | Yes | Yes | Yes |
| chr1:22003002-22009857 | *CELA3A* | Yes | Yes | Yes |
| chr1:25272547-25329136 | *RHD, RSRP1, LOC105376882* | Yes | Yes | Yes |
| chr1:103571602-103579501 | *AMY2B* | Yes | Yes | Yes |
| chr1:103617440-103618100 | *AMY2A* | Yes (chr1:103617440-103619108) | Yes | Yes |
| chr1:109690270-109690564 | *GSTM1* | Yes | Yes | Yes |
| chr1:143729673-143883065 | *FCGR1CP, H2BP2, LOC105369140, LINC02591, PDE4DIPP3* | Yes (chr1:143729673-143882601) | Yes | Yes |
| chr1:161517974-161549844 | *FCGR2A, FCGR3A, HSPA6* | Yes (chr1:161509819-161549844) | Yes | Yes |
| chr1:161589510-161591451 | *FCGR2C* | Yes | Yes | Yes |
| chr1:161595511-161673229 | *FCGR2B, FCGR2C, FCGR3B, HSPA7* | Yes | Yes | Yes |
| chr1:196830499-196831999 | *CFHR1, CFHR3* | Yes (chr1:196774886-196831999) | Yes | Yes |
| chr2:97118343-97118517 | *ANKRD36* | Yes | Yes | No, poor copy number typing |
| chr2:97532309-97541983 | *ANKRD36B* | Yes (chr2:97511137-97545738) | Yes | Yes |
| chr2:240691913-240692521 | *AQP12A* | Yes | Yes | Yes |
| chr4:69280500-69294809 | *UGT2B28, UGT2B11, LOC105377267* | Yes (chr4:69214001-69294809) | Yes | No, poor copy number typing |
| chr5:801105-825251 | *ZDHHC11* | Yes | Yes | Yes |
| chr5:32126211-32126636 | *GOLPH3* | Yes | Yes | Yes |
| chr5:141174621-141179838 | *PCDHB7, PCDHB8* | Yes | Yes | Yes |
| chr6:311879-350868 | *DUSP22* | Yes | Yes | Yes |

| | | | | |
|---|---|---|---|---|
| chr6:31026054-31027714 | *MUC22* | Yes | Yes | Yes |
| chr6:167951185-167966276 | *AFDN* | Yes | Yes | Yes |
| chr7:56832004-56832356 | ENSG00000279072 | No, different copy number distributions between Releases 1 and 2 | No | No |
| chr7:76625999-76626064 | *LINC03009, POMZP3* | Yes | Yes | Yes |
| chr7:100734168-100738613 | *ZAN* | Yes | Yes | Yes |
| chr7:100991656-100993197 | *MUC12* | Yes | Yes | Yes |
| chr7:142068646-142086717 | *MGAM* | Yes (chr7:142065334-142086717) | Yes | Yes |
| chr7:142091912-142094497 | *MGAM* | Yes | Yes | Yes |
| chr8:12137167-12138633 | *FAM66D, USP17L2* | Yes | Yes | Yes |
| chr9:42183645-42189734 | *SPATA31A6* | Yes | Yes | Yes |
| chr9:66917804-66918980 | *ZNF658* | Yes | Yes | Yes |
| chr9:66986986-66987520 | *SPATA31A3* | Yes (chr9:66986986-66989121) | Yes (chr9:66986986-66987520) | Yes |
| chr9:138219283-138219434 | *FAM157B* | Yes | Yes | Yes |
| chr10:46287262-46330084 | *ANTXRL, AGAP14P* | Yes (chr10:46287262-46337884) | Yes | Yes |
| chr10:122586059-122592595 | *DMBT1* | Yes (chr10:122579577-122592595) | Yes | Yes |
| chr10:133527356-133531734 | *CYP2E1* | Yes | Yes | Yes |
| chr10:133554843-133556047 | *SYCE1* | No, missing copy numbers | No | No |
| chr11:55603440-55651839 | *OR4C11, OR4P4, OR4S2* | Yes | Yes | No, genes had median TPM of 0 in all tissues |
| chr12:7872806-7933147 | *SLC2A14, SLC2A3* | Yes | Yes | Yes |
| chr12:8221821-8230646 | *FAM90A1, FAM66C, ZNF705A* | Yes (chr12:8176998-8230646) | Yes | No, poor copy number typing |
| chr12:10431116-10436174 | *KLRC2* | Yes | Yes | Yes |
| chr12:27495679-27502121 | *SMCO2* | Yes | Yes | Yes |
| chr15:43598826-43604152 | *CKMT1B, STRC* | No, missing copy numbers | No | No |
| chr15:43604649-43605399 | *STRC* | Yes | Yes | Yes (PCR+ only) |
| chr16:18531465-18562040 | *NOMO2* | Yes | Yes | Yes |
| chr16:22008301-22080880 | *MOSMO, LOC124903665* | Yes | Yes | No, poor copy number typing |
| chr16:28595781-28610191 | *SULT1A1, SULT1A2, LOC107984835* | Yes (chr16:28595548-28610191) | Yes | Yes |
| chr16:55810516-55820479 | *CES1* | Yes (chr16:55810516-55833055) | Yes | Yes |

| | | | | |
|---|---|---|---|---|
| chr16:75529148-75531062 | *CHST5* | Yes | Yes | Yes |
| chr17:36195285-36212569 | *CCL3L3, CCL4L2, LOC128966706* | Yes | Yes | Yes |
| chr17:36428925-36429179 | *TBC1D3F* | Yes | Yes | Yes |
| chr17:46094559-46172143 | *KANSL1, LOC107985027* | Yes | Yes | Yes |
| chr17:63894999-63910326 | *CSH1, CSHL1* | Yes | No, different copy number distributions between Releases 2 and 3 | Yes |
| chr19:4512617-4513160 | *PLIN4* | Yes | Yes | Yes (PCR+ only) |
| chr19:17332929-17339600 | *ANO8, GTPBP3* | Yes | Yes | Yes |
| chr19:40849817-40850426 | *CYP2A6, CYP2A7* | Yes (chr19:40849817-40881751) | Yes | Yes |
| chr19:43010269-43075632 | *PSG11, PSG2* | Yes | Yes | Yes |
| chr19:43257321-43259135 | *PSG9* | Yes | Yes | Yes |
| chr19:51643536-51646060 | *SIGLEC14, SIGLEC5* | Yes (chr19:51629832-51646640) | Yes | Yes |
| chr19:54803534-54821858 | *KIR2DL4, KIR3DL1* | Yes | Yes | Yes |
| chr21:13610228-13641566 | *POTED* | Yes | Yes | Yes |
| chr22:25327416-25360092 | *LRP5L* | Yes | Yes | Yes |
| chr22:38984074-38992488 | *APOBEC3B, APOBEC3B-AS1* | Yes | Yes | Yes |
| chr22:42126573-42130791 | *CYP2D6* | Yes | Yes | Yes |
| chr22:42512909-42556588 | *RRP7A, SERHL2, LOC124900479* | Yes (chr22:42516010-42556588) | Yes | Yes |
| chrX:1623106-1624453 | *ASMT* | Yes | Yes | Yes |
| chrX:48058467-48131419 | *ZNF630, ZNF630-AS1* | Yes (chrX:48058467-48060945) | Yes | Yes |
| chrX:104013169-104040404 | *H2BW1, H2BW2* | Yes | Yes | No, genes were not included in GTEx v8 |
| chrX:149715603-149716776 | *MAGEA11* | Yes | Yes | Yes |
| chrX:154150655-154156533 | *OPN1LW* | Yes | No, different copy number distributions between Releases 2 and 3 | No, poor copy number typing |
| Total | 74 | 71 | 69 | 63 |

**Supplementary table 2**: Demographic characteristics and genetic ancestry of the UK Biobank exome sequences used in this study, where available [6].

| | **Release 1 (n = 49,953)** | **Release 2 (n = 150,682)** | **Release 3 (n = 269,317)** | **Total (n = 469,952)** |
|---|---|---|---|---|
| Mean age, years | 56.6 | 56.4 | 56.6 | |
| Female, % | 54.6 | 55.3 | 53.6 | |
| Ancestry group | | | | |
| African | 919 | 1,943 | 3,578 | 6,440 |
| American | 118 | 323 | 529 | 970 |
| Central/South Asian | 1,009 | 3,085 | 4,622 | 8,716 |
| East Asian | 307 | 911 | 1,462 | 2,680 |
| European | 43,224 | 131,139 | 236,045 | 410,408 |
| Middle Eastern | 181 | 510 | 856 | 1,547 |
| Total | 45,758 | 137,911 | 247,092 | 430,761 |

**Supplementary table 3:** Number of participants included for autosomal PheWAS

| **Cohort** | **African** | **Central/South Asian** | **European** | **Total** |
|---|---|---|---|---|
| Discovery | 1,943 | 3,085 | 131,139 | 136,167 |
| Replication | 3,578 | 4,622 | 236,045 | 244,245 |
| Total | 5,521 | 7,707 | 367,184 | 380,412 |

**Supplementary table 4:** Number of participants included for chromosome X PheWAS,

| **Cohort** | **Inferred sex** | **African** | **Central/South Asian** | **European** | **Total** |
|---|---|---|---|---|---|
| Discovery | Female | 1,143 | 1,442 | 72,294 | 74,879 |
| | Male | 797 | 1,637 | 58,692 | 61,126 |
| Replication | Female | 2,041 | 2,083 | 126,025 | 130,149 |
| | Male | 1,505 | 2,509 | 109,741 | 113,755 |
| Total | | 5,486 | 7,671 | 366,752 | 379,909 |

**Supplementary Table 5:** Conditional analysis of mCNV and associated SNVs with binary and inverse normal transformed quantitative traits. All regressions were performed in Release 3 EUR participants (n = 236,045).

| Gene(s) | Phenotype | Selected SNV(s) | p-value for regression of mCNV to phenotype | p-value for regression of lead SNV to phenotype | Regression p-values with mCNV and SNV(s) as covariates | Variance inflation factors for mCNV and SNV |
|---|---|---|---|---|---|---|
| *RHD, RSRP1* | Blood potassium levels | rs3079633 | $7.38x10^{-10}$ | $2.53x10^{-9}$ | mCNV p=0.099<br>rs3079633 p=0.680 | mCNV=25.3*<br>rs3079633=25.3* |
| *GSTM1* | Cholesteryl esters in small HDL | rs140584594 | $2.52x10^{-10}$ | $1.02x10^{-11}$ | mCNV p=0.0290<br>rs140584594 p=0.000892 | mCNV=33.8*<br>rs140584594=33.8* |
| *GSTM1* | Cancer of urinary organs | rs140584594 | $5.10x10^{-9}$ | $6.49x10^{-9}$ | mCNV p=0.456<br>rs140584594 p=0.799 | mCNV=34.4*<br>rs140584594=34.4* |
| *ZAN* | Mean corpuscular volume | rs9801017,<br>rs41295942,<br>7:100261913_TCTGA_T,<br>rs117726678,<br>rs757075198,<br>rs1918352,<br>rs558587040,<br>rs200070847 | $4.32x10^{-37}$ | rs9801017 (p=$5.81x10^{-113}$)<br>rs41295942 (p=$1.70x10^{-24}$)<br>7:100261913_TCTGA_T (p=$2.92x10^{-15}$)<br>rs117726678 (p=$5.10x10^{-17}$)<br>rs757075198 (p=$5.11x10^{-10}$)<br>rs1918352 (p=$1.11x10^{-5}$)<br>rs558587040 (p=$3.61x10^{-5}$)<br>rs200070847 (p=0.028307) | mCNV = 0.783<br>rs9801017 = $1.61x10^{-10}$<br>rs41295942 = 0.00626<br>7:100261913_TCTGA_T = 0.652<br>rs117726678 = 0.0194<br>rs757075198 = 0.163<br>rs1918352 = 0.0445<br>rs558587040 = 0.0218<br>rs200070847 = 0.119 | mCNV = 3.02<br>rs9801017 = 2.08<br>rs41295942 = 1.03<br>7:100261913_TCTGA_T = 2.61<br>rs117726678 = 1.31<br>rs757075198 = 1.75<br>rs1918352 = 1.03<br>rs558587040 = 1.01<br>rs200070847 = 1.16 |
| *SULT1A1, SULT1A2* | eGFR | rs116938877 | $1.57x10^{-5}$ | $4.08x10^{-7}$ | mCNV p=$7.12x10^{-5}$<br>rs116938877 p=$1.91x10^{-6}$ | mCNV=1.01<br>rs116938877=1.01 |

*indicates variants which are highly colinear with each other, where the regression model included mCNV and SNV(s) (VIF > 5).

**Supplementary table 6:** Conditional analysis of mCNV and associated SNVs for quantitative traits, with raw phenotype measures. All regressions were performed in Release 3 EUR participants (n = 236,045).

| Gene(s) | Phenotype | Selected SNV(s) | p-value for regression of mCNV to phenotype | p-value for regression of individual SNV to phenotype | Regression p-values with mCNV and SNV as covariate | Variance inflation factors for mCNV and SNV(s) |
|---|---|---|---|---|---|---|
| *GSTM1* | Cholesteryl esters in small HDL | rs140584594 | $3.2x10^{-11}$ | $1.54x10^{-11}$ | mCNV p=0.0569<br>rs140584594 p =0.00197 | mCNV=33.7*<br>rs140584594=33.7* |
| *ZAN* | Mean corpuscular volume | rs9801017,<br>rs41295942,<br>7:100261913_TCTGA_T,<br>rs117726678,<br>rs757075198,<br>rs1918352,<br>rs558587040,<br>rs200070847 | $4.25x10^{-34}$ | rs9801017 (p=$2.24x10^{-107}$)<br>rs41295942 (p=$3.50x10^{-21}$)<br>7:100261913_TCTGA_T (p=$6.71x10^{-14}$)<br>rs117726678 (p=$4.80x10^{-17}$)<br>rs757075198 (p=$8.32x10^{-10}$)<br>rs1918352 (p=$6.80x10^{-5}$)<br>rs558587040 (p=4.04 $x10^{-4}$)<br>rs200070847 (p=0.029529) | mCNV = 0.720<br>rs9801017 = $8.79x10^{-10}$<br>rs41295942 = 0.00428<br>7:100261913_TCTGA_T = 0.609<br>rs117726678 = 0.0158<br>rs757075198 = 0.0943<br>rs1918352 = 0.0344<br>rs558587040 = 0.0418<br>rs200070847 = 0.137 | mCNV = 3.03<br>rs9801017 = 2.08<br>rs41295942 = 1.03<br>7:100261913_TCTGA_T = 2.61<br>rs117726678 = 1.31<br>rs757075198 = 1.75<br>rs1918352 = 1.03<br>rs558587040 = 1.01<br>rs200070847 = 1.16 |

*indicates variants which are highly colinear with each other, where the regression model included mCNV and SNV(s) (VIF > 5).

**Supplementary table 7:** Human Pangenome Reference Consortium samples used in this study

| Sample | Population descriptor | Population ancestry | Project |
|---|---|---|---|
| HG002 | Ashkenazi Jewish | European | Genome in a Bottle [55] |
| HG02622 | Gambian in Western Division, The Gambia - Mandinka | African | 1000 Genomes Project [44] |
| HG03453 | Mende in Sierra Leone | African | 1000 Genomes Project [44] |
| NA19240 | Yoruba in Ibadan, Nigeria | African | 1000 Genomes Project [44] |
| NA21309 | Maasai in Kinyawa, Kenya | African | HapMap Project [56] |

**Supplementary figure 1 An example copy number typing plot show clear integer cluster of copy numbers**

Copy number typing plot at the chr22:42126573-42130791 locus (*CYP2D6*) in the Release 2 cohort (n = 150,682). The points clustered along the lowest horizontal line represent individuals with a copy number of 0 and the highest cluster represents copy number of 8. This an example of a locus with clear separation between copy number clusters.

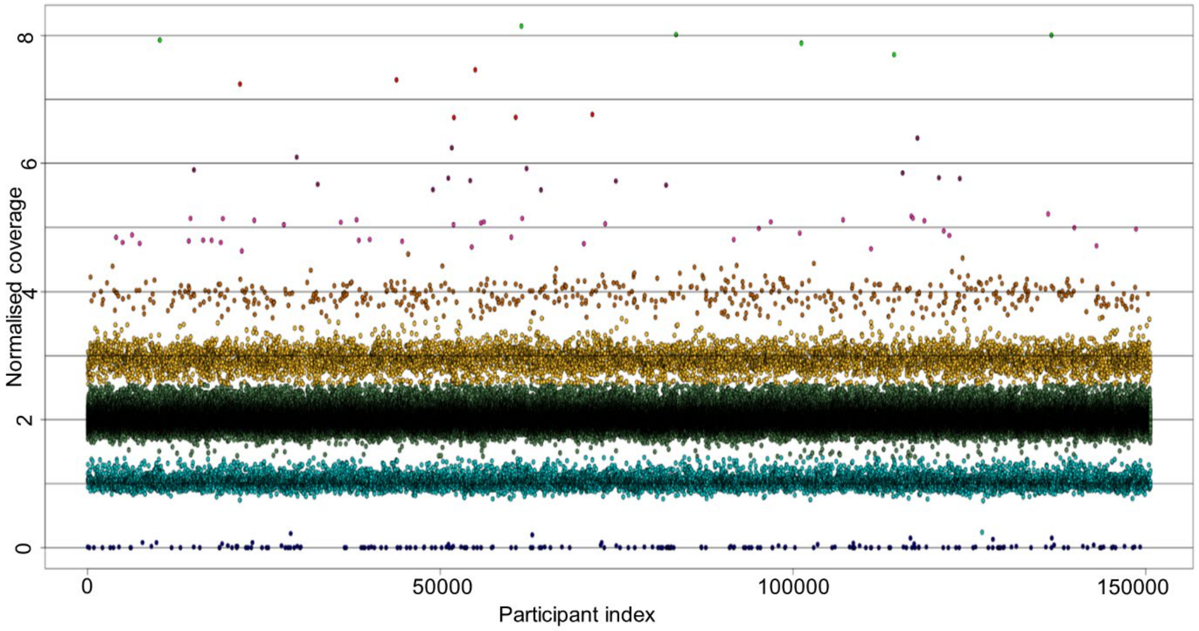

**Supplementary figure 2 An example of improving copy number calls by re-analysis**

a) The chr1:13321702-13371900 locus (PRAMEF14, PRAMEF15, PRAMEF19 genes) typed in the Release 2 cohort (n = 150,682). b) Default typing of the same locus in a batch of Release 3 samples (n = 48,355) results in clusters of samples split between two different copy numbers, as seen where a cluster is split between green (copy number 2) and yellow (copy number 3). c) Following re-analysis of b, clusters are clearly separated. The cluster which was previously split between two copy numbers is assigned as copy number 1 in light blue.

a)

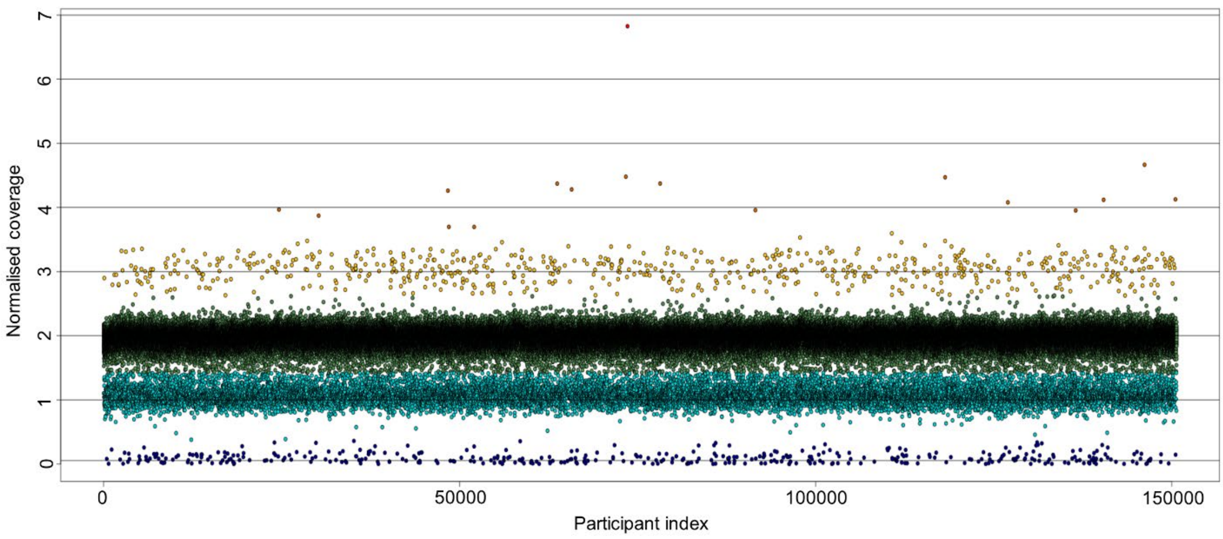


b)

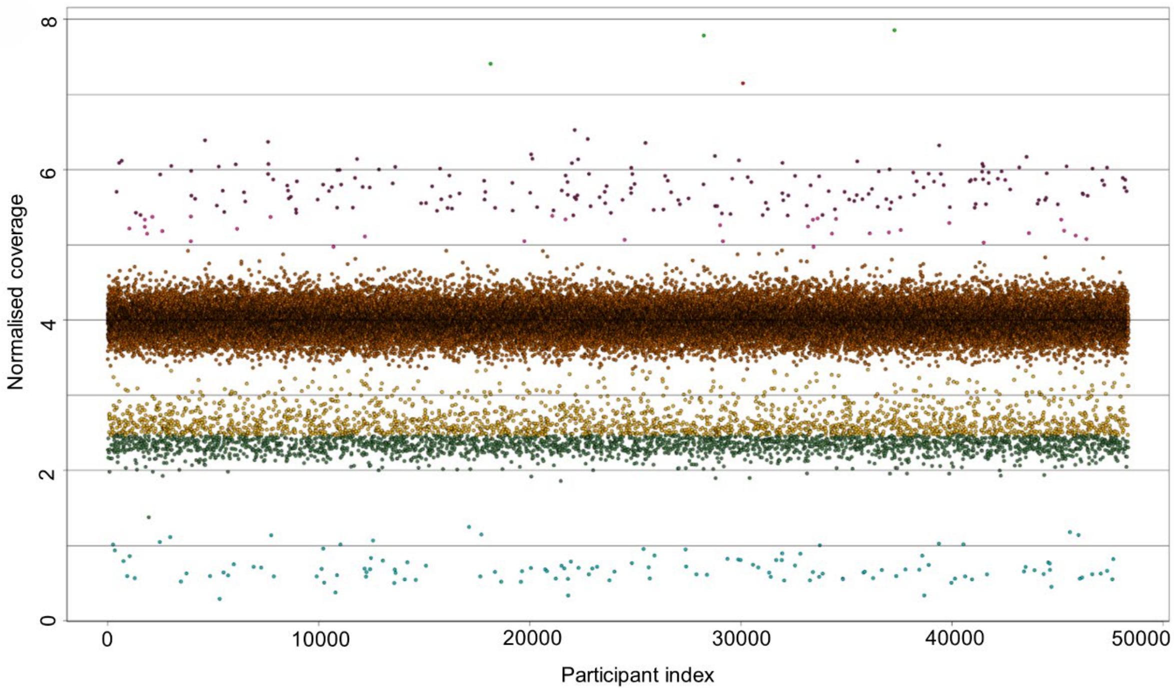


c)

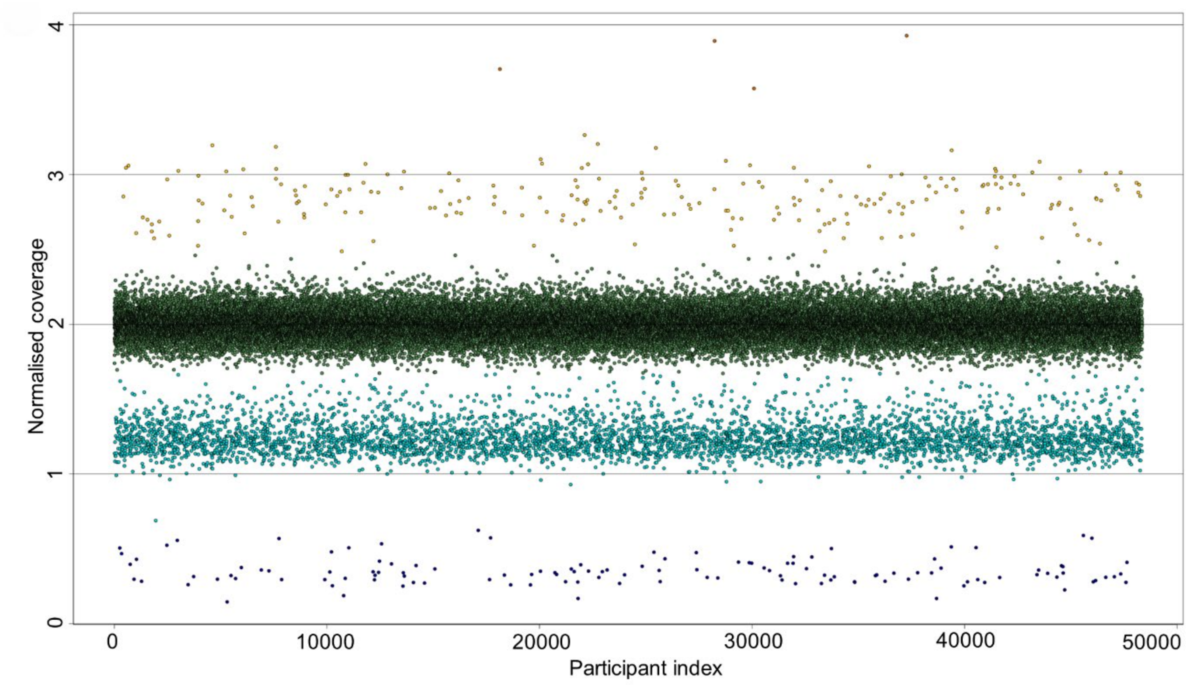
Normalised coverage
0
1
2
3
4
0
10000
20000
30000
40000
50000
Participant index

**Supplementary figure 3 An example of inconsistent copy number calls between Releases 2 and 3**

a) The chrX:154150655-154156533 locus (*OPN1LW* gene) typed in the Release 2 cohort (n = 150,682) with clearly defined copy number clusters. The lowest horizontal cluster represents copy number of 0 and the highest cluster represents copy number of 8. b) In a batch of the Release 3 exomes (n = 47,895), the lowest horizontal cluster represents copy number of 0 and the highest cluster represents copy number of 9. Although this locus was re-analysed to match the copy number distribution in the Release 2 exomes, the assignment of copy number to clusters is inconsistent. Copy number 1 (light blue) is split into 3 clusters, and copy number 3 (yellow) has not formed a clear cluster. This pattern of clustering was detected in all of the Release 3 genotyping batches.

a)

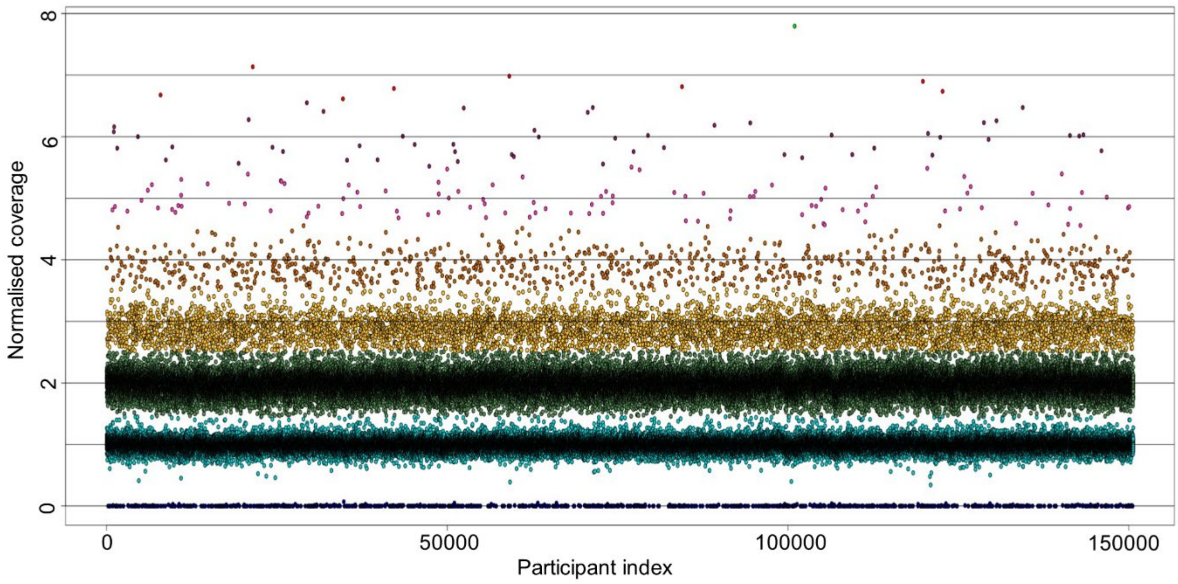


b)

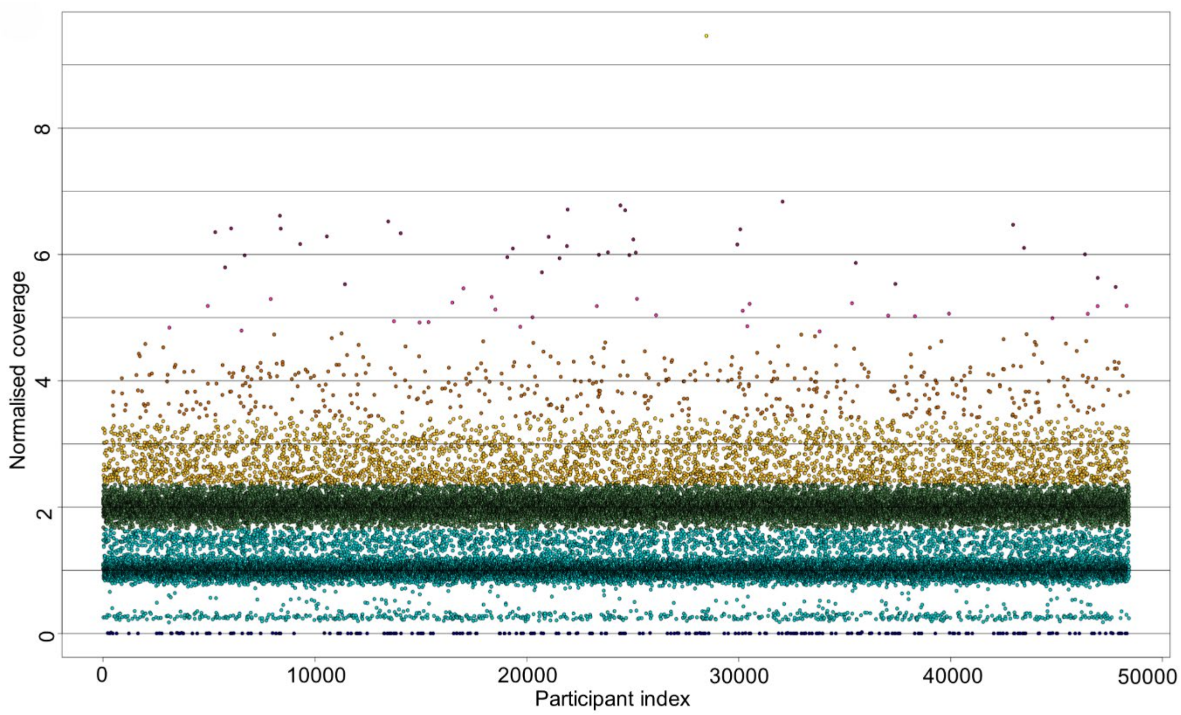

Normalised coverage
0
2
4
6
8
0
10000
20000
30000
40000
50000
Participant index

**Supplementary figure 4** **Analysis of *RHD* deletion and duplication using whole genome sequence**

Read coverage from whole genome sequence data in the chr1:25256032-25345866 region, spanning the *RHD* gene, visualised on IGV. a) Data from two individuals identified from WES as having a diploid copy number 0. Blue arrows indicate the breakpoints of the deletion, which are predicted by decreased read coverage relative to the surrounding sequence. The patches of increased read depth, including sequences mapping to ~3kb of the final intron of *RHD,* are due to sequence read mismapping from highly-similar sequence within the *RHCE* gene, likely originating from a recent gene conversion events. b) Data from four 3 copy individuals. c) Data from two 4 copy individuals. Duplication boundaries predicted by increased read coverage relative to the surrounding sequence in b and c are inconsistent and possibly reflect distinct copy number alleles. The segmental duplication track from UCSC genome browser has been inset above the gene track. The Rhesus boxes, which sponsor non-allelic homologous recombination generating RHD copy number alleles, are represented by the two yellow blocks which flank the gene, showing 98 - 99% similarity. The yellow box spanning the entire RHD gene bar the final exon represents one copy of a segmental duplication, the other carrying the RHCE gene. The grey segmental duplications map to other chromosomes (90 - 98% similarity).

a)

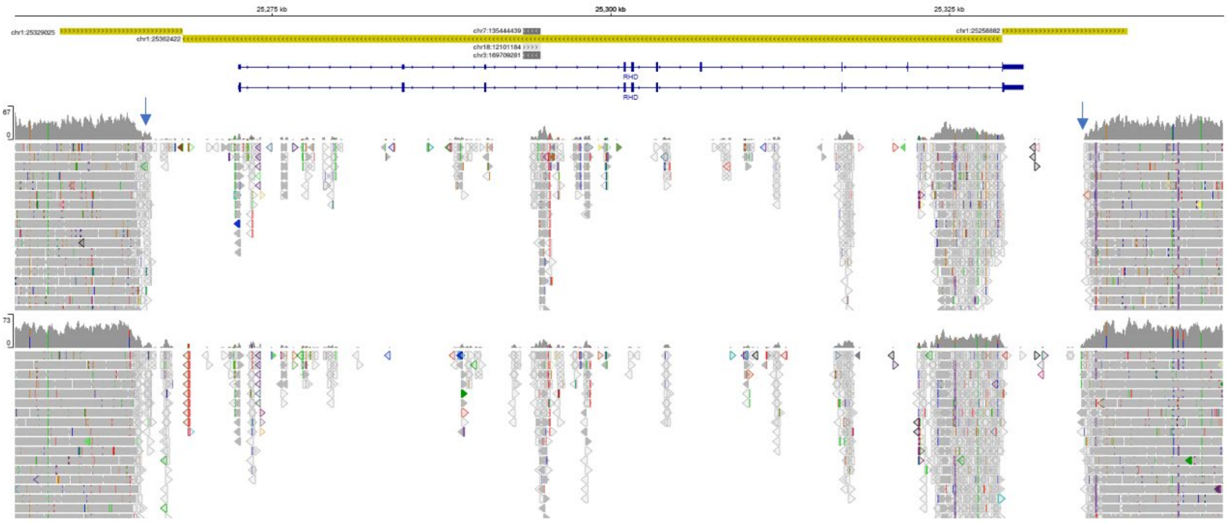

b)

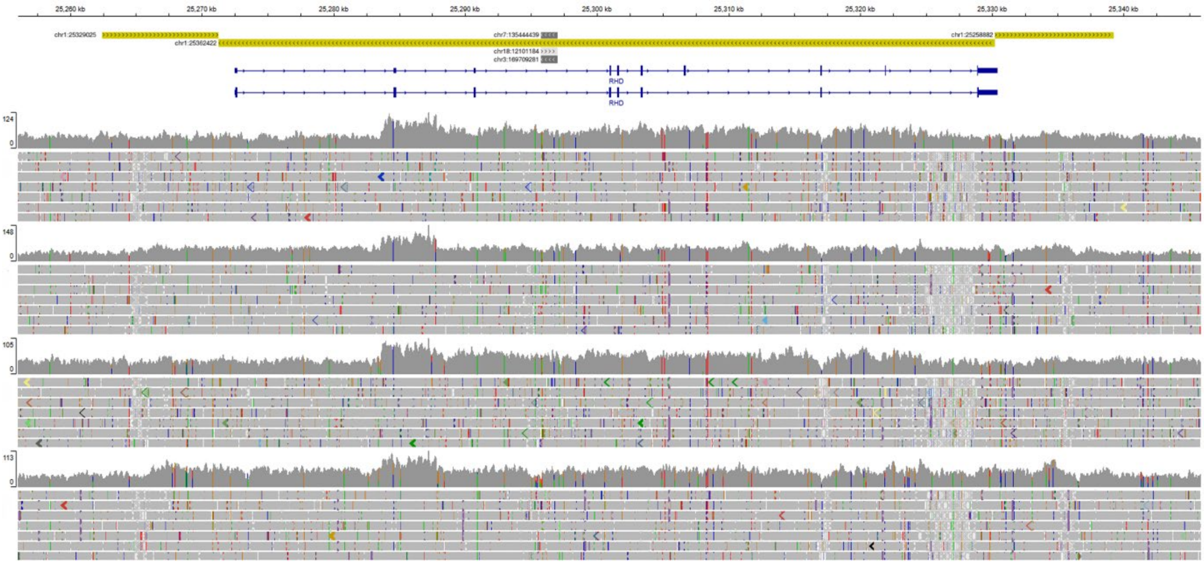
25,260 kb
25,270 kb
25,280 kb
25,290 kb
25,300 kb
25,310 kb
25,320 kb
25,330 kb
25,340 kb
chr1:25329025
chr1:25362422
chr7:135444439
chr18:12101184
chr3:169709281
chr1:25256882
RHD
RHD
124
0
148
0
105
0
113
0

c)

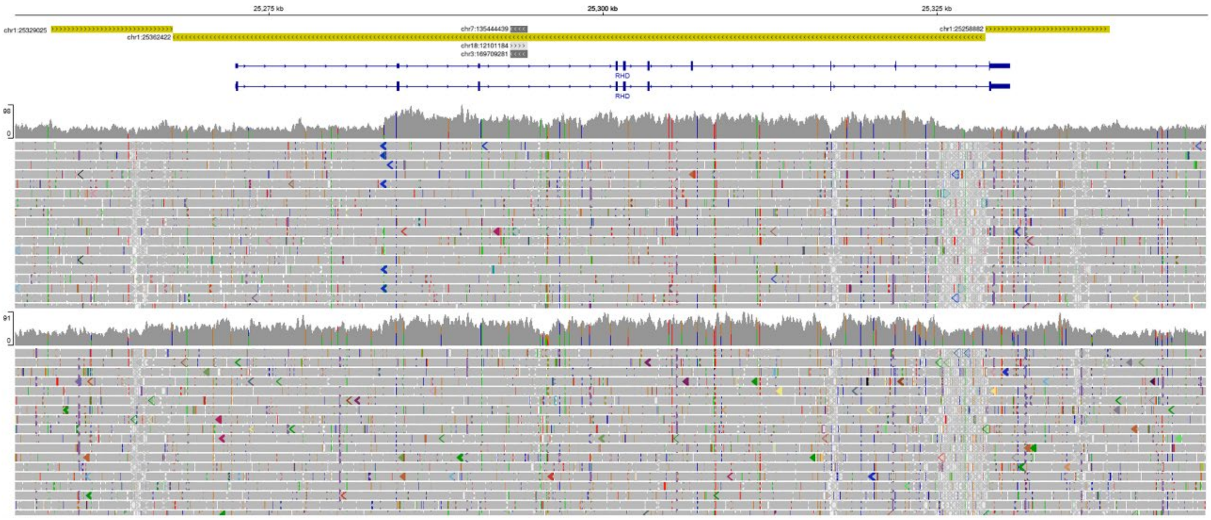
25,275 kb
25,300 kb
25,325 kb
chr1:25329025
chr1:25362422
chr7:135444439
chr18:12101184
chr3:169709281
chr1:25256882
RHD
RHD
98
0
91
0

**Supplementary figure 5** **Analysis of *AMY2B* amplification using whole genome sequence**

Sequence read depth from whole genome sequence data in the chr1:103550177-103735000 region from four individuals with diploid copy number 8 in the predicted *AMY2B* mCNV, shown on IGV. The segmental duplication track from UCSC genome browser has been inset above the gene track. Orange segmental duplications are >99% similar, while grey segmental duplications are 90 - 98% similar.

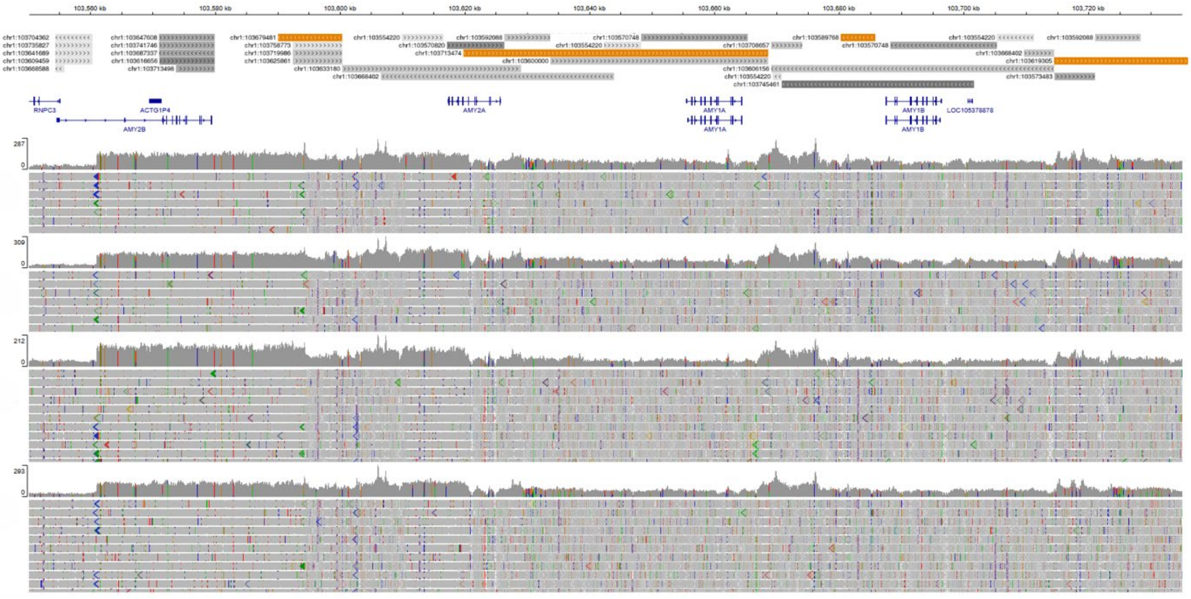

**Supplementary figure 6** **Analysis of *GSTM1* mCNV using whole genome sequence**

Read coverage from whole genome sequence data in the chr1:109672556-109710514 region from a) individuals with diploid copy number 0 and b) individuals with diploid copy number 4 in the *GSTM1* mCNV. The CNV breakpoints were predicted by decreased read coverage relative to the surrounding sequence. Arrows indicate likely boundaries of the deletion or duplication. The segmental duplication track from UCSC genome browser has been inset above the gene track. The grey segmental duplications which flank *GSTM1* are 90 - 98% similar.

a)

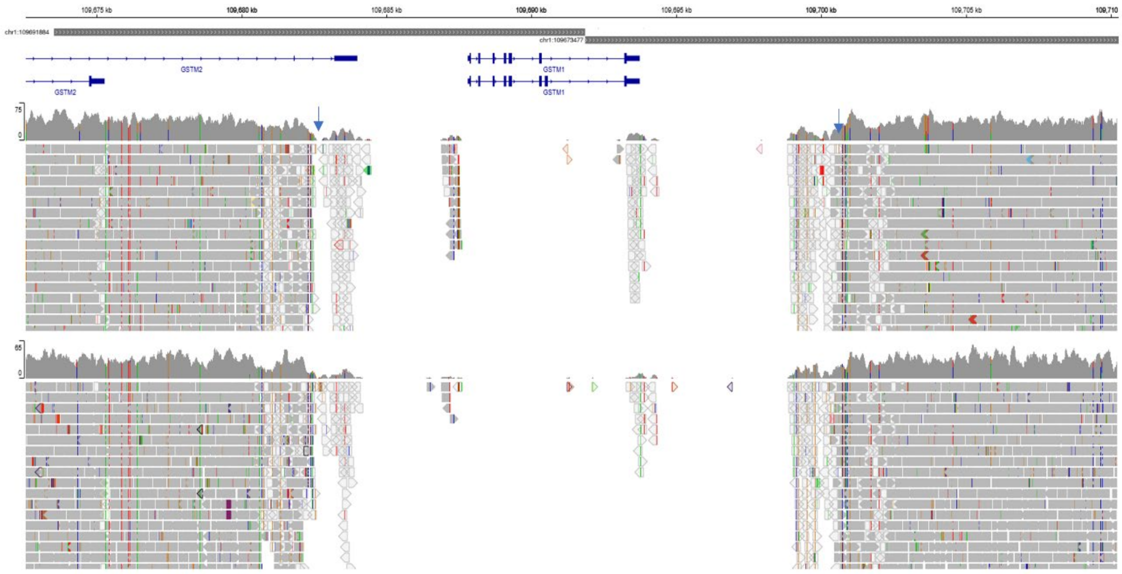


b)

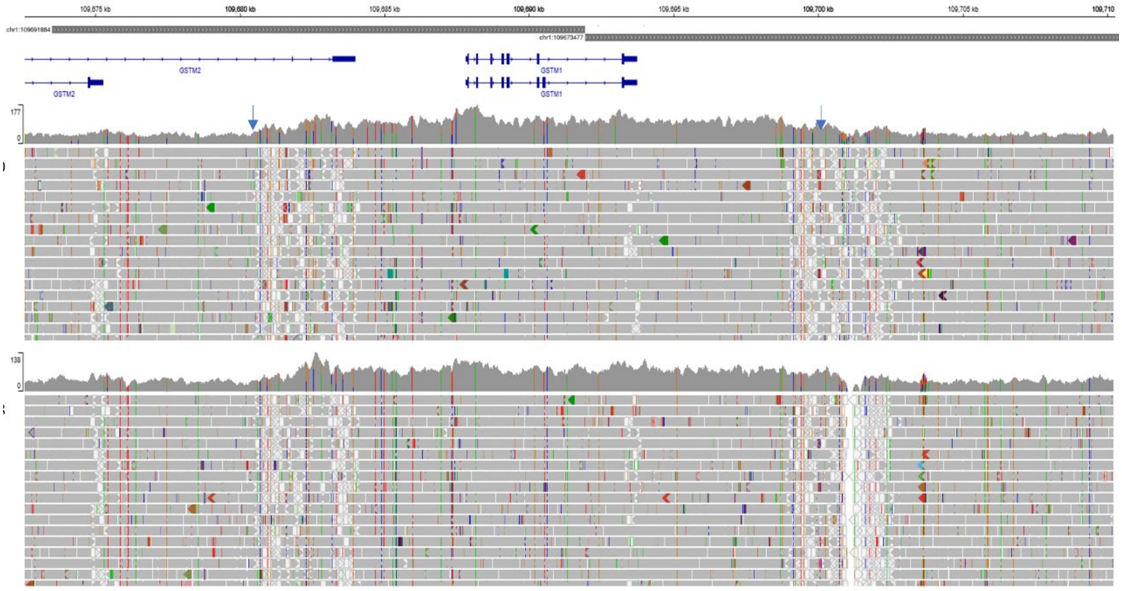

**Supplementary figure 7 Analysis of GOLPH3 deletion and duplication using whole genome sequence**

Read coverage from whole genome sequence data in the chr5:32096077-32178303 region, spanning the GOLPH3 gene, visualised on IGV. a) Data from individuals with diploid copy number 6 and b) individuals with diploid copy number 1 in the *GOLPH3* mCNV. The CNV breakpoints were predicted by increased or decreased read coverage relative to the surrounding sequence. Arrows indicate the breakpoints of the duplication. There appear to be two deletions in this region, which are separate from the duplication. No segmental duplication has been detected in this region.

a)

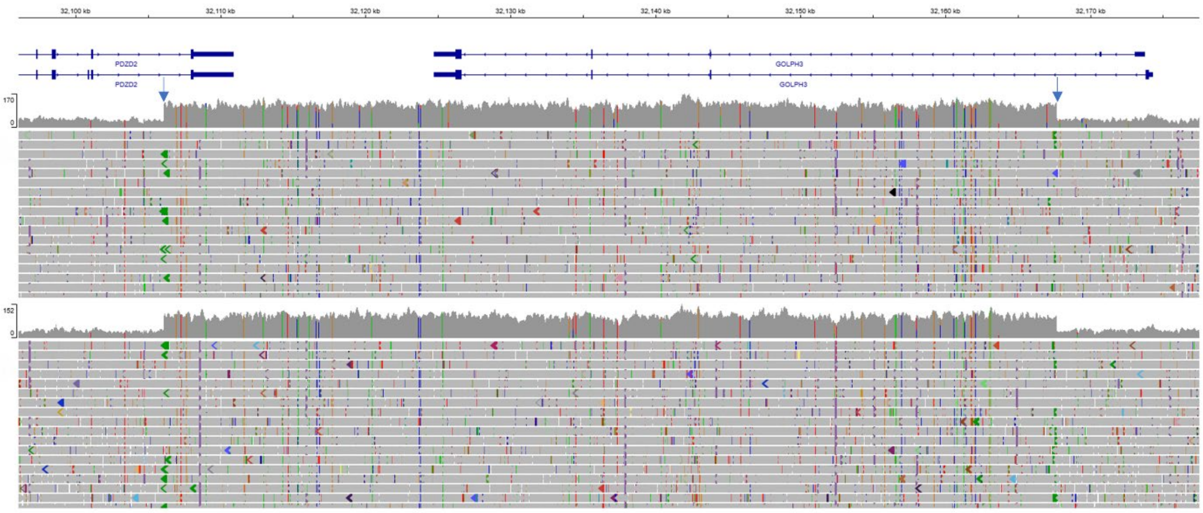


b)

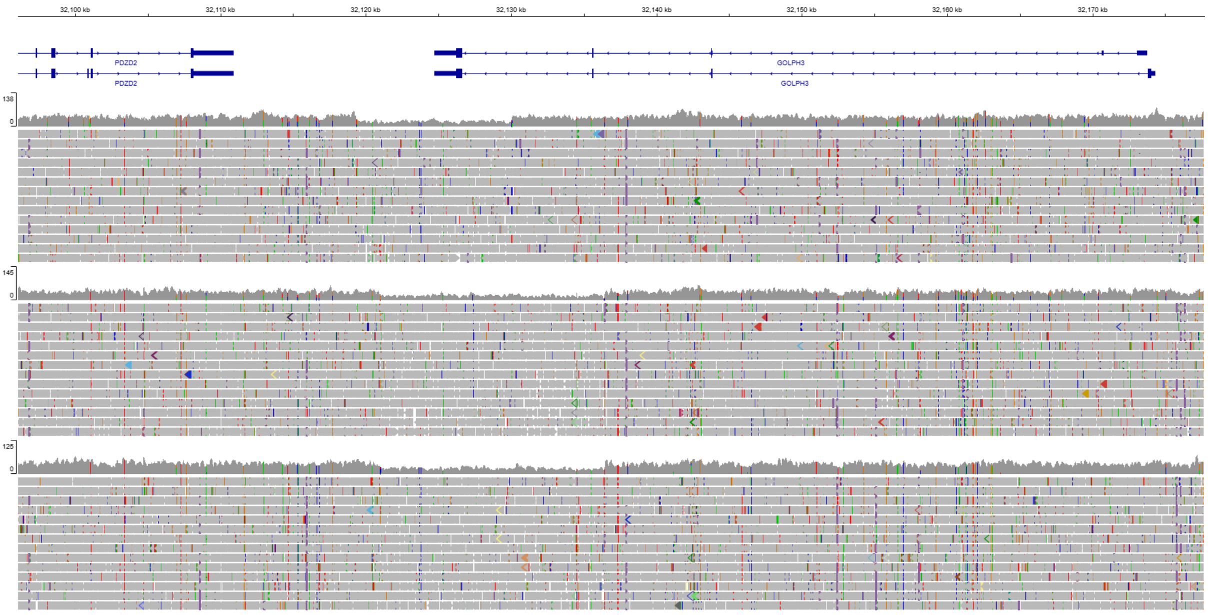

**Supplementary figure 8 Analysis of ZAN deletion and duplication using whole genome sequence**

Read coverage from whole genome sequence data in the chr7:100719965-100753273 region, spanning the *ZAN* gene, visualised on IGV. a) Data from individuals with diploid copy number 0 and b) individuals with diploid copy number 3 in the *ZAN* mCNV. The CNV breakpoints were predicted by decreased read coverage relative to the surrounding sequence. Arrows indicate the breakpoints of the deletion. The breakpoints of the duplication could not be visually predicted. No segmental duplication has been detected in this region.

a)

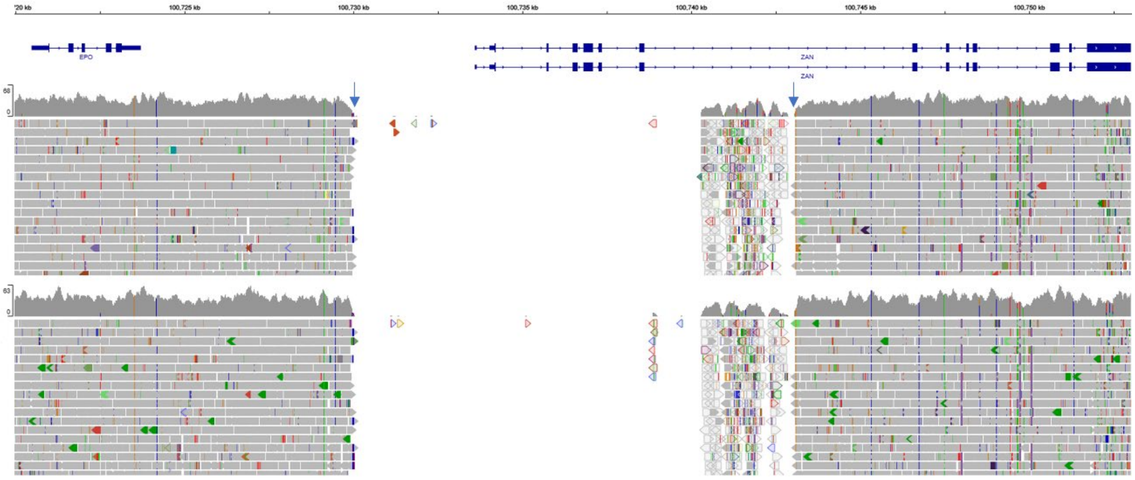

b)

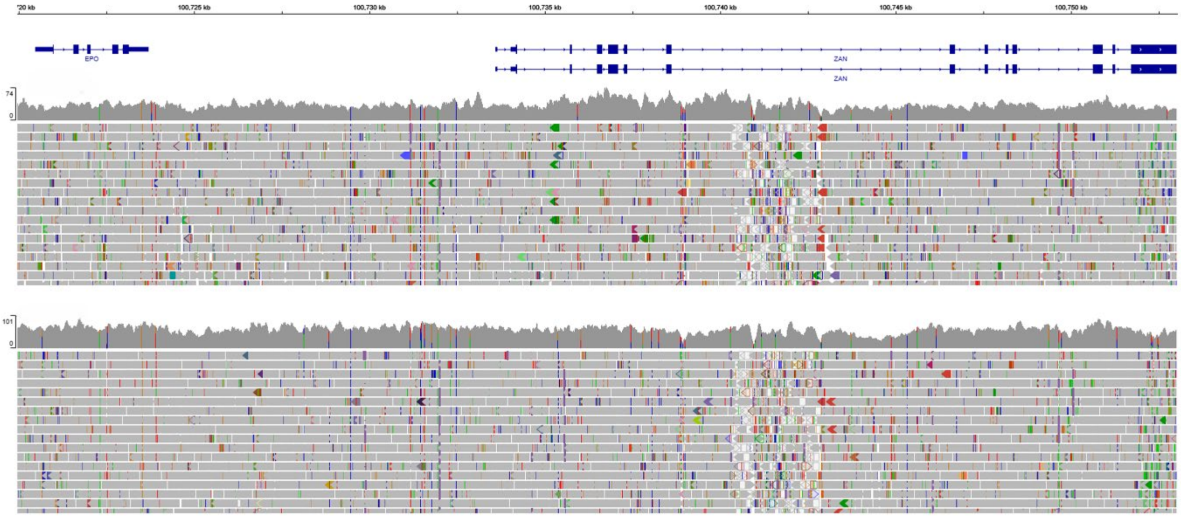

**Supplementary figure 9** **Analysis of *SULT1A1* mCNV using whole genome sequence**

Read coverage from whole genome sequence data in the chr16:28592638-28621039 region, spanning the *SULT1A1* gene, visualised on IGV. a) Data from individuals with diploid copy number 0 and b) the highest predicted diploid copy number (the top sample has copy number 11 and the bottom sample had copy number 8) in the predicted *SULT1A1/SULT1A2* mCNV. The CNV breakpoints were predicted by increased and decreased read coverage relative to the surrounding sequence. Arrows indicate the breakpoints of the deletion and duplication. The segmental duplication track from UCSC genome browser has been inset above the gene track. The grey segmental duplications are 90 - 98% similar.

a)

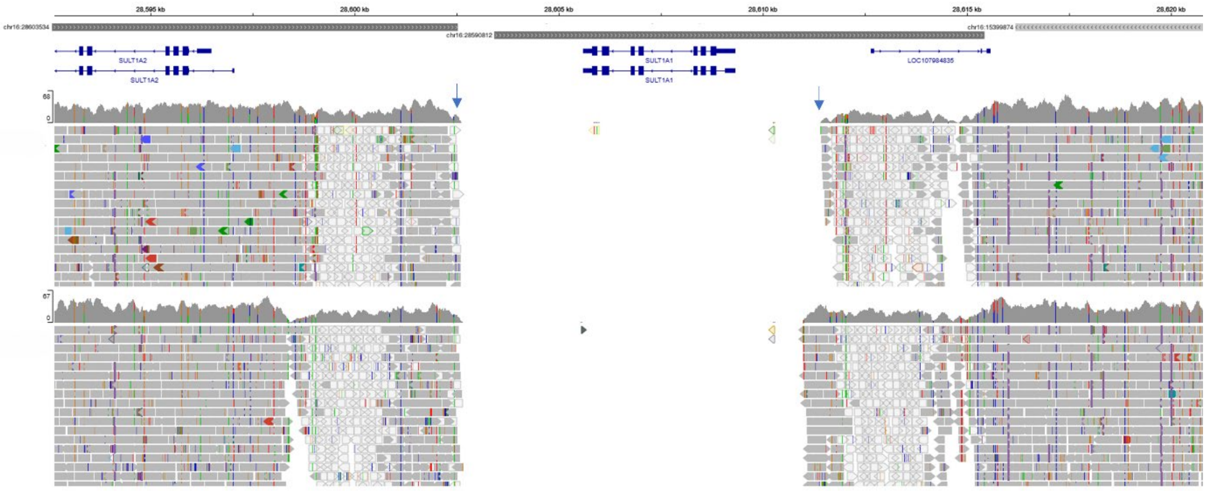


b)

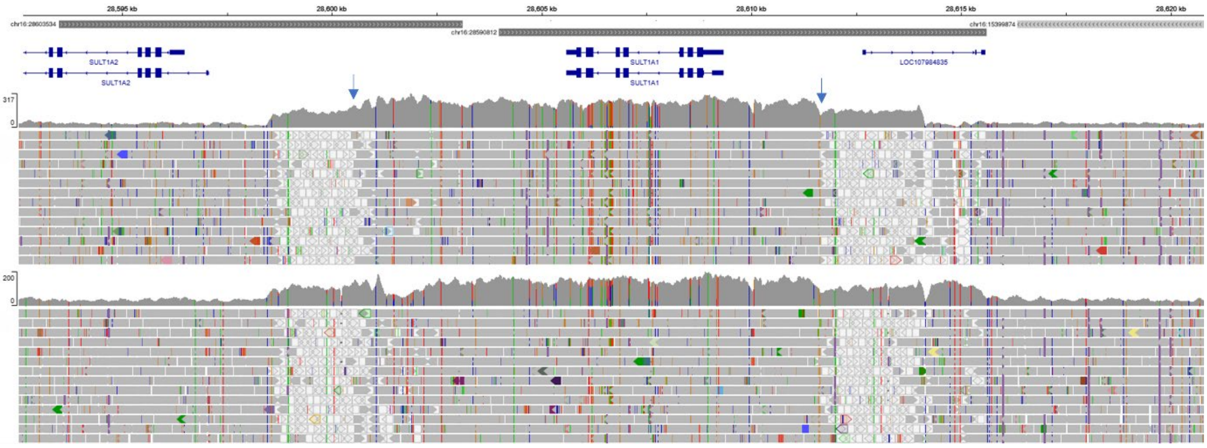

**Supplementary figure 10 Dot plot showing extent of *RHD* deletion**

a) Dot plot of T2T-CHM13 *RHD* region (x axis) compared with the HG03453 maternal assembly (y axis), to estimate the position of the *RHD* deletion. The dotted rectangle indicates the extent of the deletion. b) UCSC Genome browser annotation for T2T-CHM13 (chr1:25098780-25176740).

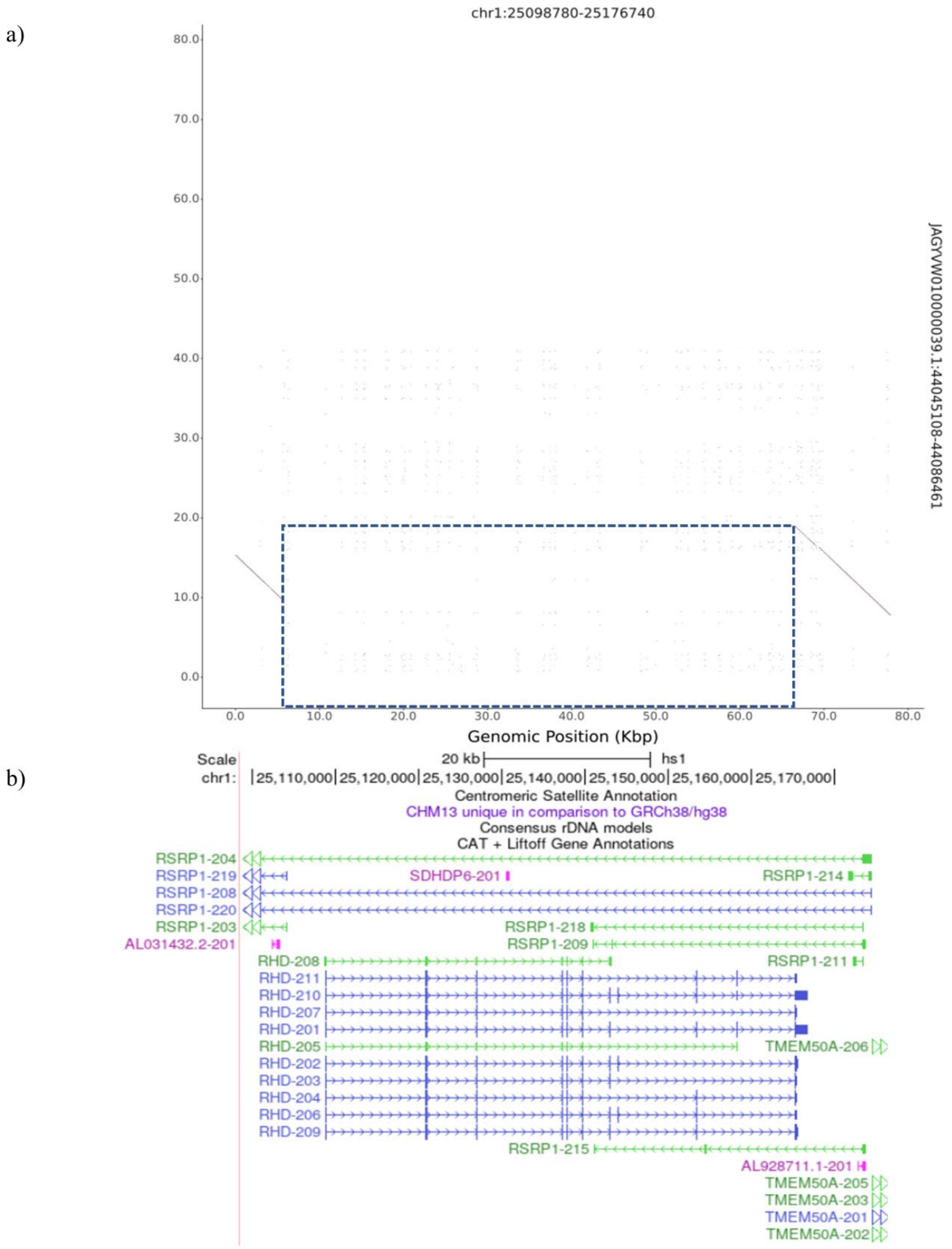

**Supplementary figure 11** **SVbyeye plot showing *RHD* deletion**

HG01123 paternal assembly showing *RHD* duplication aligned to T2T-CHM13 (top).

**Supplementary figure 12 Dot plot showing extent of *RHD* duplication**

T2T-CHM13 *RHD* region (x axis) compared with the reverse complement of the HG01106 paternal assembly (y axis). The dotted rectangle indicates the duplicated region. b) b) UCSC Genome browser annotation for T2T-CHM13 (chr1:25098780-25176740).

a)

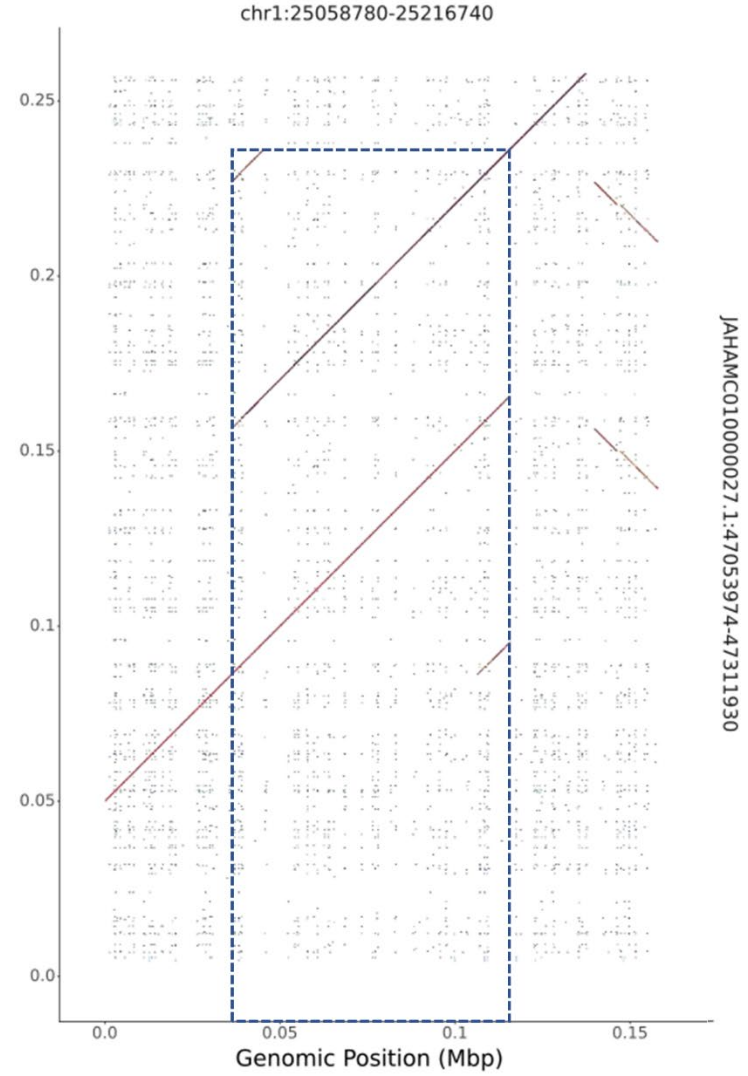


b)

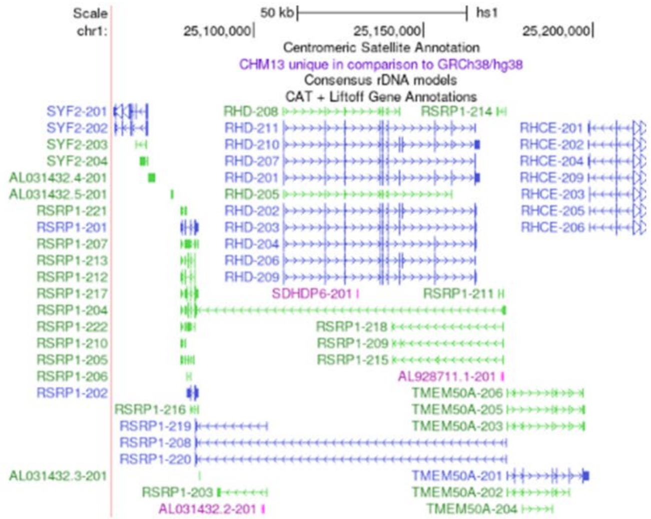

**Supplementary figure 13** **SVbyeye plot showing *RHD* duplication**

HG01106 paternal assembly (bottom) showing *RHD* duplication aligned to T2T-CHM13 (top).

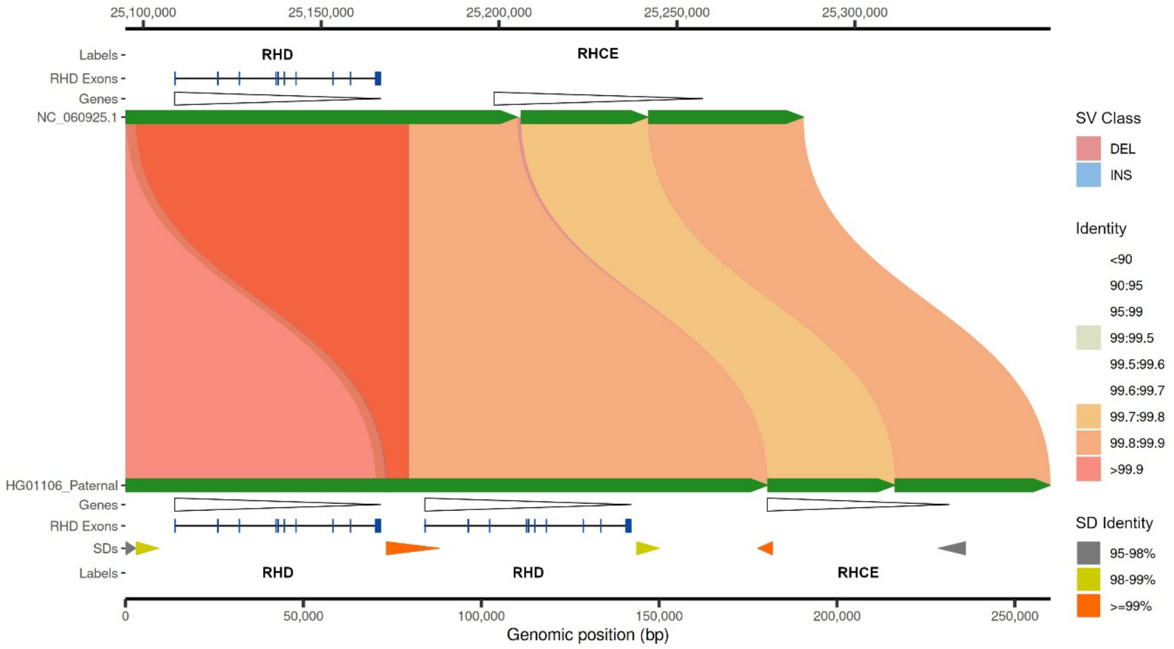

**Supplementary figure 14** **Dot plot showing extent of *AMY2B* duplication**

a) T2T-CHM13 amylase region (x axis) compared with the reverse complement of the HG00741 paternal assembly (y axis). The dotted rectangle indicates the duplicated region. b) UCSC Genome browser annotation for T2T-CHM13 (chr1:103376007-103955680).

a)

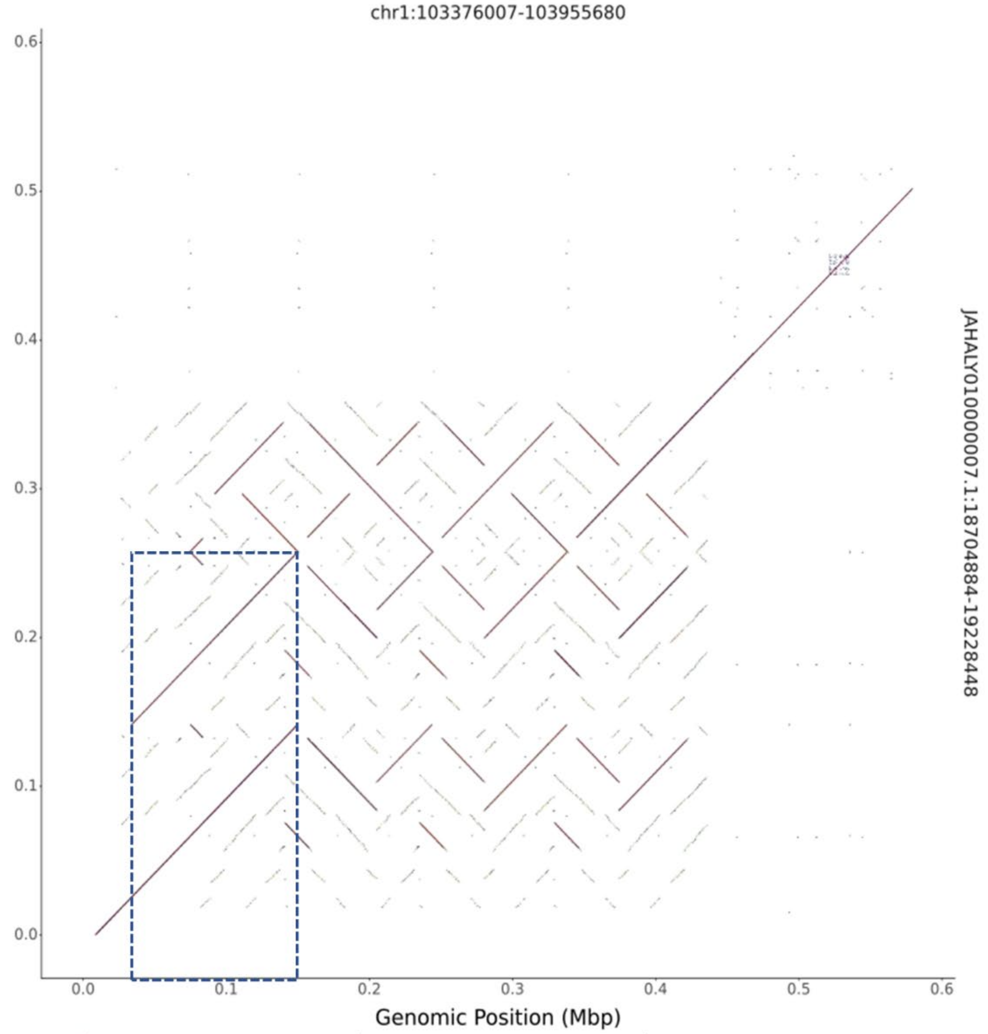


b)

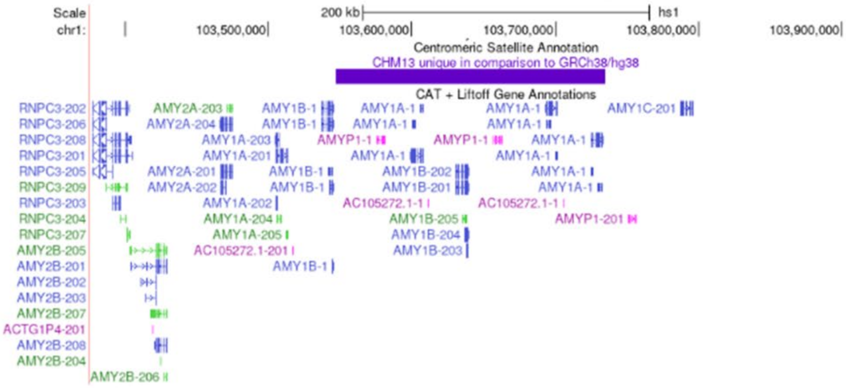

**Supplementary figure 15 Dot plot showing extent of *GSTM1* deletion**

a) GRCh38 *GSTM1*region (x axis) compared with the reverse complement of the HG02622 paternal assembly (y axis). The dotted rectangle indicates the deletion region. b) UCSC Genome browser annotation for GRCh38 (chr1:109677817-109703745). c) UCSC Genome browser annotation for the HG02622 paternal assembly (JAHAOO010000002.1:22898188-22925723)

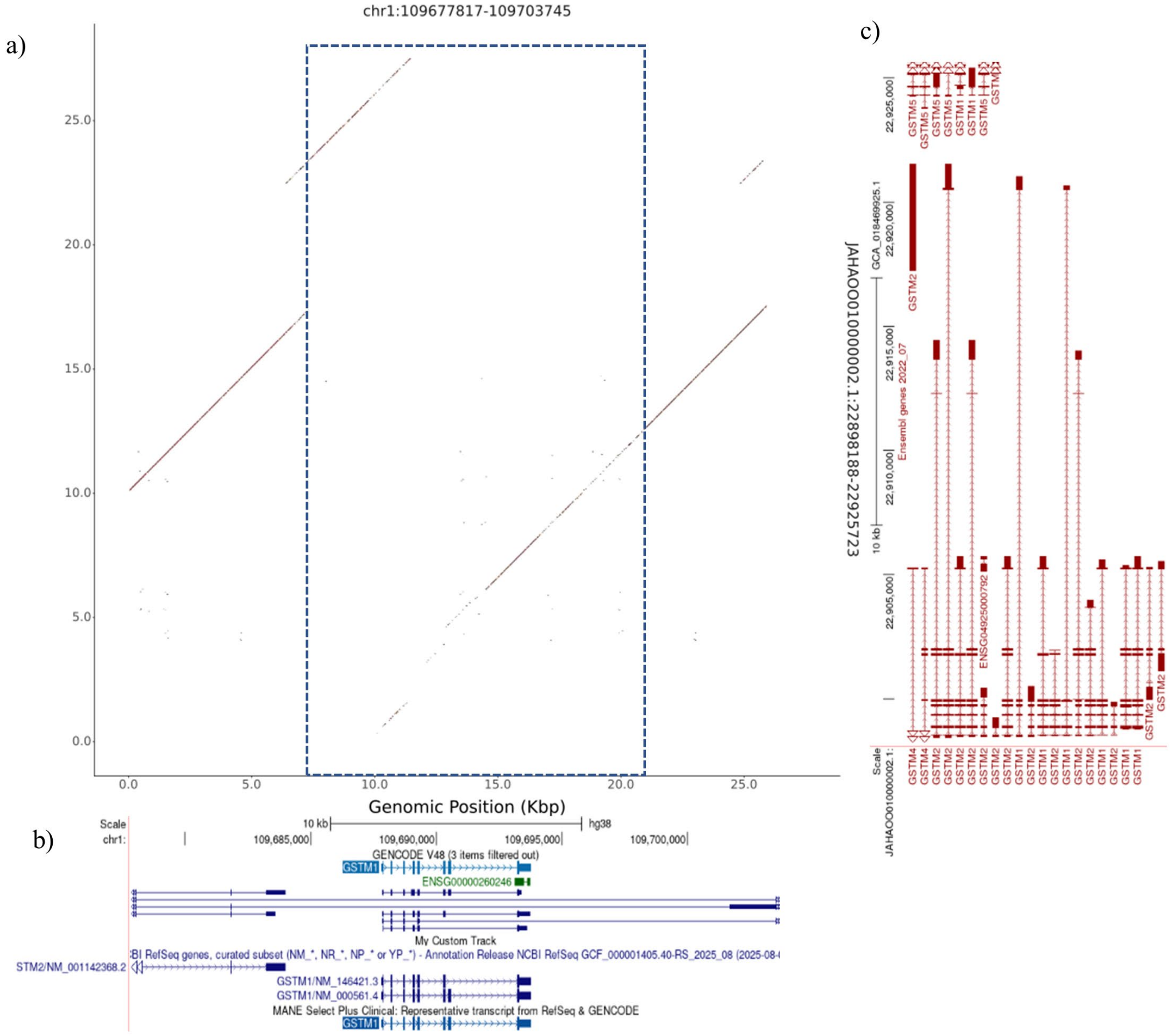

**Supplementary figure 16** **Dot plot showing extent of *GSTM1* duplication**

a)GRCh38 *GSTM1*region (x axis) compared with the reverse complement of the NA19240 paternal assembly (y axis). The dotted rectangle indicates the duplicated region. b) UCSC Genome browser annotation for GRCh38 (chr1:109677817-109703745).

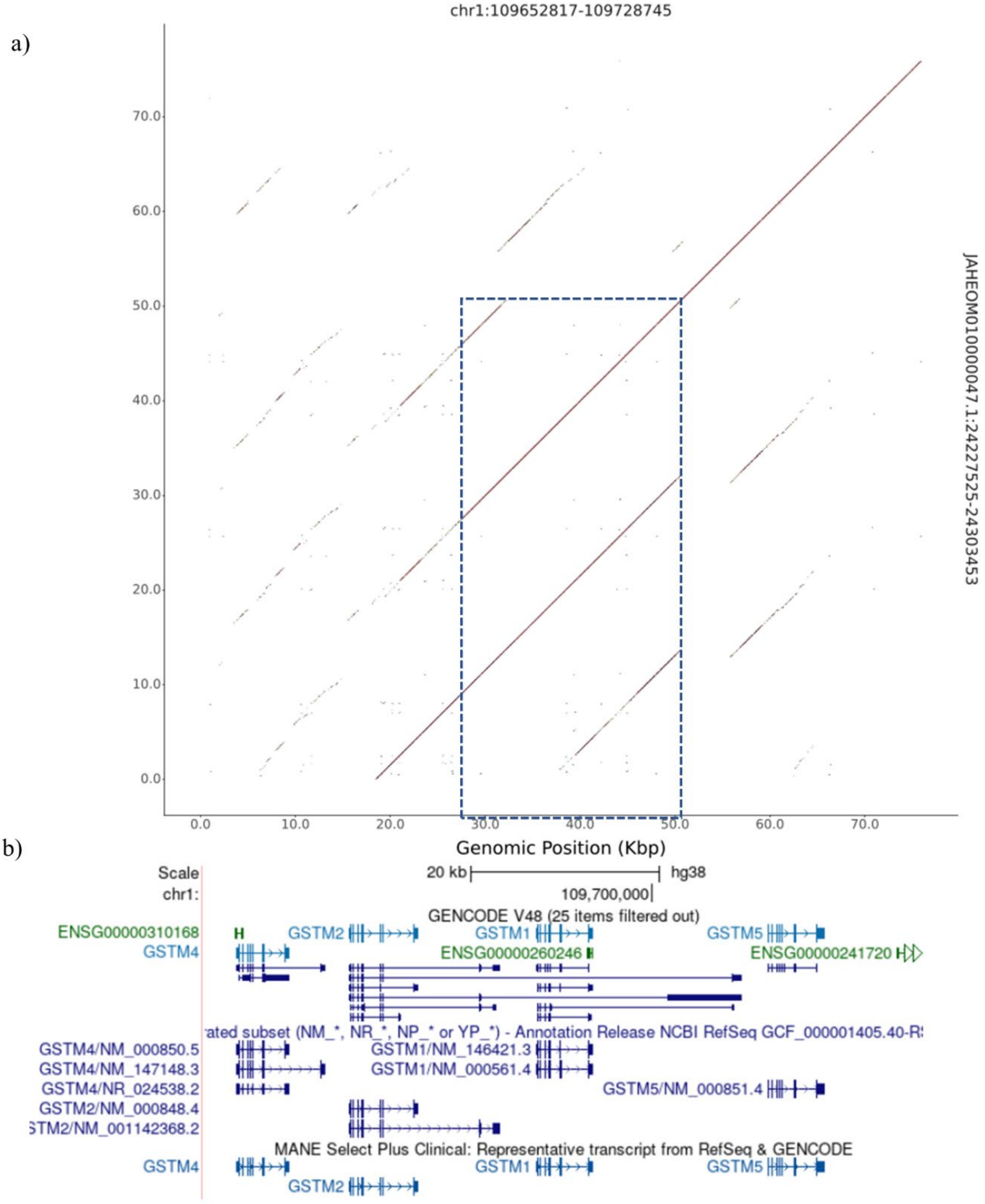

**Supplementary figure 17** **Svbyeye plot showing the deletion of GSTM1.**

HG002 maternal assembly (bottom) showing GSTM1 deletion when aligned to GRCh38 (top).

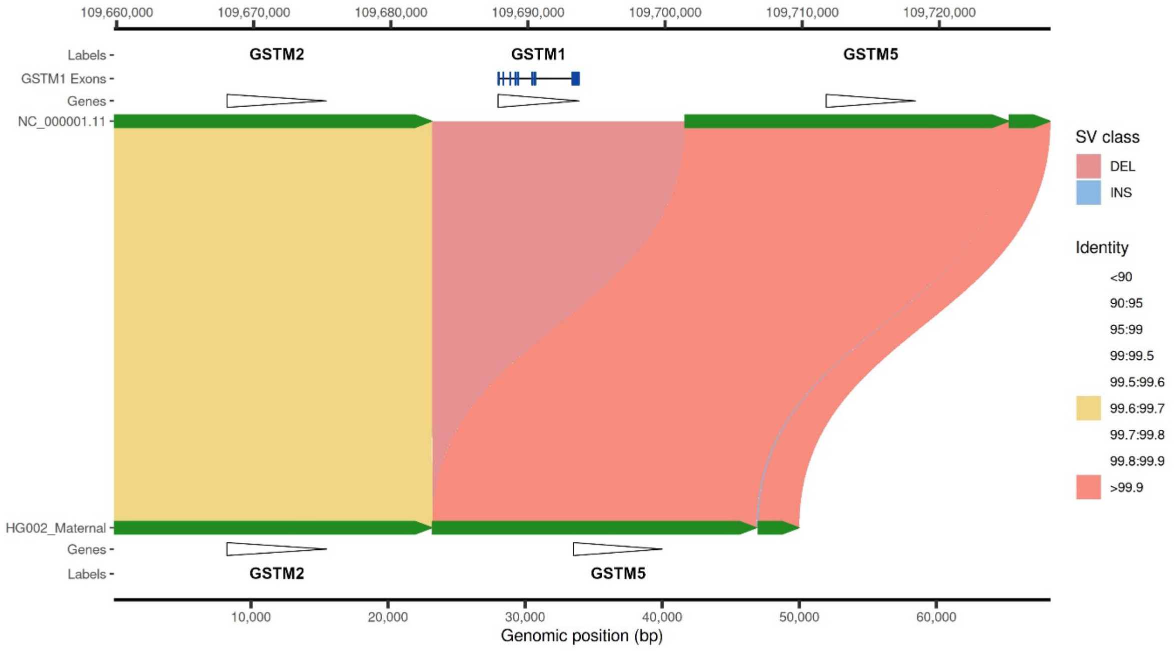

**Supplementary figure 18** **SVbyeye plot showing *GSTM1* duplication**

Duplication of *GSTM1* in the NA19240 paternal assembly (bottom) when aligned to GRCh38 (top). Segmental duplications are also annotated here. The large duplication event is accompanied by a smaller deletion.

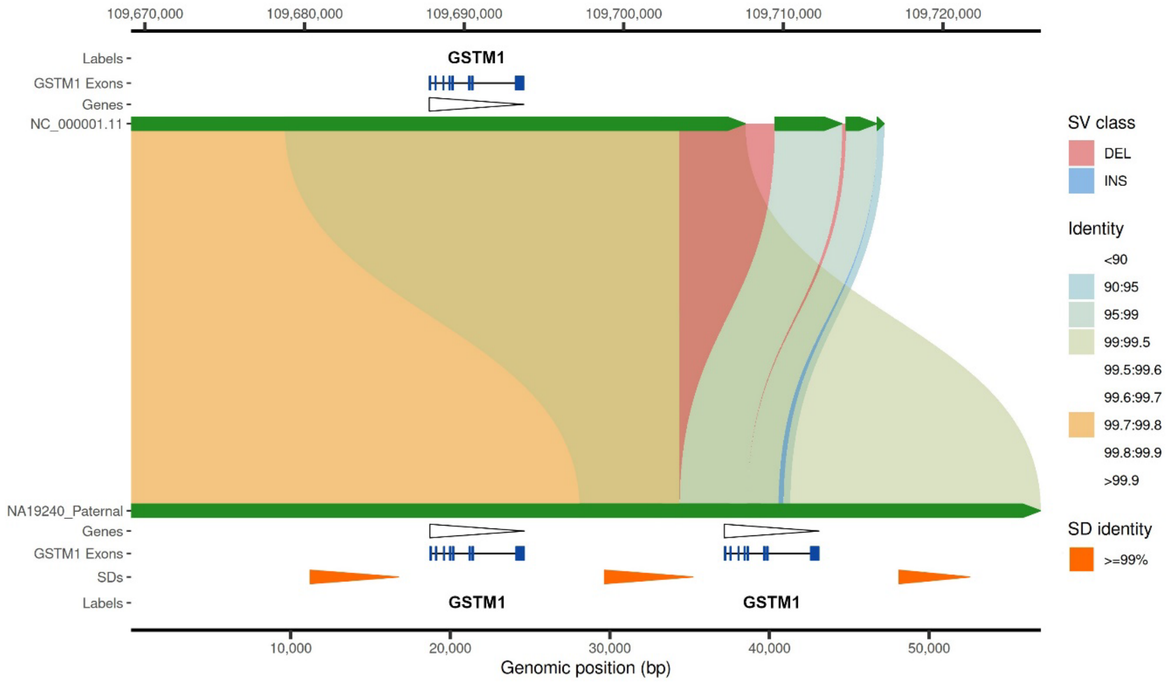

**Supplementary figure 19 Dot plot showing extent of *ZAN* deletion**

a) T2T-CHM13 *ZAN* region (x axis) compared with the HG002 paternal assembly (y axis). The dotted rectangle indicates the deleted region, which aligns to the first 7 exons of *ZAN*. b) UCSC genome browser annotation for T2T-CHM13 (chr7:101963655-102047894).

a)

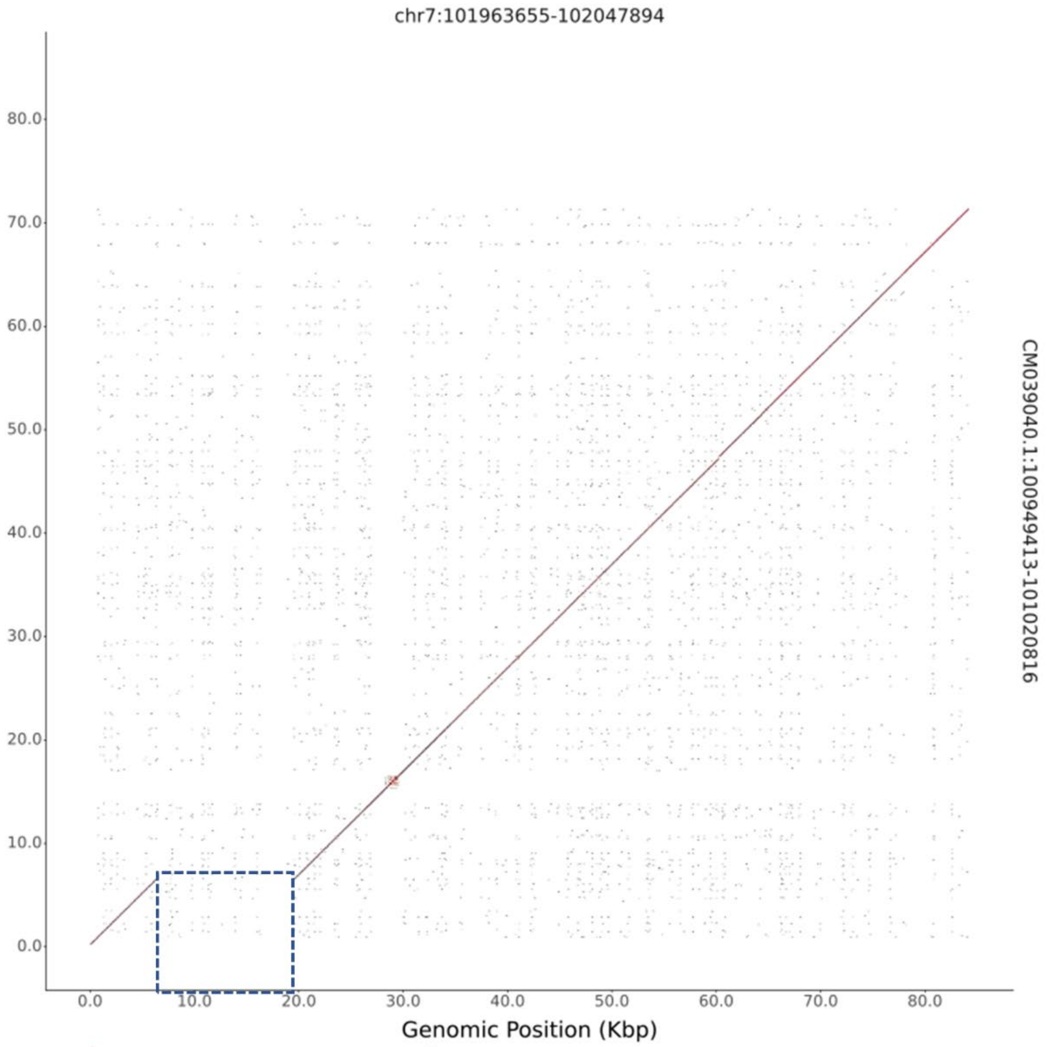


b)

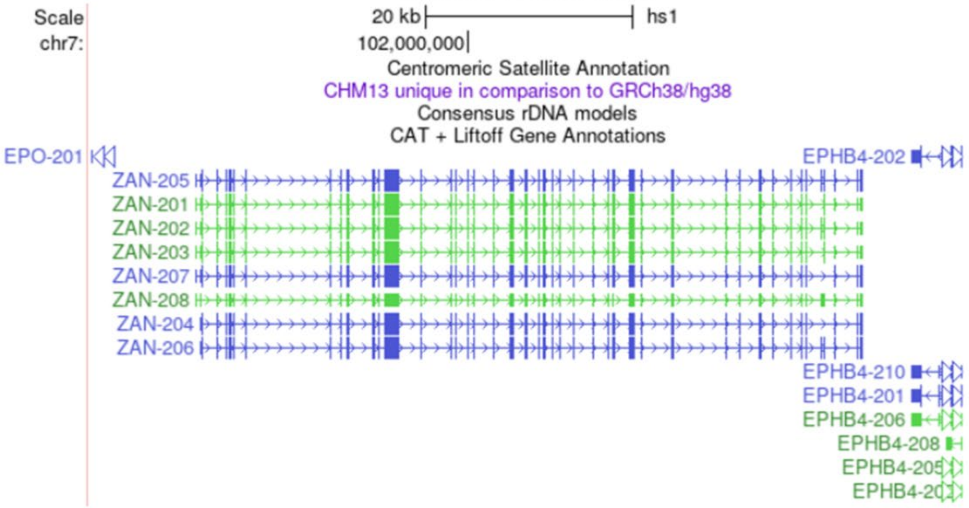

**Supplementary figure 20 Dot plot showing extent of *SULT1A1* duplication**

a) T2T-CHM13 *SULT1A1* region (x axis) compared with the reverse complement NA21309 maternal assembly (y axis). The dotted rectangle indicates the position of the duplication, which aligns to the entire *SULT1A1* gene. b) UCSC genome browser annotation for T2T-CHM13 (chr16:28760654-28815437).

a)

b)

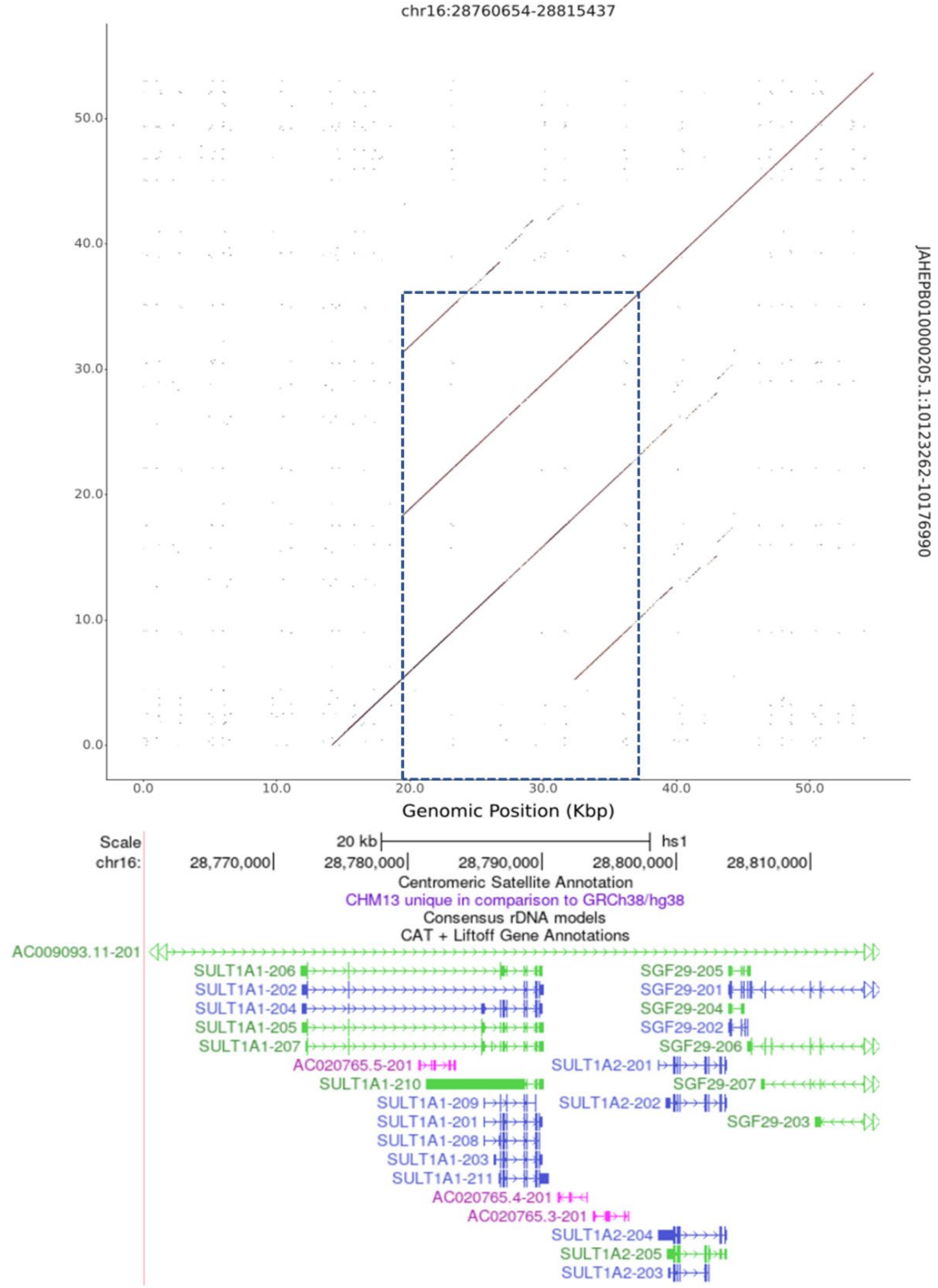

**Supplementary figure 21** **Svbyeye plot showing *SULT1A1* duplication**

HG02514 maternal assembly (bottom) showing *SULT1A1* duplication aligned to T2T-CHM13 (top).

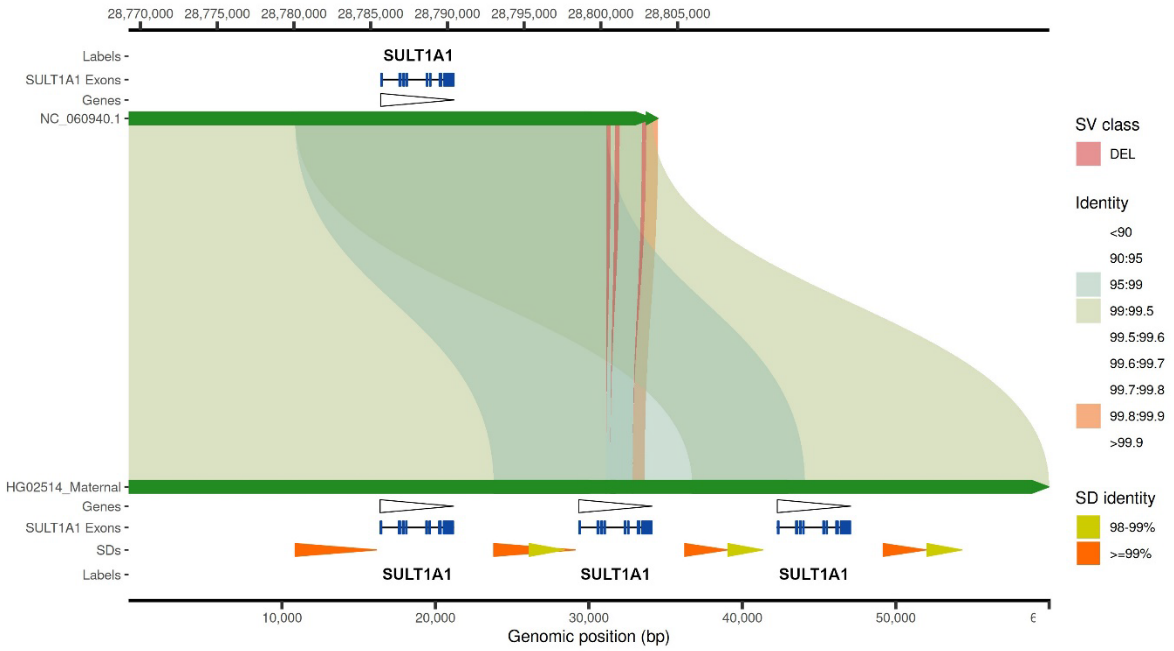

**Supplementary figure 22 Correlation of expression and copy number for all mCNVs**

Correlation between biological molecule and mCNV copy number, for all tissues, for each gene. The top row is based on UK Biobank proteomic data from blood plasma, other rows are GTEx transcript-level data from RNAseq. Light grey tiles for UK Biobank proteomic data indicate that the corresponding protein was not included in the data. For the expression data: dark grey tiles indicate that there was no difference in copy number as well as no difference in expression level between samples; green tiles indicate that there was no difference in copy number between samples; and turquoise indicates that there was no difference in expression level between samples. The *FCGR2C* and *MGAM* genes were annotated on two separate mCNVs each. *FCGR2C* region 1 is chr1:161589510-161591451 and region 2 is chr1:161595511-161673229. *MGAM* region 1 is chr7:142065334-142086717 and region 2 is chr7:142091912-142094497.

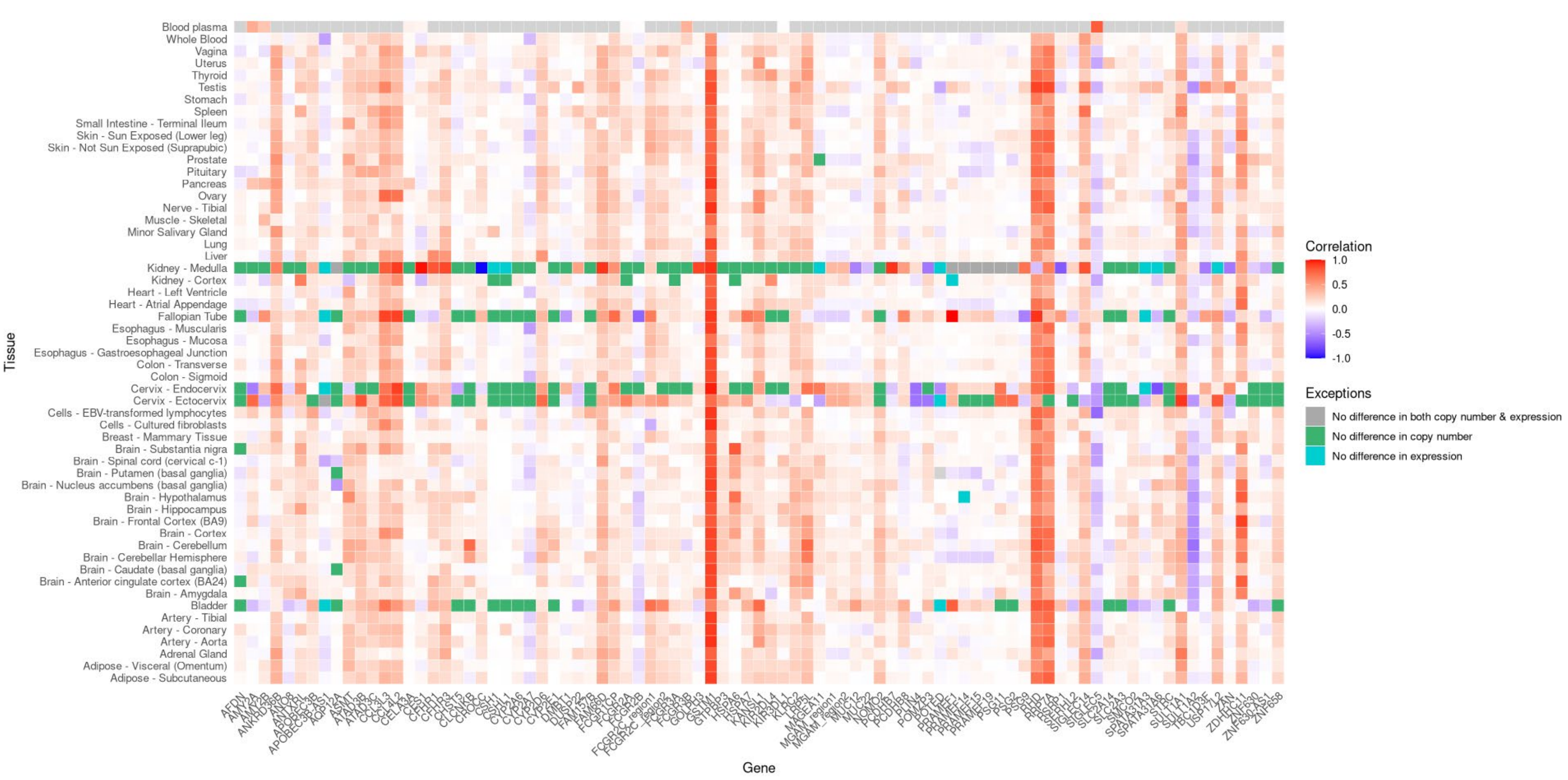
Blood plasma
Whole Blood
Vagina
Uterus
Thyroid
Testis
Stomach
Spleen
Small Intestine - Terminal Ileum
Skin - Sun Exposed (Lower leg)
Skin - Not Sun Exposed (Suprapubic)
Prostate
Pituitary
Pancreas
Ovary
Nerve - Tibial
Muscle - Skeletal
Minor Salivary Gland
Lung
Liver
Kidney - Medulla
Kidney - Cortex
Heart - Left Ventricle
Heart - Atrial Appendage
Fallopian Tube
Esophagus - Muscularis
Esophagus - Mucosa
Esophagus - Gastroesophageal Junction
Colon - Transverse
Colon - Sigmoid
Cervix - Endocervix
Cervix - Ectocervix
Cells - EBV-transformed lymphocytes
Cells - Cultured fibroblasts
Breast - Mammary Tissue
Brain - Substantia nigra
Brain - Spinal cord (cervical c-1)
Brain - Putamen (basal ganglia)
Brain - Nucleus accumbens (basal ganglia)
Brain - Hypothalamus
Brain - Hippocampus
Brain - Frontal Cortex (BA9)
Brain - Cortex
Brain - Cerebellum
Brain - Cerebellar Hemisphere
Brain - Caudate (basal ganglia)
Brain - Anterior cingulate cortex (BA24)
Brain - Amygdala
Bladder
Artery - Tibial
Artery - Coronary
Artery - Aorta
Adrenal Gland
Adipose - Visceral (Omentum)
Adipose - Subcutaneous
Tissue
AFDN
AMY2A
AMY2B
ANKRD36B
ANO8
ANTXRL
APOBEC3B
APOBEC3B-AS1
AQP12A
ASMT
ATAD3B
ATAD3C
CCL3L3
CCL4L2
CELA3A
CES1
CFHR1
CFHR3
CHST5
CLCNKB
CROCC
CSH1
CSHL1
CYP2A6
CYP2A7
CYP2D6
CYP2E1
DMBT1
DUSP22
FAM157B
FAM66D
FCGR1CP
FCGR2A
FCGR2B
FCGR2C_region1
FCGR2C_region2
FCGR3A
FCGR3B
GOLPH3
GSTM1
GTPBP3
HSPA6
HSPA7
KANSL1
KIR2DL4
KIR3DL1
KLRC2
LRP5L
MAGEA11
MGAM_region1
MGAM_region2
MUC12
MUC22
NOMO2
PCDHB7
PCDHB8
PLIN4
POMZP3
POTED
PRAMEF1
PRAMEF14
PRAMEF15
PRAMEF19
PSG11
PSG2
PSG9
RHD
RRP7A
RSRP1
SERHL2
SIGLEC14
SIGLEC5
SLC2A14
SLC2A3
SMCO2
SPATA31A3
SPATA31A6
STRC
SULT1A1
SULT1A2
TBC1D3F
USP17L2
ZAN
ZDHHC11
ZNF630
ZNF630-AS1
ZNF658
Gene
Correlation
1.0
0.5
0.0
-0.5
-1.0
Exceptions
No difference in both copy number & expression
No difference in copy number
No difference in expression
